# Why epidemic risk at the 2026 World Cup may not be what you think

**DOI:** 10.64898/2026.05.28.26354384

**Authors:** Justin Lessler, Claire P. Smith, Praachi Das, Abagael L. Sykes, Alessandra Urbinati, Kelly Geith, Kimberly A. Powers, Jessica T. Davis, Stephanie C. Kern-Allely, George G. Vega Yon, Eric T. Lofgren, Carl A.B. Pearson, Alessandro Vespignani

## Abstract

**Background:** The 2026 FIFA World Cup may bring over one million visitors to North America from around the globe to participate in mass gathering events. The nature of the event and recent news have raised concerns for some that the tournament could lead to infectious disease outbreaks or fuel existing epidemics.

**Objective:** To systematically assess the infectious disease threat posed to the United States by the tournament.

**Design:** A multi-institutional team evaluated pathogen-specific risk across three dimensions: importation, outbreak potential, and impact to identify a priority pathogen list. A systematic screening protocol ensured common criteria and that pathogen information was collected when necessary to inform inclusion.

**Results:** Increased risk from the World Cup is near zero for 63 of 77 evaluated pathogens. Pathogens were predominantly excluded as threats due to low excess importation risk and low outbreak potential if introduced. The remaining priority pathogens fall into five categories: (a) mosquito borne pathogens with the potential for sustained transmission in some host cities, (b) seasonal respiratory viruses, (c) chronic infections with high prevalence outside the United States, (d) pathogens present in the United States with likely increased transmission at World Cup activities, and (e) high-consequence infectious threats.

**Limitations:** Data availability is variable across diseases. Impact calculations may not reflect actual costs to host cities. Disease incidence in World Cup travelers may differ from national incidence rates.

**Conclusion:** While infectious disease outbreaks at the 2026 FIFA World Cup are possible, in an already highly connected world where large gatherings are frequent, the elevated risk from the tournament is not as extreme as it first may seem.

**Primary Funding Source:** US Centers for Disease Control and Prevention

## Introduction

Because the 2026 FIFA World Cup will bring over one million visitors from around the world to North America to participate in large gatherings, it has raised concern for potential infectious disease outbreaks^1^. These concerns have been heightened due to the recent outbreak of Andes-lineage hantavirus on a cruise ship^2^ and an expanding outbreak of Ebola in central Africa ^3^. In response, many localities and groups have developed lists of potential pathogens of concern, but these are often expansive, with unclear criteria and minimal accounting for existing underlying travel patterns. Hence, they may give a false impression of broadly increased infectious disease risk around the tournament, and low risk during “normal” times.

While there are many examples of past infectious disease outbreaks associated with sports tournaments and other mass gathering events, the number of outbreaks is low compared to the number and frequency of such events ^4–7^. Further, World Cup games may carry lower risk for many infectious diseases than other types of mass gatherings, as they will be held in modern sporting stadiums that routinely host events with upwards of 50,000 attendees (stadium capacities range from 46,000 to 93,000 ^8^). This stands in contrast to, for instance, summer music festivals that often take place in ad hoc outdoor venues without permanent water and sanitation facilities (e.g., porta-a-potties).

Still, there are aspects of the 2026 FIFA World Cup that reasonably raise concerns for infectious disease transmission in ways that typical summer gatherings and routine sporting events do not. Most notably, the World Cup is one of the most international of sporting events, with national teams having large, committed fan bases who are willing to travel long distances to watch their teams play. The teams represented span the world, and many countries that are home to diseases absent from North America have qualified for the tournament. The tournament will also lead to watch parties and post-game celebrations in homes, hotels, restaurants and bars where transmission might occur. This congregating is happening against the backdrop of recent outbreaks of measles ^9^ and arboviruses ^10^ in the United States, and multiple global disease crises, all of which raise concern that the traditional firewalls against infectious disease outbreaks have broken down.

To help focus thinking around infectious threats at the 2026 FIFA World Cup, we here attempt to provide an empirical assessment of what diseases actually pose increased risk to US host cities around the tournament based on analysis of travel patterns and application of standardized criteria.

## Methods

We brought together teams from across Insight Net ^11^, a multi-institution network devoted to infectious disease forecasting and analytics, who worked closely with the US Centers for Disease Control and Prevention to assess the risk posed by 77 pathogens across three dimensions: excess importation risk, outbreak risk, and impact. This initial risk set of 77 pathogens is based on an initial list of infectious health threats from the Council of State and Territorial Epidemiologists (CSTE) (final public version at ^12^).

### Excess Importations

We determined baseline travel using the GLEAM-EPIRisk ^13,14^ metapopulation framework. Travel data was based on scheduled seat flight data aggregated to catchment areas of major transportation hubs level (approximately 3,200 geographic units worldwide) for summer 2026 sourced from OAG (Official Airline Guide), reflecting typical June travel patterns. We generated excess travel scenarios for a 10%, 20% and 35% increase in travel from countries home to World Cup teams, with appropriate allocations distributed across US host cities. The 20% increase scenario was considered the most plausible for the purposes of screening (see supplemental material for details). Where available, public pathogen surveillance data were used to calculate in-country infection prevalences. If timely surveillance data were unavailable, historic seasonal incidence rates per country or region level were used to compute importations. When other options were unavailable, limiting cases or hypothetical scenarios were determined on a per-pathogen basis. All the final surveillance data were input into the GLEAM mobility framework to compute pathogen-specific monthly importation risks accounting for mobility patterns, disease natural history, and excess traffic scenarios (see supplemental Table SX for importation assessment for each pathogen).

### Expected Outbreak Size

Outbreak risk assessment was based on pathogen-specific branching process simulations of epidemic spread over a 17-day period ^15^, representing the longest any one city hosts group play. Simulation parameters (the basic reproductive number, generation time, overdispersion and susceptibility) were determined based on a rapid review of the literature. For pathogens deemed likely to transmit more efficiently at World Cup-related events, increases in transmission due to the World Cup of 1.2x were applied (regardless of mode of transmission) based on analysis of COVID-19 incidence during the 2022 FIFA World Cup in Qatar (see supplement for details) ^16^. Expected outbreak sizes resulting from a single introduction were based on the mean of 1 million simulations. Full methods, supporting analyses and parameter estimates are available in the supplemental material.

### Impact Analysis

To capture the impact of disease severity and associated public health response for a pathogen, we translated all impacts to US dollar costs using established approaches and combined these into a single pathogen impact estimate. Where available, we included three different costs in our calculation: (i) medical costs (e.g., outpatient treatment, hospitalization), (ii) productivity losses (e.g., workforce loss from morbidity and mortality), and (iii) public health response costs (e.g., outbreak investigation, contact tracing). Costs were extracted from peer review literature where possible, and additional media such as news articles if needed, and when a combined cost estimate was available we used that. For pathogens where no public health responses costs could be found we looked for costs of analogous pathogens, and if unavailable, assumed response costs to be negligible. In cases where productivity losses were not reported in the literature, we calculated the cost by multiplying the DALY per case (extracted from existing research or calculated from the Global Burden of Disease Study online tool^17^) and the GDP per capita for the U.S., estimated at $94,430 for 2026^18^. To standardize the assessments, costs for each pathogen were computed in US dollars using historic currency conversion rates and accounting for inflation where appropriate (see supplement for full methods).

### Assessment Protocol

To standardize assessment and streamline the process, we developed and applied a step-wise protocol for screening each pathogen for inclusion in a list of “priority” pathogens of elevated concern around the tournament (Figure 1, supplementary methods). Briefly, this protocol first divided pathogens into those currently present in the United States (endemic pathogens) and those that are absent,except for importations (non-endemic pathogens).

**Figure 1.**
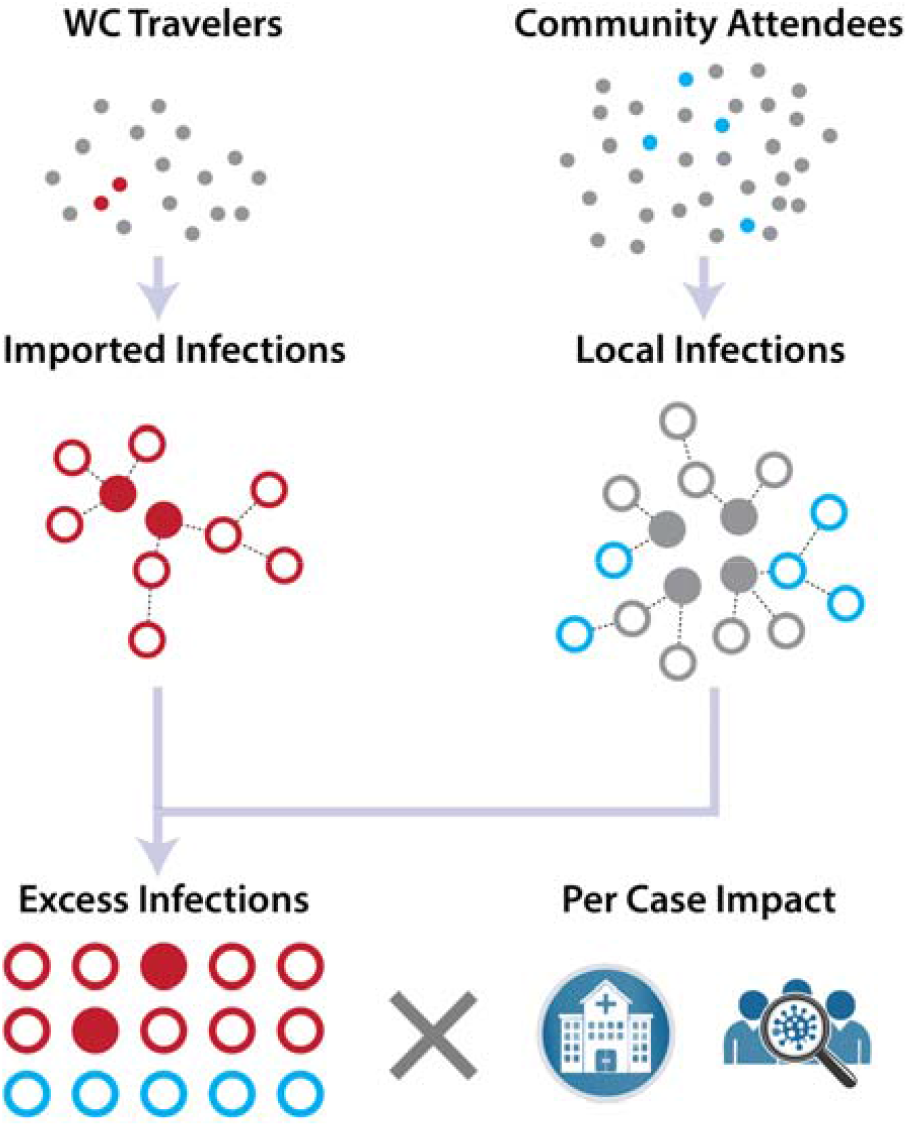
*Top row:* The expected number of excess importations of infection (red dots) and infected individuals from the local community (blue dots) present at World Cup events are determined by applying country specific incidence data to the appropriate populations. *Middle row:* Imported and local infections (solid dots) seed outbreaks at World Cup-related events. However, some of these infections would have occurred even if the tournament had not taken place (grey dots) and do not count as additional cases for impact analysis. *Bottom row:* Excess infections from World Cup-related activities are multiplied by estimates of per case impacts inclusive of treatment costs, costs of public health response, and the impact of reduced health (all quantified in US dollars) to determine the full expected impact.

Non-endemic pathogens were included on the priority pathogen list if there was evidence that the pathogen might have the ability to establish endemic transmission (e.g., presence of competent vectors and evidence of recent transmission) and there was a non-negligible importation risk, as endemicity of a new pathogen was considered to be an outcome with high enough impact to exceed any reasonable threshold. Pathogens deemed not to have the potential for sustained transmission in the United States were assessed based on the expected excess infections (expected imports plus the number of infections likely to occur in an outbreaks caused by imported infections, see above), and the per-infection impact, with $25,000 used as the threshold for inclusion.

Endemic pathogens were divided into pathogens for which there is evidence supporting the potential for elevated transmission during World Cup-related events and all others. Pathogens without evidence of potential transmission amplification were excluded if expected importations would not increase local prevalence by more than 5%. For all others, expected excess infections were determined based on the number of importations, the expected size of outbreaks seeded by each imported infection, and the difference between the number of additional infections expected to be caused by individuals from the community participating in World Cup-related activities minus the number they would have caused had they not participated in such activities (Figure 1, supplemental material). Excess infections were multiplied by per-case impact and compared with the threshold of $25,000 to determine inclusion.

The number of non-imported infections at World Cup-related activities who might transmit was calculated by multiplying national prevalence or incidence rates by the size of the metro areas of host cities by the percentage of the overall population expressing interest in attending World Cup games ^19^ (see Supplemental Material).

## Results

In the primary scenario of a 20% increase in travel flow from participating countries during the tournament window, the implied number of excess trips ranges from a few hundred for the Democratic Republic of Congo (likely fewer in light of the recently announced travel ban^20^) to well above 100,000 for the United Kingdom. For the 10 pathogens where surveillance data permitted a formal importation analysis (dengue, chikungunya, yellow fever, measles, pertussis, mumps, rubella, mpox clade I, cholera and typhoid), the expected importations across all venues and the full tournament period ranged from <0.01 for yellow fever to 4.7 for chikungunya. Importation risk from hypothetical Marburg (50 infections in Ghana) and Ebola (1,000 infections in DR Congo, no travel ban) were even lower at 0.002 and 0.0007 expected importations, respectively. For some pathogens where ad-hoc rate-based analyses were used, many projected much higher importation rates, particularly Hepatitis B (18,386 chronically infected individuals projected to visit) and seasonal respiratory viruses (e.g., 946 influenza importations).

In systematic screening, 43 of 52 endemic pathogens were excluded because there was no evidence to support elevated transmission risk during World Cup-related activities and expected importations would not increase local prevalence by more than 5% (Figure 2). Among endemic pathogens with likely elevated transmission at World Cup-related activities, two (mumps and chickenpox) were excluded because the number and impact of projected excess infections did not reach our inclusion threshold.

**Figure 2.**
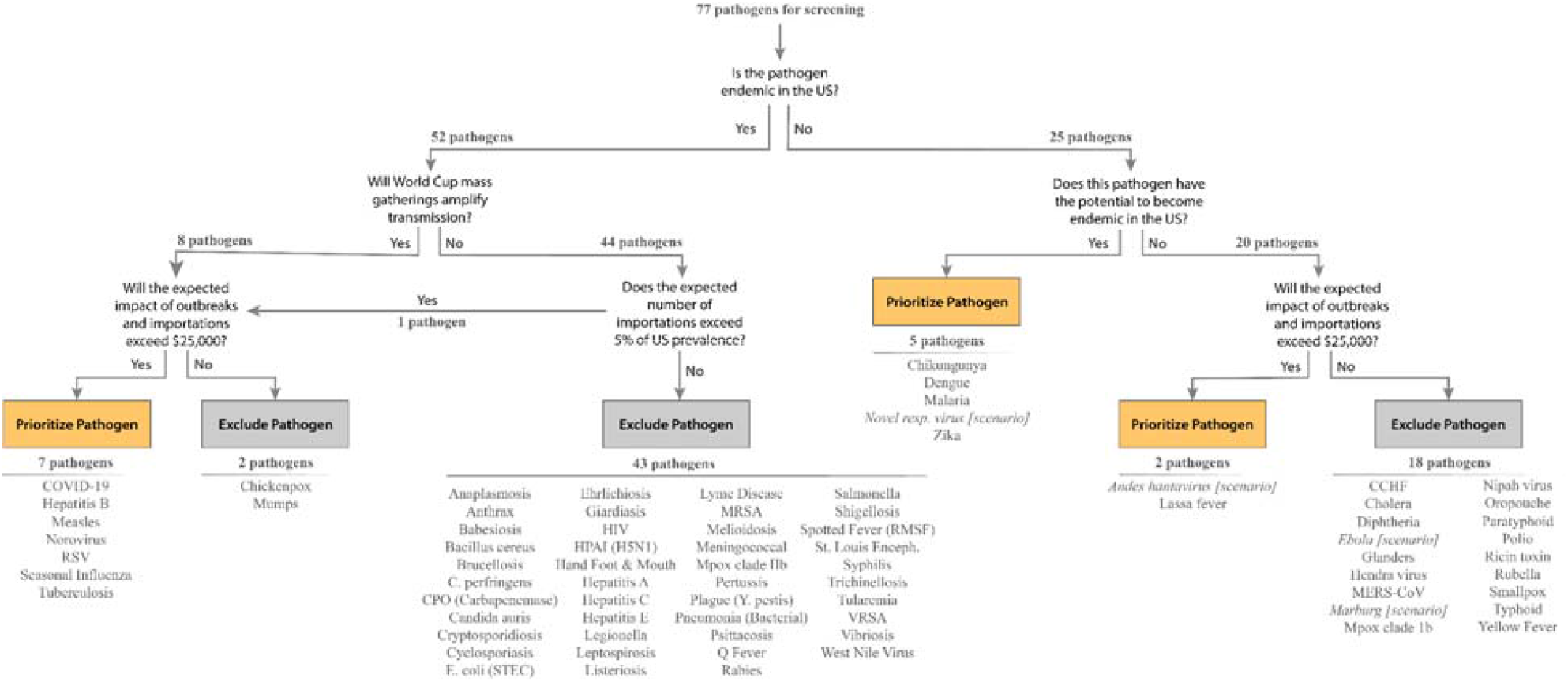
Pathogen screening process with final pathogen classifications.

Of the 25 non-endemic pathogens, 18 were excluded because the combination of expected importations, subsequent outbreak sizes, and impact did not reach our inclusion threshold. These include bioterrorism agents (e.g., smallpox), for which non-zero importation risk was considered outside of the scope of review, and high-impact pathogens (e.g., Ebola, Marburg) for which expected impact was very low despite very high per-infection impacts.

The remaining 14 priority pathogens can be grouped into five categories. First are mosquito borne pathogens with the potential for sustained transmission in some host cities. These are dengue, zika, chikungunya and malaria, and are only of elevated concern to host cities in the southern United States (e.g., Miami, Houston).

Second are the seasonal respiratory viruses, COVID-19, influenza and RSV, which may be in season in other parts of the world and may have elevated transmission at World Cup-related activities. All of these have high numbers of expected imported infections (>900) and, with the exception of RSV (which has a lower *R*_*t*_), the potential for significant excess onward transmission during the tournament. However, though absolute numbers are high (over 44,000 excess infections for COVID-19), the per-capita risk remains low (less than 60 per 10,000 among those participating in World Cup activities, and less than 5.5 per 10,000 in host communities overall).

Third are chronic infections with significantly higher prevalence in countries participating in the 2026 World Cup, compared to the United States. These are hepatitis B and tuberculosis, each with a very different expected impact. The expected 18,000+ people chronically infected with hepatitis B expected to travel for the World Cup are projected to incur no additional direct costs and only infect ∼10 additional individuals (who are unlikely to be detected during the tournament, but have a projected impact of over $25,000 per case due to long term sequelae). In contrast, projected active TB importations based on source country reactivation rates are low (15 importations) but could have significant local impact. Along with locally present infections, it is expected they would cause an additional 29 infections and have the potential to spark expensive contact tracing efforts.

Fourth are locally present pathogens with high outbreak potential and expected increased transmission risk at World Cup-related activities: measles and norovirus. Measles has a low expected number of additional infections due to the World Cup (1.1 additional infections), but is included based on per-infection impact. In contrast, norovirus has a low per-infection impact, but our analysis shows that over 6,500 additional norovirus infections may be expected (∼9 infections per 10,000 attendees).

The final group consists of high-consequence infectious pathogens with a low probability of being present at the tournament, but very high impact if they are. This group includes Lassa fever, for which we project a ∼0.3% increased probability (i.e., and expected 0.003 importations) of importation based on previous trends, but has a high enough cost if imported to warrant inclusion (assuming a response similar to that warranted by Ebola). We also included two pathogens for which we performed a scenario analysis based on high underlying uncertainty, Andes lineage hantavirus and a hypothetical novel respiratory pathogen (assumed to be pandemic influenza-like). In both scenarios the number of infections we would expect from a single introduction during the World Cup is relatively small, 2 for hantavirus and 18 for a novel respiratory pathogen, but the overall projected impact is large.

The full list of excluded and priority pathogens with referenced justifications is provided in supplementary table 1.

## Discussion

The 2026 FIFA World Cup will bring travelers from around the world to the United States, Canada and Mexico, and wherever people go, infectious disease goes as well. However, after subjecting 77 pathogens to careful screening based on importation potential, expected onward transmission and impact, fewer than 20% were identified as having meaningfully potential health impact. These priority pathogens fall into five clear categories, mosquito borne diseases only of concern to a small number of host cities, seasonal respiratory viruses that might experience off-season amplification during the tournament, chronic infections with low prevalence in the United States, highly contagious diseases already present in the country, and high-consequence pathogens with a very low probability of being introduced. Though notable, in an already highly connected world ^21^, the increased additional infectious disease risk due to the World Cup is small even for the pathogens on this list; and while public health and clinical vigilance is always warranted, the tournament is unlikely to substantively change the landscape of infectious disease risk in the United States.

Many readers may be surprised by some of the pathogens that did not make this list, in particular, most sexually transmitted infections (STIs) and gastrointestinal illnesses (GI) associated with food borne outbreaks. Though media reports associating STIs^22^ and high condom demand^23^ with sporting events have attracted attention, the systematic studies that have been performed of STI transmission around major sporting tournaments, including the 2010 World Cup in South Africa and the 2012 Olympics in London, show no evidence of increased transmission ^24–26^. Likewise, while GI illness is frequently associated with mass gathering events, the majority of reported outbreaks are associated with gatherings where sanitation facilities and access to clean water may be limited. In contrast, World Cup venues have modern (and usually inspected) sanitation and food preparation facilities used to dealing with large crowds. Further, there is no indication that attendees to the World Cup will be staying in informal settlements or other conditions associated with elevated transmission of GI illness (and reliance on such accommodations seems highly unlikely, given the cost of attendance).

The exclusion of Ebola and Marburg, particularly given recent events, might be surprising given that Lassa fever made the list. However, Lassa fever is far more common than these other diseases, with over 100,000 infections per year ^26^, and we presume an importation would warrant a similar reaction as to its more deadly cousins.

This work is subject to some notable limitations. First, we do not take into account differences between who travels and who is likely to have diseases when calculating importation rates. Particularly for conditions associated with low socio-economic status and co-infection, like active tuberculosis infection, incidence in those traveling for the World Cup will likely be far lower than projected based on overall population prevalence in countries of origin (this problem is particularly acute when we use rates based on region or development index). Likewise, the age distribution of infections might reduce true importation rates. Second, cost calculations are, in general, based on broad population analyses. These costs may be centered in populations unlikely to attend World Cup-related activities (e.g., very young or very old people) and might reflect costs that seem irrelevant to calculation of acute risk (e.g., sequelae that might only present decades later). Third, fully standardized methods were not possible across pathogens due to varying pathogen specific data limitations, negatively impacting comparability. Fourth, we do not explicitly account for US travel or heterogeneities in pathogen presence throughout the country, which may have meaningful effects on local disease risk. Finally, the speed and scope of the assessment meant that use of formal systematic review methods and other highly rigorous techniques was not possible in the assessments. All of these limitations can be reduced in future efforts by development of more refined tools, techniques and data sources for risk assessment.

The 2026 FIFA World Cup will bring people from around the world to host cities in the United States, Canada and Mexico. In light of the tournament, clinicians and public health officials should broaden their diagnostic criteria to account for the international nature of the event and maintain effective prevention efforts (e.g., condom distribution). Though such prudence is warranted, it is important to temper caution with a realistic assessment of what infectious disease risk the tournament might actually pose. A systematic approach, such as the one taken here, can provide a more focused list of pathogens than we might generate through more ad-hoc approaches, and forces us to examine our assumptions about risk. It is our hope that refined versions of our tools and protocol can help in assessing infectious disease at future international gatherings, whether they be high profile events such as the 2028 Olympic Games, or smaller events of interest to local physicians and health departments.

## Data Availability

Code for the analyses can be found at https://github.com/ACCIDDA/wc-risk-assessment

## Notes

### Competing Interest Statement

The authors have declared no competing interest.

## Bibliography

1. Bevenour G. World Cup Set to Kick Off US Inbound Travel Rebound. Oxford Economics. November 13, 2025. Accessed May 28, 2026. https://www.oxfordeconomics.com/resource/world-cup-set-to-kick-off-us-inbound-travel-rebound/

2. Islam MS, Chughtai AA, Wood JG, Sawleshwarkar S, Muscatello DJ, Seale H. Hantavirus outbreak on a cruise ship in the South Atlantic. Lancet. Published online May 15, 2026. doi:10.1016/S0140-6736(26)00934-7

3. Ebola. Accessed May 26, 2026. https://www.who.int/emergencies/situations/ebola-outbreak---drc-2026

4. Yan X, Fang Y, Li Y, Jia Z, Zhang B. Risks, epidemics, and prevention measures of infectious diseases in major sports events: Scoping review. JMIR Public Health Surveill. 2022;8(12):e40042. doi:10.2196/40042

5. Gallien Y, Fournet N, Delamare H, Haroutunian L, Tarantola A. Epidemiological surveillance and infectious disease outbreaks during mass international summertime sports gatherings: A narrative review. Infect Dis Now. 2024;54(4S):104889. doi:10.1016/j.idnow.2024.104889

6. Schenkel K, Williams C, Eckmanns T, et al. Enhanced surveillance of infectious diseases : the 2006 FIFA World Cup experience, Germany. Euro Surveill. 2006;11(12):234–238. doi:10.2807/esm.11.12.00670-en

7. Alhussaini NWZ, Elshaikh UAM, Hamad NA, Nazzal MA, Abuzayed M, Al-Jayyousi GF. A scoping review of the risk factors and strategies followed for the prevention of COVID-19 and other infectious diseases during sports mass gatherings: Recommendations for future FIFA World Cups. Front Public Health. 2022;10:1078834. doi:10.3389/fpubh.2022.1078834

8. 2026 FIFA World Cup Stadiums. US Soccer Players. Accessed May 26, 2026. https://ussoccerplayers.com/soccer-fact-sheets/2026-fifa-world-cup-stadiums

9. CDC. Measles Cases and Outbreaks. Measles (Rubeola). May 22, 2026. Accessed May 26, 2026. https://www.cdc.gov/measles/data-research/index.html

10. Kiplagat SJ, Rodriguez DM, Rivera A, Paz-Bailey G, Wong JM, Adams LE. Increase in travel-associated and locally acquired dengue cases - United States, 2024. MMWR Morb Mortal Wkly Rep. 2026;75(18):227–233. doi:10.15585/mmwr.mm7518a1

11. Insight Net – National Outbreak Analytics & Disease Modeling Network. Accessed May 26, 2026. https://insightnet.us/

12. Applied Epidemiology Recommendations for Mass Gatherings Preparedness and Response: World Cup 2026. Accessed May 27, 2026. https://cdn.ymaws.com/www.cste.org/resource/resmgr/publications/CSTE_Applied_Epidemiology_Re.pdf

13. Balcan D, Gonçalves B, Hu H, Ramasco JJ, Colizza V, Vespignani A. Modeling the spatial spread of infectious diseases: the GLobal Epidemic and Mobility computational model. J Comput Sci. 2010;1(3):132–145. doi:10.1016/j.jocs.2010.07.002

14. Davis JT, Chinazzi M, Perra N, et al. Cryptic transmission of SARS-CoV-2 and the first COVID-19 wave. Nature. 2021;600(7887):127–132. doi:10.1038/s41586-021-04130-w

15. Diekmann O, Heesterbeek JAP. Mathematical Epidemiology of Infectious Diseases. John Wiley & Sons; 2000.

16. Dergaa I, Ben Saad H, Zmijewski P, et al. Large-scale sporting events during the COVID-19 pandemic: insights from the FIFA World Cup 2022 in Qatar with an analysis of patterns of COVID-19 metrics. Biol Sport. 2023;40(4):1249–1258. doi:10.5114/biolsport.2023.131109

17. GBD Results. Institute for Health Metrics and Evaluation. Accessed May 28, 2026. http://vizhub.healthdata.org/gbd-results/

18. [No title]. Accessed May 28, 2026. https://www.imf.org/external/datamapper/NGDPDPC@WEO/USA/DEU/CAN

19. Do you plan to buy tickets to attend a live match of the FIFA World Cup 2026, hosted in the summer by the U.S., Canada, and Mexico? Accessed May 27, 2026. https://yougov.com/en-us/daily-results/20260415-a34db-1

20. U.S. Embassy Kinshasa. Update on Travel from the DRC to the United States. U.S. Embassy in the Democratic Republic of the Congo. May 21, 2026. Accessed May 27, 2026. https://cd.usembassy.gov/update-on-travel-from-the-drc-to-the-united-states/

21. Recent Developments in International Tourism to the United States. Accessed May 27, 2026. https://www.congress.gov/crs-product/IN12589

22. Analyte Health, Inc. STDcheck.com Data Reveals Genital Herpes Rate Rose Average Of 56% In Super Bowl-Hosted Cities Last 3 Years. Cision PR Newswire. May 25, 2022. Accessed May 27, 2026. https://www.prnewswire.com/news-releases/stdcheckcom-data-reveals-genital-herpes-rate-rose-average-of-56-in-super-bowl-hosted-cities-last-3-years-301555101.html

23. Lee BY. 2026 Winter Olympics Village Ran Out Of Condoms After Just 3 Days. Forbes. February 13, 2026. Accessed May 27, 2026. https://www.forbes.com/sites/brucelee/2026/02/13/2026-winter-olympics-village-ran-out-of-condoms-after-just-three-days/

24. Delva W, Richter M, De Koker P, Chersich M, Temmerman M. Sex work during the 2010 FIFA World Cup: results from a three-wave cross-sectional survey. PLoS One. 2011;6(12):e28363. doi:10.1371/journal.pone.0028363

25. Sile B, Mohammed H, Crook P, et al. Epidemiology of sexually transmitted infections in visitors for the London 2012 Olympic Games: A review of attendees at sexual health services: A review of attendees at sexual health services. Sex Transm Dis. 2015;42(12):710–716. doi:10.1097/OLQ.0000000000000370

26. Hall V, Charlett A, Hughes G, et al. Olympics and Paralympics 2012 mass gathering in London: time-series analysis shows no increase in attendances at sexual health clinics. Sex Transm Infect. 2015;91(8):592–597. doi:10.1136/sextrans-2014-051826

