## Supplemental materials for "Why epidemic risk at the 2026 World Cup may not be what you think"

Code for the analyses described herein can be found at https://github.com/ACCIDDA/wc-risk-assessment.

#### Section S1: Excess Importation Risk

| Pathogen | Active Cases Computation (monthly) | Reference |
|---|---|---|
| Mpox Clade I | (cases in last 6 weeks / 42) × 30; countries with Community Transmission only | [1] |
| Dengue | Average of last 6 months available; countries with ≥ 5 months of data | [2] |
| Measles | Average of last 6 months available; countries with ≥ 4 months of data | [3] |
| Cholera | Incidence rate × population / 100,000) / 3; ceiling of ECDC band | [4] |
| Pertussis | Annual cases (2025) / 12 | [5] |
| Diptheria | Annual cases (2025) / 12 | [6] |
| Yellow Fever | Annual cases (2025) / 12 | [7] |
| Typhoid | Annual cases (2025) / 12 | [8] |
| Rubella | Annual cases (2025) / 12 | [9] |
| Mumps | Annual cases (2025) / 12 | [10] |
| Chikungunya | Incidence rate × population / 100,000) / 3; ceiling of ECDC band | [11] |
| Ebola | Scenario only (anchored to reported Bundibugyo ebolavirus activity) | |
| Marburg | Scenario only (hypothetical case counts) | |

A modeling framework was developed to estimate the risk of disease importation into the United States during the 2026 FIFA World Cup (WC2026), held across eleven US host cities. For each participating country, the framework computed the probability that infectious travelers reached the United States under baseline travel conditions and under scenarios of elevated travel demand driven by WC2026 fan mobility. Results were produced at two geographic scales: the United States as a whole and the individual US host cities.

Active case estimates were derived from publicly available surveillance data accessed on May 8, 2026. For pathogens with active surveillance signals, monthly active cases were estimated as the average monthly count over the last months of available data with reported activity. When fewer than six months of historical data were available, the most recent month with reported activity was used as a fallback. Dengue case counts were obtained from the WHO Dengue Global Dashboard [2]. Chikungunya case counts were obtained from the ECDC Chikungunya Monthly Situation Report [11]. Yellow fever case counts were obtained from the WHO Immunization Data Portal [7]; a reporting threshold of 0.001 expected imports per month was applied at the country level, and 24 source countries fell below the threshold and were excluded from the reported results, with a combined cumulative contribution below 0.01 expected imports per month. Measles, pertussis, mumps, rubella, and typhoid case counts were obtained from the WHO Immunization Data Portal [3] [5] [10] [9] [8]. Mpox Clade I case counts were obtained from the WHO Mpox Global Dashboard [1], and active cases were estimated only for countries with confirmed community transmission. Cholera case counts were obtained from the ECDC Cholera Monthly Situation Report [4].

Ebola and Marburg were treated as hypothetical scenarios for risk planning. Marburg was modeled as a fully hypothetical outbreak, with no confirmed active activity at the data access date. Case counts were set manually to represent a plausible outbreak size. Ebola was anchored to reported Bundibugyo ebolavirus activity in the Democratic Republic of the Congo. A scenario of 1,000 active cases was specified as a hypothetical upper bound on the current situation. Both scenarios were intended for risk planning rather than as forecasts and did not incorporate ground-level outbreak detail.

The mobility component of the framework was based on GLEAM-EpiRisk [12] [13], a metapopulation model that combined international flight data and commuting networks to estimate the probability that an infectious individual in a source country reached a destination. Air travel was represented through origin-destination passenger data from OAG (Official Airline Guide), aggregated to catchment areas of major transportation hubs, defining approximately 3,200 geographic units worldwide. Because the data captured full passenger itineraries from origin to final destination, connecting flights were implicitly included, and indirect routing through intermediate hubs was accounted for. Local mobility flows were derived from a gravity model calibrated on population and distance. The baseline scenario used OAG summer 2026 origin-destination data without any WC2026 perturbation and reflected typical June travel patterns between each participating country and the United States.

WC2026 was expected to generate substantial increases in international travel to the United States. Four scenarios were modeled: a baseline with no excess travel above OAG flows, and three perturbed scenarios in which total travel from the source country to the United States increased by 10%, 20%, and 35%, respectively. The excess volume in each scenario was defined as a fraction of the total baseline flow from the source country to the United States. For example, in a +10% scenario for a country with 50,000 annual travelers to the United States, the excess volume corresponded to 5,000 additional travelers.

At the country level, the full excess volume was added to the source country's US-bound link in the flight network, increasing the overall importation probability proportionally. At the city level, the excess was routed to the US host cities where the country's team played group-stage matches, with cities hosting more matches receiving a proportionally larger share. The departure distribution across source basins followed each basin's existing share of US-bound traffic. For cities with no existing direct air connection, the excess was distributed proportionally to each basin's total US-bound flow, creating new effective origin-destination pathways. This approach reflected the assumption that WC2026 fans traveled to the specific cities where their national team played and that their departure patterns mirrored existing connectivity from the source country. Only group-stage match cities were used for excess traffic redistribution. Knockout-stage venues were not included because the schedule was not deterministic at the time of scenario construction.

For each pathogen and scenario, the framework computed the full probability distribution of imported cases P(K = k) into the United States, from which several quantities were derived: the expected number of imported infectious cases per month, E[K]; the probability that no cases reached the United States from the source country in a given month, P(K = 0); the probability that at least one case reached the United States in a given month, P(K 1); and the excess expected imports per month at +10%, +20%, and +35%, defined as the difference between expected imports under a WC-concentrated scenario and the baseline. These quantities were computed using the GLEAM-EPIRisk mobility framework, which integrated the number of active cases in the source country, the per-individual daily probability of flying to the United States derived from OAG origin-destination data, the length of the traveling window of exposed carriers, and global flight and commuting networks.

For each pathogen and scenario, a relative risk score was also computed for every indexed US destination basin (approximately 400 cities and metropolitan areas). The relative risk represented the probability that, conditional on at least one case reaching the United States, the case arrived in a given city. Relative risk values were normalized to sum to one across all US destinations and were reported for all indexed cities, including non-host cities, which could emerge as relevant importation destinations through airline routing and connectivity. The use of relative risk avoided overstating the precision of the model and focused attention on the comparative ranking of cities. Several assumptions and limitations of the presented framework should be noted. The model captured flight-based mobility, including indirect routing through non-US hubs, since it operated on origin-destination passenger data. It did not account for domestic overland travel within source countries or after arrival in the United States. Active case counts reflected the best available surveillance data as of May 8, 2026, and may have underestimated the true burden due to underreporting. The incubation period was used as a proxy for the traveling window of exposed carriers, and cases that became symptomatic before departure were assumed not to travel. Excess traffic scenarios were approximations, and actual WC2026 travel patterns will depend on team performance, ticket availability, and booking behavior. Knockout-stage cities were excluded from excess traffic redistribution because the schedule was not deterministic at the time of scenario construction.

For a subset of pathogens we make ad hoc importation estimates based on alternate data sources combined with per country excess traveler estimates derived from the above approach. In particular, we take the following approaches: - Apply country or region-specific estimates to World Cup excess traveler counts as follows:

$$\textit{Expected importations} = \sum_i \textit{Excess travelers from country } i \times \textit{Incidence in country } i \times 39/365$$

- Apply historical importation rates to World Cup excess traveler counts

$$\textit{Expected importations} = \sum_i \textit{Excess travelers from country } i \times \textit{Historical importation rate to US} \times 39/365 \times 0.2$$

- Use the importation estimates for an analogous pathogen, scaled by the difference in global occurrence

$$\textit{Expected importations for pathogen } j = \textit{Expected importations for pathogen } k \times \frac{\textit{Global incidence of pathogen } j}{\textit{Global incidence of pathogen } k}$$

Details on how the above approached were applied for each pathogen can be found in Supplementary Table 1.

#### Section S2: Outbreak Risk

##### (A) Excess risk matric calcualtion

We calculate the expected number of excess infections due to the World Cup (WC), $WCI_i$, as follows:

$$\begin{aligned} WCI_i &= (\textit{Excess importations} \times \textit{WC per infection outbreak size}) \\ &+ [\textit{Local presence} \times (\textit{WC per infection outbreak size} - \textit{Baseline per infection outbreak size})] \end{aligned}$$

We multiply this value by the expected impact per infection to obtain the excess risk from the World Cup for pathogen $i$, $WCR_i$:

$$WCR_i = WCI_i \times \textit{Impact per infection}$$

Here, *Excess importations* is the number of international travelers infected with pathogen $i$ coming to the World Cup (see section S2.A). *Local presence* is the number of infections expected to be present at World Cup related activities from sources other than people traveling for the World Cup (see section S2.C). WC per infection outbreak size is the average size of an outbreak resulting from the presence of a single infection (inclusive of that infection) at World Cup related activities. *Baseline per infection outbreak size* is the average size of an outbreak resulting from the presence of a single infection (inclusive of that infection) if the World Cup were not occurring. Per infection outbreak sizes are calculated using a branching process model (see S2.B). *Impact per infection* is the summation of the medical costs, public health response costs and productivity losses associated with a symptomatic case of the pathogen. In some cases, it was also necessary to use the cost associated with infection (i.e. including asymptomatic infections) instead (see section S2.E).

##### (B) Branching process model

The overall outbreak risk can be quantified by the expected number of additional infections resulting from an imported infection. A baseline, "back of the envelope" assessment of outbreak risk associated with the games themselves can be calculated based on simple branching process simulation. These simulations rely on two inputs: the offspring distribution (i.e. how many infections each infection produces) and the number of generations of transmission.

To determine the number of generations, we divide the total duration of opportunity for spread during the worst-case continuous mass-gathering context, T, by the average generation time for a pathogen, $g$, and then round to the nearest integer. We sample $T = [1, 17]$ days based on the assumption that infectious contributors arrive uniformly over the longest duration from first to final game during group play in any US city (15 days in NY/NJ) plus two additional days.

For the distribution of offspring in a fully susceptible population, $E$, we assume a negative binomial parameterization:

$$E \sim \text{NegBin}\left(\mu = R_0, \text{size} = k\right)$$

where this parametrization corresponds to the R `{stats} nbinom(mu, size)` formulation, $R_0$ is the basic reproductive number for a particular context (World Cup or baseline), and $k$ is the offspring overdispersion (i.e. low $k$ = high variability in offspring, high $k$ = low variability in offspring).

Susceptible individuals become infected by exposure, which corresponds to binomial thinning of $E$ by the susceptible fraction $S$. The combination of these processes, infections ($I$), remains a negative binomial deviate with modified parametrization:

$$I \sim \text{NegBin}\left(\mu = S \times R_0, \text{size} = k\right)$$

We assume, pessimistically, that these chains of transmission do not interfere (i.e. no susceptible depletion). Therefore, each subsequent generation of infections looks like the conditional sum of independent, identically distributed draws from the same negative binomial distribution, which again is negative binomially distributed with modified parameters. If generation n has In individuals, then the next generation is distributed as:

$$I_{n+1} \sim \text{NegBin}\left(\mu = S \times R_0 \times I_n, \text{size} = k * I_n\right)$$

To create an outbreak size sample for an index infection, we start with $I_0 = 1$, iteratively draw out to the sampled generation limit using the above distributions, and sum the generations. To build up the outbreak distribution, we sample 1 million times. Per the protocol requirements, we compute the distribution for both baseline and mass-gathering R0 values, but assume no other parameter changes.

We obtained estimates for $R0$, $S$, $k$, and $g$ per the parameter review protocol (section S2.D).

**(C) Local presence**

There are two pools of infected individuals that we might be interested in when assessing World Cup risk. The first is the overall number of infected individuals in the community and the second is the number of infected individuals from the local community who participate in World Cup related activities. For simplicity, we assume that prevalence among those participating in World Cup related activities is the same as the prevalence in the overall community.

$$\textit{Number of infected individuals in community} = \textit{Prevalence} \times \textit{Community size}$$

Community size was assumed to be 84.6 million based on the combined populations of the metro-areas for US host cities [CITE]. While we know of no good estimate of the proportion of people in the local community who will attend World Cup-related events, recent surveys indicate that 8% of people are planning to attend local matches [CITE].

$$\textit{Number of infected individuals at WC related activities} = \\ \textit{Prevalence} \times \textit{Community size} \times 0.08$$

##### (D) Rapid parameter review protocol

Estimates of the reproduction number, $R_0$, overdispersion parameter, $k$, and generation time, $g$, were obtained through a literature search conducted using the Google Scholar database. Alternative spellings of the pathogen name are used when applicable. We limit the search to articles in English. No limits are provided for a date range. Rather than a comprehensive literature review, we employ a structured approach where we search until we find parameter estimates that meet pre-defined stopping criteria, as described in the following subsection. The key terms searched for each pathogen are compiled in the table below. For pathogens where humans are dead-end hosts, the Control of Communicable Diseases Manual was consulted as the primary source.

| Parameter | Search terms |
|---|---|
| Reproduction number ($R_0$) | "[Pathogen name] AND ("reproduction number" OR "reproductive number" OR R0)" |
| Overdispersion | "[Pathogen name] AND ("over dispersion" OR "over-dispersion" OR overdispersion OR "overdispersion parameter")" - [NOTE: additional terms of "secondary transmission" and "superspread" were added when needed to further filter papers] |
| Generation time | "[Pathogen name] AND ("generation time" OR "generation interval" OR "transmission interval" OR "serial interval")" |

Sources for all parameter estimates were stored in a Zotero folder. For each source, we document the pathogen name, disease category, parameter of interest and estimate, type of estimate, uncertainty estimates, type of uncertainty, location of study, and pathogen variant (when applicable) along with a link to the reference.

Stopping criteria for the parameter review were applied as follows:

- The first ~50 articles returned from the queries in Google Scholar were searched to find a systematic review. If a review was available, we used our expert judgement to determine whether it should be included based on the timeliness, comprehensiveness and reliability of the review. If deemed appropriate, the pooled mean/median estimates and their uncertainty estimates were extracted.
- If systematic reviews were unavailable, we searched for articles reporting individual estimates or articles cited by those articles and selected them based on the same criteria of expert judgement. If published estimates referred to a canonical source, the canonical source article was used. Each estimate and its reported uncertainty (when available) were extracted.

- In some instances, parameter estimate(s) could not be determined for a particular pathogen following the aforementioned literature search criteria and alternate approaches to obtain an estimate were used. If an analogous pathogen could be identified for which an estimate for the parameter of interest could be determined using the same search criteria, this estimate was used as a proxy for our pathogen of interest. For example, the R0 of paratyphoid fever was assumed to be the same as that of typhoid and the k of pertussis was used as a proxy for chickenpox, diphtheria, mumps and rubella. In certain instances a direct parameter estimate was not available, however other information that could be used to infer the parameter estimate was available. Below we detail these ad hoc analyses for each parameter.

**Overdispersion** If no direct estimates of $k$ were available but there was published data on the offspring distribution from an outbreak or, in the case of point source outbreaks, the distribution of outbreak sizes, we derived a maximum likelihood estimate of $k$ based on the assumption that reported data on the offspring or outbreak size distribution, $x$, follows a negative binomial distribution with mean $R_0$. Specifically, the MLE is the value of $k$ which maximizes $\sum_i \log[f(x_i|R_0, k)]$ where $f(x|\mu, k)$ is the probability mass function of the negative binomial distribution.

In the absence of outbreak size data, we searched the literature for a measure of superspreading in the context of the Pareto rule where the general assumption is that 20% of infections are responsible for 80% of transmission. The following key terms are used: "[pathogen name]" AND and ("20/80" OR "80/20"), "[pathogen name] AND "pareto rule", "[pathogen name]" AND "superspreader" ", "[pathogen name]" AND "transmission" AND "distribution".

We derived an estimate of $k$ from the Pareto rule (in the form of $y$ percent of infections are responsible for $z$ percent of transmission) as follows. For a given $R_0$ and $k$ value, we simulate 10,000 values $(x_1, x_2, ..., x_10000)$ for the number of secondary infections produced by an infectious individual from the negative binomial function. We then calculate $z_k$, the percentage of secondary infection caused by the top $y$ percent of spreaders as follows:

$$z_k = \frac{\sum_{i=1}^{10000} x_i \mathbf{I}(x_i \geq q_{1-z})}{\sum_{i=1}^{10000} x_i}$$

Where $q_{1-z}$ is the $(1-z)$th quantile of $x_1, x_2, ..., x_{10000}$. We then compute $z_k$ for a range of k values. We initially search over $k \in (0, 1]$ if $z_1 < z$ and continue to iteratively double the search range until the maximum $z_k < z$. Our estimated value of $k$ is that which minimizes $|z_k - z|$.

**Generation time** Generation time and/or serial interval estimates are not available for most enteric pathogens. Instead, we searched for estimates of the incubation period and used them as a proxy for generation time. Sources of disease statistics, such as the Centers for Disease Control and Prevention (CDC), World Health Organization (WHO) and any relevant local or national government agency websites were also used to determine these values.

For each combination of pathogen and parameter, we followed the approach described below to obtain a measure of central tendency for each parameter estimate from the estimates provided by the articles selected through our literature search. Note that any articles which only reported a range for the parameter estimate were not included if articles which reported a point estimate were available. To implement the criteria described below, we also made the assumption that median estimates are approximately equivalent to the mean.

- If a pooled estimate was obtained from a systematic review, or if there was only one article that met our criteria and reported a point estimate, we used this value as the central estimate for the parameter.
- If multiple articles were selected and all reported a 95% confidence interval (CI), 95% credible interval (CrI), 95% high posterior density interval (HPDI) or standard error (SE), the SEs were reconstructed from the intervals under the assumption of a normal distribution and a meta-analysis approach using the `rm()` function from the `{metafor}` package in R was implemented to obtain a central estimate (Method 1).

- If the articles selected reported some combination of a range, IQR, CI, CrI, HPDI, or did not report any uncertainty estimates, we used the 25% trimmed mean as the central estimate (Method 2A). If fewer than 5 estimates were collectively obtained from the selected articles, we calculated the mean of means rather than a trimmed mean (Method 2B).

Table S3: Estimates of basic reproduction number ($R_0$) obtained for each pathogen following the literature search guidelines from the parameter review protocol.

| Pathogen | Estimate | Uncertainty type | Location | Reference |
|---|---|---|---|---|
| Candida auris | 0.943 (0.4–1.27) | IQR | US | [14] |
| Chickenpox | – (8–10) | range | | [15] |
| Chickenpox | 6.47 (5.62–7.55) | 95% CI | Belgium | [16] |
| Chickenpox | 3.83 (3.32–4.49) | 95% CI | England and Wales | [16] |
| Chickenpox | 4.85 (3.89–6.04) | 95% CI | Finland | [16] |
| Chickenpox | 5.46 (5.16–5.76) | 95% CI | Germany | [16] |
| Chickenpox | 5.22 (4.53–6.14) | 95% CI | Ireland | [16] |
| Chickenpox | 7.71 (6.01–10.06) | 95% CI | Israel | [16] |
| Chickenpox | 3.31 (2.82–3.83) | 95% CI | Italy | [16] |
| Chickenpox | 8.28 (6.74–10.42) | 95% CI | Luxembourg | [16] |
| Chickenpox | 16.91 (11.5–24.18) | 95% CI | Netherlands | [16] |
| Chickenpox | 5.72 (4.72–6.81) | 95% CI | Slovakia | [16] |
| Chickenpox | 3.91 (3.53–4.38) | 95% CI | Spain | [16] |
| Chickenpox | 9 | | Maryland | [17] |
| Chickenpox | 8.5 | | Massachusetts | [17] |
| Cholera | 1.44 (1.06–2.63) | range | Haiti | [18] |
| Cholera | 2.1 (0.8–7.3) | 95% CI | Peru | [19] |
| Cholera | 1.52 (1.14–1.96) | 95% CI | Zimbabwe | [20] |
| Cholera | 2 (1.41–2.8) | range | Africa | [21] |
| Cholera | 1.04 (0.85–1.55) | 95% CI | Mexico | [22] |
| COVID-19 | 5.94 (5.19–6.68) | 95% CI | | [23] |
| Diphtheria | 2.6 (1.7–4.3) | range | | [24] |
| Ebola | 1.4 | | DRC | [25] |
| Ebola | 1.8 | | DRC | [25] |
| Ebola | 1.93 (1.74–2.78) | IQR | DRC | [25] |
| Ebola | 2.7 (1.9–2.8) | 95% CI | DRC | [25] |
| Ebola | 2.98 (2.11–4.36) | 95% CrI | DRC | [25] |
| Ebola | 2.7 (2.5–4.1) | 95% CI | Uganda | [25] |
| Ebola | 0.89 | | Guinea | [25] |
| Ebola | 1 (0.77–1.35) | range | Guinea | [25] |
| Ebola | 1.26 (1.22–1.29) | 95% CI | Guinea | [25] |
| Ebola | 1.6 (1.15–2.05) | range | Guinea | [25] |
| Ebola | 1.21 (1.21–1.21) | 95% CI | Guinea | [25] |
| Ebola | 1.49 (1.48–1.5) | 95% CI | Guinea | [25] |
| Ebola | – (0.85–1.22) | range | Guinea | [25] |
| Ebola | 1.18 (1.17–1.19) | 95% CI | Guinea | [25] |
| Ebola | 1.24 (1.04–1.42) | 95% CI | Guinea | [25] |
| Ebola | – (1.2–2.02) | range | Guinea, Liberia, Nigeria, Sierra Leone | [25] |
| Ebola | 1.3 (0.36–3.37) | range | Guinea, Liberia, Sierra Leone | [25] |
| Ebola | 1.96 | | Guinea, Liberia, Sierra Leone | [25] |

*(continued on next page)*

(continued from previous page)

| Pathogen | Estimate | Uncertainty type | Location | Reference |
|---|---|---|---|---|
| Ebola | 1.35 (1.35–1.35) | 95% CI | Guinea, Liberia, Sierra Leone | [25] |
| Ebola | – (1.24–1.56) | range | Guinea, Liberia, Sierra Leone | [25] |
| Ebola | 1.8 (1.5–2) | range | Guinea, Liberia, Sierra Leone | [25] |
| Ebola | 1.4 | | Guinea, Liberia, Sierra Leone | [25] |
| Ebola | 5.92 (2.97–11.12) | 95% HPDI | Guinea, Liberia, Sierra Leone | [25] |
| Ebola | 2 | | Liberia | [25] |
| Ebola | 2.11 (1.88–2.71) | other | Liberia | [25] |
| Ebola | 1.4 (0.1–3.9) | 95% CI | Liberia | [25] |
| Ebola | – (0.9–4.6) | range | Liberia | [25] |
| Ebola | 3.02 (2.61–3.49) | 95% CI | Liberia | [25] |
| Ebola | 1.82 (1.77–1.83) | 95% CI | Liberia | [25] |
| Ebola | 1.95 | | Liberia | [25] |
| Ebola | 2.49 (2.38–2.6) | 95% CI | Liberia | [25] |
| Ebola | 1.54 | | Liberia | [25] |
| Ebola | 1.8 (1.77–1.83) | 95% CI | Liberia | [25] |
| Ebola | 2.01 | | Liberia | [25] |
| Ebola | 1.7 (1.1–2.6) | 95% CI | Liberia | [25] |
| Ebola | 2.06 (1.93–2.27) | 95% CI | Liberia | [25] |
| Ebola | 0.5 (0.2–1) | 90% CI | Africa, Europe, USA | [25] |
| Ebola | 3.2 (0.8–16.4) | 95% CrI | Nigeria | [25] |
| Ebola | 10 (3–18.5) | 95% CrI | Nigeria | [25] |
| Ebola | – (0.4–12) | range | Nigeria | [25] |
| Ebola | 9.01 | | Nigeria | [25] |
| Ebola | 2.5 | | Sierra Leone | [25] |
| Ebola | 1.9 (1.86–1.95) | 95% CI | Sierra Leone | [25] |
| Ebola | 1.3 (1.26–2.53) | range | Sierra Leone | [25] |
| Ebola | 1.7 | | Sierra Leone | [25] |
| Ebola | 2.18 (1.24–3.55) | 95% HPDI | Sierra Leone | [25] |
| Ebola | – (2.1–3.81) | range | Sierra Leone | [25] |
| Ebola | – (0.6–5.9) | range | Sierra Leone | [25] |
| Ebola | – (0.72–1.77) | range | Sierra Leone | [25] |
| Ebola | 1.26 | | Sierra Leone | [25] |
| Ebola | 1.61 (1.56–1.66) | 95% CI | Sierra Leone | [25] |
| Ebola | 1.67 (1.59–1.76) | 95% CI | Sierra Leone | [25] |
| Ebola | – (1.21–1.6) | range | Sierra Leone | [25] |
| Ebola | – (1.7–4.8) | range | Sierra Leone | [25] |
| Ebola | 1.63 (0–4) | range | Sierra Leone | [25] |
| Ebola | 2.24 (1.52–4.51) | 95% CI | Sierra Leone | [25] |
| Ebola | – (0.5–8.4) | range | Sierra Leone | [25] |
| Ebola | 2 (1.8–2.2) | other | Sierra Leone | [25] |
| Ebola | 2.39 (2.05–2.84) | 95% CrI | Sierra Leone | [25] |
| Ebola | 2.9 (1.2–6.9) | 90% CrI | Sierra Leone | [25] |
| Ebola | 0.29 (0.11–0.55) | | Sierra Leone | [25] |

(continued on next page)

(continued from previous page)

| Pathogen | Estimate | Uncertainty type | Location | Reference |
|---|---|---|---|---|
| Ebola | 0.93 (0.15–2.3) | 95% CI | Sierra Leone | [25] |
| Ebola | 1.71 (1.4–1.82) | 95% CI | Sierra Leone | [25] |
| Ebola | 2.13 | | Sierra Leone | [25] |
| Ebola | 5.15 (3.95–6.69) | 95% CI | DRC | [25] |
| Ebola | 1.08 | | DRC | [25] |
| Ebola | 7.73 | | DRC | [25] |
| Ebola | 1.83 | | DRC | [25] |
| Ebola | 1.9 | | DRC | [25] |
| Ebola | – (1.99–2.7) | range | Uganda | [25] |
| Hantavirus (Andes virus) | 2.12 | | | [26] |
| Hepatitis A | – (1.71–3.67) | range | Australia | [27] |
| Hepatitis A | 3.2 (2.85–3.54) | 95% CI | KY | [28] |
| Hepatitis A | 2 (1.8–2.2) | 95% CI | China | [29] |
| Hepatitis B | 1.53 | | | [30] |
| Hepatitis B | – (5–10) | range | | [17] |
| Hepatitis B | 1.3 | | | [31] |
| Hepatitis B | 4 | | | [31] |
| Hepatitis B | 0.53 | | | [32] |
| Hepatitis B | 2.66 | | | [32] |
| Hepatitis B | 2.406 | | China | [33] |
| Hepatitis B | 1.316 | | China | [34] |
| Hepatitis B | 3.65 | | Tonga | [35] |
| Hepatitis E | 2.25 (2.08–2.39) | | Uganda | [36] |
| Hepatitis E | 1.71 (1.32–2.47) | range | Namibia | [37] |
| Hepatitis E | 6.23 (2.8–11.4) | 95% CrI | Uganda | [38] |
| Lassa Fever | 1.74 | | | [39] |
| Lassa Fever | 1.68 | | | [39] |
| Lassa Fever | 1.84 | | | [39] |
| Lassa Fever | 1.23 (1.22–1.24) | 95% CI | | [39] |
| Lassa Fever | 1.33 (1.29–1.37) | 95% CI | | [39] |
| Lassa Fever | – (1.08–1.36) | range | | [39] |
| Marburg | 1.59 (1.53–1.66) | 95% CI | Angola | [40] |
| Measles | – (7.1–10.9) | range | | [41] |
| Measles | – (10.3–11.3) | | Germany | [41] |
| Measles | 13 | | Italy | [41] |
| Measles | 8.3 | | Denmark | [41] |
| Measles | – (12.5–18) | | UK, North America | [41] |
| Measles | – (15.4–17) | | Denmark | [41] |
| Measles | 27 | | Canada | [41] |
| Measles | – (7.1–29.3) | | Europe | [41] |
| Measles | – (21–57) | | UK | [41] |
| Measles | – (3.7–203.3) | | Africa, India | [41] |
| Measles | 4.6 | | Senegal | [41] |
| Measles | – (6.2–7.7) | | Luxembourg | [41] |
| Measles | 23 | | Netherlands | [41] |
| Measles | – (4.7–15.7) | | Niger | [41] |
| Measles | – (22.1–32.1) | | Germany | [41] |

(continued on next page)

*(continued from previous page)*

| Pathogen | Estimate | Uncertainty type | Location | Reference |
|---|---|---|---|---|
| Measles | – (10.7–18.1) | | USA | [41] |
| Measles | – (6.2–9.5) | | UK | [41] |
| Meningococcal | 1.31 (1.03–1.64) | 95% HPDI | Italy | [42] |
| MERS-CoV | 0.75 (0.54–1.09) | 95% CI | Global | [43] |
| MERS-CoV | 0.91 (0.36–1.44) | 95% CI | South Korea and Saudi Arabia | [43] |
| MERS-CoV | 2 | | Riyadh | [44] |
| MERS-CoV | 2.8 | | Riyadh | [44] |
| MERS-CoV | 3.5 | | Jeddah | [44] |
| MERS-CoV | 6.7 | | Jeddah | [44] |
| Mpox Clade Ib | 0.9 (0.71–1.09) | 95% CI | Burundi | [45] |
| Mumps | 7.1 | | Baltimore | [17] |
| Mumps | 4.3 | | North America | [17] |
| Mumps | 4.75 (2.65–5.56) | range | China | [46] |
| Nipah virus | 0.33 (0.19–0.59) | 95% CI | Bangladesh | [47] |
| Norovirus | 1.99 (1.58–2.44) | 95% CI | China | [48] |
| Norovirus | 2.75 (2.38–3.65) | IQR | US | [49] |
| Norovirus | 1.5 (1–2.2) | 95% CI | Australia | [50] |
| Norovirus | 2.62 | | Spain | [51] |
| Norovirus | 3.05 (1.8–5.02) | 95% CrI | England | [52] |
| Paratyphoid Fever[a] | 0.995 | | Taiwan | [53] |
| Pertussis | 5.5 (5.44–5.57) | 95% CrI | Finland | [54] |
| Pertussis | 5.6 (5.51–5.63) | 95% CrI | Germany | [54] |
| Pertussis | 5.9 (5.82–6.01) | 95% CrI | Italy | [54] |
| Pertussis | 5.3 (5.26–5.35) | 95% CrI | Netherlands | [54] |
| Pertussis | 5.4 (5.27–5.4) | 95% CrI | UK | [54] |
| Pertussis | – (4.1–4.9) | range | UK | [17] |
| Pertussis | 5.3 | | North America | [17] |
| RSV | 2.18 (1.49–3.24) | 95% CI | | [55] |
| RSV | 3 (2.4–3.6) | SD | | [56] |
| RSV | 8.9 | | | [57] |
| RSV | 1.7 | | | [58] |
| Rubella | – (2.7–3.8) | range | Americas | [59] |
| Rubella | – (3.7–6.7) | range | Europe | [59] |
| Rubella | 6.7 | | West Germany | [17] |
| Rubella | 6 | | UK | [17] |
| Rubella | 7.8 | | Germany | [60] |
| Rubella | 3.7 | | UK | [60] |
| Rubella | 4.2 | | Italy | [60] |
| Rubella | 4.2 | | Denmark | [60] |
| Rubella | 6.4 | | Netherlands | [60] |
| Rubella | 3.4 | | Finland | [60] |
| Rubella | 5.2 (4–6.7) | 90% CI | Africa | [61] |
| Seasonal Influenza (A/B) | 1.28 (1.19–1.37) | IQR | USA, UK, Canada, Australia, France, Norway,Brasil, Israel, Taiwan, Italy, Switzerland, Mexico | [62] |

*(continued on next page)*

*(continued from previous page)*

| **Pathogen** | **Estimate** | **Uncertainty type** | **Location** | **Reference** |
|---|---|---|---|---|
| Tuberculosis (TB) | 1.02 (1.01–1.04) | 95% CrI | | [63] |
| Tuberculosis (TB) | 2.1 (1.54–2.66) | 95% CrI | | [64] |
| Typhoid Fever | 0.995 | | Taiwan | [53] |
| Yellow Fever | 4.81 (1.35–11) | range | | [65] |

*Notes:*

[a] Value for typhoid (analogous pathogen)

Table S4: Estimates of overdispersion parameter ($k$) obtained for each pathogen following the literature search guidelines from the parameter review protocol.

| Pathogen | Estimate | Uncertainty type | Location | Reference |
|---|---|---|---|---|
| Candida auris[a] | 0.019 | | US | [66, 67] |
| Chickenpox[b] | 0.93 | | | – |
| Cholera | 4.5 (0.3–Inf) | 95% CI | Mexico | [22] |
| COVID-19 | 0.41 (0.23–0.6) | 95% CI | 12 countries including China, India, USA | [68] |
| Diphtheria[b] | 0.93 | | | – |
| Ebola | 0.31 (0.25–0.37) | 95% CI | Guinea | [25] |
| Ebola | 0.19 | | Guinea | [25] |
| Ebola | 1.6 | | Guinea | [25] |
| Ebola | 0.03 | | Guinea, Liberia, Sierra Leone | [25] |
| Ebola | 0.52 | | Guinea, Liberia, Sierra Leone | [25] |
| Ebola | 2.2 (2–2.4) | IQR | Liberia | [25] |
| Ebola | 0.82 | | Liberia | [25] |
| Ebola | 0.09 (0.03–0.2) | 90% CI | Africa, Europe, USA | [25] |
| Ebola | 0.45 (0.19–1.32) | 95% CI | Sierra Leone | [25] |
| Ebola | 0.065 (0.037–0.11) | 95% HPDI | Sierra Leone | [25] |
| Ebola | 0.37 | | Sierra Leone | [25] |
| Ebola | 0.47 | | Sierra Leone | [25] |
| Ebola | 0.27 (0.2–0.33) | 95% CrI | DRC | [25] |
| Ebola | 0.24 | | Africa | [25] |
| Hantavirus (Andes virus)[c] | 0.155 | | | [26] |
| Hepatitis A | 1.71 (1.14–3.4) | 95% CI | China | [29] |
| Hepatitis B[a] | 0.17 | | EU and US | [69] |
| Hepatitis B[a] | 0.128 | | | [70] |
| Hepatitis B[a] | 0.406 | | | [71] |
| Hepatitis E[a] | 0.001 | | | [72] |
| Lassa Fever[d] | 0.14 | | | [73] |
| Marburg [e] | 0.35 | | | – |
| Measles | 0.32 (0.07–1.17) | 95% CI | | [74] |
| Measles | 0.7757 (0.6687–0.899) | 95% CI | | [75] |
| Meningococcal[a] | 0.824 | | US | [76] |
| Meningococcal[a] | 0.311 | | US | [77] |
| MERS-CoV | 0.14 (0.06–0.32) | 95% CI | Global | [43] |
| MERS-CoV | 0.16 (0.11–0.64) | 90% CI | Singapore | [78] |
| MERS-CoV | 0.12 (0.0018–0.1392) | 95% CI | South Korea | [79] |
| MERS-CoV | 0.06 (0.03–0.09) | 95% CI | South Korea and Saudi Arabia | [43] |
| Mpox Clade Ib[f] | 0.257 | | | [80] |
| Mumps[b] | 0.93 | | | – |

*(continued on next page)*

*(continued from previous page)*

| **Pathogen** | **Estimate** | **Uncertainty type** | **Location** | **Reference** |
|---|---|---|---|---|
| Nipah virus[g] | 0.05 | | | [47] |
| Norovirus | 0.42 (0.28–0.62) | 95% CrI | | [81] |
| Paratyphoid Fever[a] | 0.02 | | | [82] |
| Pertussis | 0.93 (0.11–0.53) | 95% CI | | [83] |
| RSV[h] | 0.94 | | | – |
| Rubella[b] | 0.93 | | | – |
| Seasonal Influenza (A/B) | 0.94 (0.59–1.72) | 95% CI | Maryland | [84] |
| Tuberculosis (TB) | 0.16 (0.14–0.17) | 95% CI | Victoria, Australia | [85] |
| Typhoid Fever[a] | 0.003 | | | [82] |
| Yellow Fever[i] | 0.27 | | | – |

*Notes:*

[a] Approximated from outbreak sizes

[b] k taken from pertussis (analogous)

[c] MLE estimate based on transmission chains presented in paper

[d] From version of pareto rule, 1% of cases cause 20% of infections

[e] Using k estimated for Ebola (analogous)

[f] Assuming same k as gonorrhoea (analogous)

[g] Calculated using Pareto rule; 5% of cases cause 86% of transmission

[h] Assuming same k as that of seasonal influenza (analogous)

[i] k calculated using Pareto rule (assuming R0 = 4.18)

Table S5: Estimates of generation time ($g$, in days) obtained for each pathogen following the literature search guidelines from the parameter review protocol.

| Pathogen | Estimate | Uncertainty type | Location | Reference |
|---|---|---|---|---|
| Candida auris | 10 | | UK | [86] |
| Chickenpox[a] | 14 | | | [87] |
| Chickenpox[a] | 14 | | UK | [88] |
| Chickenpox[a] | 14 | | UK | [88] |
| Chickenpox[a] | 15.1 | | UK | [88] |
| Chickenpox[a] | 13.8 | | UK | [88] |
| Cholera | 5 | | | [89] |
| Cholera | 3 | | Bangladesh | [90] |
| COVID-19[a] | 2.99 (2.48–3.49) | 95% CI | | [91] |
| Diphtheria[a] | 7.8 (6.3–9.7) | 95% CrI | | [24] |
| Ebola[a] | 15.4 (13.2–17.5) | 95% CI | | [25] |
| Hantavirus (Andes virus)[a] | 23 (16–30) | SD | Argentina | [26] |
| Hepatitis A | 23.9 (1.6–79.1) | 95% CI | China | [29] |
| Hepatitis A[a] | 27 (20–32) | range | | [92] |
| Hepatitis A | 30.2 (25.2–33) | 95% CI | Italy | [93] |
| Hepatitis B[b] | 90 (60–150) | range | | [94] |
| Hepatitis B | 90 (60–120) | range | | [95] |
| Hepatitis B[b] | 75 (45–180) | range | | [96] |
| Hepatitis E[b] | 29.8 (24.1–36) | 95% CI | UK | [97] |
| Hepatitis E[b] | 36 | | | [98] |
| Hepatitis E[b] | 34 (15–64) | range | | [96] |
| Lassa Fever | 7.5 | | | [99] |
| Marburg | 9 (8.2–10) | 95% CI | Angola | [40] |
| Measles | 12.2 (11.9–12.5) | 95% CI | | [100] |
| Measles | 11.2 (11.1–11.4) | 95% CI | | [100] |
| Measles | 11.9 (11.5–12.1) | 95% CI | | [100] |
| Measles | 11.1 (10.9–11.3) | 95% CI | | [100] |
| Measles | 11.7 (11.3–12) | 95% CI | | [101] |
| Meningococcal | 8 (1–21) | range | Netherlands | [102] |
| MERS-CoV[a] | 14.6 (12.9–16.5) | 95% CI | South Korea | [103] |
| MERS-CoV[a] | 14.13 (13.9–14.7) | 95% CI | Saudi Arabia | [104] |
| MERS-CoV | 7.6 (2.5–23.1) | 95% CI | Saudi Arabia | [105] |
| Mpox Clade Ib[c] | 8.59 (7.18–10.05) | 95% CI | Kamituga and Goma, DRC | [106] |
| Mumps[d] | 18.3 (13.8–24.2) | 95% CI | US (Baltimore) | [107] |
| Mumps[d] | 18 (14.2–22.9) | 95% CI | US (Baltimore) | [107] |
| Nipah virus | 13 (12–14) | IQR | | [47] |
| Norovirus | 3.77 (3.36–4.25) | 95% CI | | [93] |
| Paratyphoid Fever[b] | 5.5 (1–10) | range | | [108] |
| Paratyphoid Fever[b] | 5.5 (1–10) | range | | [96] |
| Pertussis[a] | 9.5 (8.9–10) | 95% CI | | [83] |
| Pertussis[a] | 20.9 (17.9–22.7) | 95% CI | Netherlands | [109] |
| Pertussis[a] | 9.7 (8.73–10.66) | 95% CI | China | [110] |
| Pertussis[a] | 9 (5–13) | IQR | China | [111] |
| RSV | 7.5 (7–8.1) | 95% CI | | [87] |

*(continued on next page)*

*(continued from previous page)*

| **Pathogen** | **Estimate** | **Uncertainty type** | **Location** | **Reference** |
|---|---|---|---|---|
| RSV | 10.38 (8.73–12.47) | 95% CI | | [93] |
| Rubella | 18.3 | | Unspecified | [87] |
| Seasonal Influenza (A/B) | 2.8 (2.7–3) | 95% CI | | [112] |
| Seasonal Influenza (A/B) | 2.5 | | | [113] |
| Seasonal Influenza (A/B) | 3.6 (2.9–4.3) | 95% CI | | [114] |
| Seasonal Influenza (A/B) | 2.5 (1.8–3.3) | 95% CI | | [115] |
| Seasonal Influenza (A/B) | 4.9 (3.3–6.3) | 95% CI | | [115] |
| Seasonal Influenza (A/B) | 3.4 (2.7–4.1) | 95% CI | | [116] |
| Seasonal Influenza (A/B) | 3.4 (2.7–4.1) | 95% CI | | [116] |
| Tuberculosis (TB)[c] | 525.6 (470.85–594.95) | 95% CI | Netherlands | [117] |
| Tuberculosis (TB)[c] | 602.25 | | Lima, Peru | [118] |
| Tuberculosis (TB)[c] | 1277.5 | | Lima, Peru | [118] |
| Tuberculosis (TB)[c] | 206.5 (159.6–267.4) | 95% CI | Netherlands | [119] |
| Typhoid Fever[b] | 18 (6–30) | range | | [108] |
| Typhoid Fever[b] | 11 (3–60) | range | | [96] |
| Yellow Fever | 18 | | | [120] |
| Yellow Fever | 27.5 | | Angola | [121] |

*Notes:*

[a] Article provides estimate of serial interval

[b] Incubation period used as a proxy for generation time

[c] Article provides estimate of serial interval

[d] Article provides estimate of serial interval

Table S6: Central estimates of reproduction number ($R_0$), overdispersion ($k$), and generation time ($g$) obtained using one of three methods per the parameter review protocol.

| Pathogen | Parameter | Estimate | Method |
|---|---|---|---|
| Candida auris | $R_0$ | 0.94 | Single estimate |
| Chickenpox | $R_0$ | 6.24 | Method 2A |
| Cholera | $R_0$ | 1.65 | Method 2A |
| COVID-19 | $R_0$ | 5.94 | Single estimate |
| Diphtheria | $R_0$ | 2.60 | Single estimate |
| Ebola | $R_0$ | 1.81 | Method 2A |
| Hantavirus (Andes virus) | $R_0$ | 2.12 | Single estimate |
| Hepatitis A | $R_0$ | 2.59 | Method 1 |
| Hepatitis B | $R_0$ | 1.98 | Method 2A |
| Hepatitis E | $R_0$ | 3.40 | Method 2B |
| Lassa Fever | $R_0$ | 1.58 | Method 2A |

| Pathogen | Parameter | Estimate | Method |
|---|---|---|---|
| Marburg | $R_0$ | 1.59 | Single estimate |
| Measles | $R_0$ | 14.77 | Method 2A |
| MERS-CoV | $R_0$ | 2.30 | Method 2A |
| Mpox Clade Ib | $R_0$ | 0.90 | Single estimate |
| Mumps | $R_0$ | 5.38 | Method 2B |
| Nipah virus | $R_0$ | 0.33 | Single estimate |
| Norovirus | $R_0$ | 2.45 | Method 2A |
| Paratyphoid Fever | $R_0$ | 1.00 | Single estimate |
| Pertussis | $R_0$ | 5.45 | Method 2A |
| RSV | $R_0$ | 3.94 | Method 2B |
| Rubella | $R_0$ | 5.20 | Method 2A |
| Seasonal Influenza (A/B) | $R_0$ | 1.28 | Single estimate |
| Tuberculosis (TB) | $R_0$ | 1.52 | Method 1 |
| Typhoid Fever | $R_0$ | 1.00 | Single estimate |
| Yellow Fever | $R_0$ | 4.81 | Single estimate |
| Candida auris | k | 0.02 | Single estimate |
| Chickenpox | k | 0.93 | Single estimate |
| Cholera | k | 4.50 | Single estimate |
| COVID-19 | k | 0.41 | Single estimate |
| Diphtheria | k | 0.93 | Single estimate |
| Ebola | k | 0.35 | Method 2A |
| Hantavirus (Andes virus) | k | 0.16 | Single estimate |
| Hepatitis A | k | 1.71 | Single estimate |
| Hepatitis B | k | 0.23 | Method 2B |
| Hepatitis E | k | 0.00 | Single estimate |
| Lassa Fever | k | 0.14 | Single estimate |
| Marburg | k | 0.35 | Single estimate |
| Measles | k | 0.63 | Method 1 |
| MERS-CoV | k | 0.09 | Method 1 |
| Mpox Clade Ib | k | 0.26 | Single estimate |
| Mumps | k | 0.93 | Single estimate |
| Nipah virus | k | 0.05 | Single estimate |
| Norovirus | k | 0.42 | Single estimate |
| Paratyphoid Fever | k | 0.02 | Single estimate |
| Pertussis | k | 0.93 | Single estimate |
| RSV | k | 0.94 | Single estimate |
| Rubella | k | 0.93 | Single estimate |
| Seasonal Influenza (A/B) | k | 0.94 | Single estimate |
| Tuberculosis (TB) | k | 0.16 | Single estimate |
| Typhoid Fever | k | 0.00 | Single estimate |
| Yellow Fever | k | 0.27 | Single estimate |
| Candida auris | g | 10.00 | Single estimate |
| Chickenpox | g | 14.00 | Method 2A |
| Cholera | g | 4.00 | Method 2B |
| COVID-19 | g | 2.99 | Single estimate |
| Diphtheria | g | 7.80 | Single estimate |
| Ebola | g | 15.40 | Single estimate |
| Hantavirus (Andes virus) | g | 23.00 | Single estimate |
| Hepatitis A | g | 27.03 | Method 2B |
| Hepatitis B | g | 85.00 | Method 2B |
| Hepatitis E | g | 33.27 | Method 2B |
| Lassa Fever | g | 7.50 | Single estimate |

| Pathogen | Parameter | Estimate | Method |
|---|---|---|---|
| Marburg | g | 9.00 | Single estimate |
| Measles | g | 11.61 | Method 1 |
| MERS-CoV | g | 14.14 | Method 1 |
| Mpox Clade Ib | g | 8.59 | Single estimate |
| Mumps | g | 18.12 | Method 1 |
| Nipah virus | g | 13.00 | Single estimate |
| Norovirus | g | 3.77 | Single estimate |
| Paratyphoid Fever | g | 5.50 | Method 2B |
| Pertussis | g | 12.27 | Method 2B |
| RSV | g | 8.80 | Method 1 |
| Rubella | g | 18.30 | Single estimate |
| Seasonal Influenza (A/B) | g | 3.08 | Method 2A |
| Tuberculosis (TB) | g | 652.96 | Method 2B |
| Typhoid Fever | g | 14.50 | Method 2B |
| Yellow Fever | g | 22.75 | Method 2B |

##### (E) Impact assessment

We can quantify the impact of an infectious disease risk related to the World Cup in two cost terms:

1. The per case cost (assuming symptomatic infection equates to a case)
2. The cost of the pathogen establishing endemicity

The focus of the impact calculations is to capture an approximate cost, therefore rough estimations and assumptions are satisfactory in almost all cases for the purposes of our assessment.

The impact per case can be further broken down to:

1. Direct costs (i.e. medical costs, public health response costs)
2. Indirect costs (i.e. productivity losses)

For pathogens that have an available empirical impact cost per case that includes both direct and indirect costs, no further calculation of the individual case impact is needed. In all other instances, we will use the following calculation to estimate the impact per case.

Let $I_p$ be the per case impact of pathogen $p$. We can express $I_p$ as:

$$I_p = \textit{Direct costs per case of p in USD} + \textit{Indirect costs per case of p in USD}$$

Direct costs are composed of the following:

- Medical costs - These are the costs incurred during hospitalization or outpatient treatment. If multiple costs are presented for different levels of disease severity, these will be included in our estimate at the same proportion that they are present in the population. E.g. If 70% of norovirus infections receive outpatient treatment costing $100, and the remaining 30% receive hospitalization costing $10,000, our overall estimate will be: (100 * 0.7) + (10,000 * 0.3) = $3,070.
- Public health response - These are costs incurred by public health departments when responding to an outbreak of a pathogen, incorporating costs such as outbreak investigation, quarantine, and contact tracing. It is important to note that we did not include a fixed response cost in our calculations and instead distributed this over the cost per case. In cases where we could not find a public health cost for the pathogen or an analogous pathogen, we assumed that the cost of the response was negligible.

Indirect costs are composed of the following: - Productivity losses - These are costs incurred as a result of morbidity impacting a patient's ability to work or premature death. In cases where this information was not readily available, we estimated the loss by multiplying the DALYs per case (calculated from existing literature or the Global Burden of Disease Study online tool) and the 2026 GDP per capita for the U.S.

(extracted from the International Monetary Fund). For pathogens where local spread in the US are not expected (i.e. only imported cases are expected), productivity losses were not included in the total cost per case.

Note that for some pathogens it was relevant to calculate the impact per infection. In these situations, impact per infection, $J_p$ was calculated as follows:

$$J_p = I_p \times (1 - \textit{asymptomatic proportion of } p)$$

For non-endemic pathogens that have the potential to become endemic, we must also include the cost associated with establishing endemicity (e.g. the future productivity losses, medical costs, and cost of control). This cost is calculated at the pathogen level.

Let $M_p$ be the endemicity impact of pathogen $p$. We can express $M_p$ as:

$$M_p = \textit{Risk of long term endemicity of } p \times \textit{Cost of endemicity of } p \textit{ in USD}$$

For the purposes of this assessment, the cost of endemicity is considered to be very large. Therefore, any disease with a potential to establish long term endemicity is assumed to have a considerable impact and is subsequently included on the priority list.

| Pathogen | Total Cost per Case in USD (ref) | Total Cost per Infection in USD (ref) |
|---|---|---|
| Brucellosis | 20,450 [122] | |
| Crimean-Congo Hemorrhagic Fever | 1,830 [123] | |
| Chickenpox | 730 [124] | |
| Cholera | 1,190 [125] [122] | |
| COVID-19 | 488 [126] [127] | 366 [128] |
| Diptheria | 179,000 [127] [129] | |
| Escherichia coli | 7,860 [122] | |
| Ebola | 4,300,000 [130] | |
| Hantavirus | 500,000 [131] | |
| Hepatitis A | 67,700 [122] | |
| Hepatitis B | 25,700 [127] [132] [133] [134] | 10,300 [94] |
| Hepatitis E | 67,700 [122] | |
| Lassa Fever | 4,300,000 [130] | |
| Marburg | 4,300,000 [130] | |
| Measles | 45,600 [135] | |
| Meningococcal Disease | 413,000 [127] [136] [137] | |
| Mpox Clade Ib | 3,840 [138] | |
| Mumps | 3,950 [139] | |
| Norovirus | 636 [122] [140] | 496 [141] |
| Oropouche | 2700 [142] | |
| Paratyphoid Fever | 1,490 [122] | |
| Pertussis | 9,320 [127] [143] [144] [145] [146] | |
| Pneumonia (bacterial) | 8,900 [147] | |
| RSV | | 688 [148] [149] [150] [151] |
| Rubella | 9,340 [152] [153] [154] [155] | |
| Salmonella | 18,100 [122] | |
| Seasonal Influenza (A/B) | 1,510 [152] [156] | 1,230 [157] |
| Tuberculosis | 73,720 [158] [159] | 39,800 [160] [161] |
| Typhoid Fever | 1,490 [122] | |
| Yellow Fever | 259,000 [127] [162] | |

Costs were adjusted for inflation using the U.S. Bureau of Labor Statistics' CPI Inflation Calculator [163]. Costs reported in other currencies were converted to U.S. dollars using the IRS' Yearly Average Currency Exchange Rates [164].

### References


1. Global Mpox Trends. Available from: https://worldhealthorg.shinyapps.io/mpx_global/ [Accessed on: 2026 May 28]
2. Organization WH. Global dengue surveillance. Available from: https://worldhealthorg.shinyapps.io/dengue_global/ [Accessed on: 2026 May 28]
3. WHO. WHO Immunization Data portal - Measles. en. Available from: https://immunizationdata.who.int/global/wiise-detail-page [Accessed on: 2026 May 28]
4. Cholera worldwide overview. en. 2026 Apr. Available from: https://www.ecdc.europa.eu/en/all-topics-z/cholera/surveillance-and-disease-data/cholera-monthly [Accessed on: 2026 May 28]
5. WHO. WHO Immunization Data portal - Pertussis. Available from: https://immunizationdata.who.int/global/wiise-detail-page/pertussis-reported-cases-and-incidence?CODE=Global&YEAR= [Accessed on: 2026 May 28]
6. WHO. WHO Immunization Data portal - Diphtheria. en. Available from: https://immunizationdata.who.int/global/wiise-detail-page [Accessed on: 2026 May 28]
7. WHO. WHO Immunization Data portal - Yellow Fever. en. Available from: https://immunizationdata.who.int/global/wiise-detail-page [Accessed on: 2026 May 28]
8. WHO. WHO Immunization Data portal - Typhoid. en. Available from: https://immunizationdata.who.int/global/wiise-detail-page [Accessed on: 2026 May 28]
9. WHO. WHO Immunization Data portal - Rubella. en. Available from: https://immunizationdata.who.int/global/wiise-detail-page [Accessed on: 2026 May 28]
10. WHO. WHO Immunization Data portal - Mumps. en. Available from: https://immunizationdata.who.int/global/wiise-detail-page [Accessed on: 2026 May 28]
11. Disease Control and Prevention EC for. Chikungunya virus disease worldwide overview. en. 2026 Mar. Available from: https://www.ecdc.europa.eu/en/chikungunya-monthly [Accessed on: 2026 May 28]
12. Balcan D, Gonçalves B, Hu H, Ramasco JJ, Colizza V, and Vespignani A. Modeling the spatial spread of infectious diseases: the GLobal Epidemic and Mobility computational model. Journal of computational science 2010 Aug; 1:132–45. DOI: 10.1016/j.jocs.2010.07.002. Available from: https://pmc.ncbi.nlm.nih.gov/articles/PMC3056392/ [Accessed on: 2026 May 28]
13. Davis JT, Chinazzi M, Perra N, Mu K, Piontti APY, Ajelli M, Dean NE, Gioannini C, Litvinova M, Merler S, Rossi L, Sun K, Xiong X, Halloran ME, Longini IM, Viboud C, and Vespignani A. Cryptic transmission of SARS-CoV-2 and the first COVID-19 wave in Europe and the United States. eng. 2021 Mar. DOI: 10.1101/2021.03.24.21254199
14. Paul P, Forsberg K, Vallabhaneni S, Lockhart SR, Litvintseva AP, Kerins JL, Tang AS, Jegede O, Barrett PM, Ross K, Slayton R, and Jernigan JA. 1268. Transmissibility of Candida auris by Type of Inpatient Healthcare Facility. Open Forum Infectious Diseases 2018 Nov; 5:S386–S387. DOI: 10.1093/ofid/ofy210.1101. Available from: https://pmc.ncbi.nlm.nih.gov/articles/PMC6252786/ [Accessed on: 2026 May 20]
15. Orenstein WA, Offit PA, Edwards KM, and Plotkin SA, eds. Plotkin's vaccines. eng. 8th edition. Philadelphia, PA: Elsevier, 2024
16. A. Nardone, F. de Ory, M. Carton, D. Cohen, P. van Damme, I. Davidkin, M.C. Rota, H. de Melker, J. Mossong, M. Slacikova, A. Tischer, N. Andrews, G. Berbers, G. Gabutti, N. Gay, L. Jones, S. Jokinen, G. Kafatos, M.V. Martinez de Aragon, F. Schneider, Z. Smetana, B. Vargova, R. Vranckx, and E. Miller. The comparative sero-epidemiology of varicella zoster virus in 11 countries in the European region. VAccine 2007; 25:7866–72. DOI: 10.1016/j.vaccine.2007.07.036
17. Anderson RM and May RM. Directly transmitted infections diseases: control by vaccination. eng. Science 1982 Feb; 215:1053–60. DOI: 10.1126/science.7063839
18. Mukandavire Z, Smith DL, and Morris Jr JG. Cholera in Haiti: Reproductive numbers and vaccination coverage estimates. en. Scientific Reports 2013 Jan; 3:997. DOI: 10.1038/srep00997. Available from: https://www.nature.com/articles/srep00997 [Accessed on: 2026 May 20]

19. Smirnova A, Sterrett N, Mujica OJ, Munayco C, Suárez L, Viboud C, and Chowell G. Spatial dynamics and the basic reproduction number of the 1991–1997 Cholera epidemic in Peru. en. PLOS Neglected Tropical Diseases 2020 Jul; 14:e0008045. DOI: 10.1371/journal.pntd.0008045. Available from: https://journals.plos.org/plosntds/article?id=10.1371/journal.pntd.0008045 [Accessed on: 2026 May 20]
20. Mukandavire Z, Liao S, Wang J, Gaff H, Smith DL, and Morris JG. Estimating the reproductive numbers for the 2008–2009 cholera outbreaks in Zimbabwe. Proceedings of the National Academy of Sciences 2011 May; 108:8767–72. DOI: 10.1073/pnas.1019712108. Available from: https://www.pnas.org/doi/full/10.1073/pnas.1019712108 [Accessed on: 2026 May 20]
21. Adeniyi EO. Transmission Dynamics And Control Of Cholera In Africa: A Mathematical Modelling Approach. en. 2024 Nov. Available from: https://hdl.handle.net/10315/42813 [Accessed on: 2026 May 20]
22. Moore SM, Shannon KL, Zelaya CE, Azman AS, and Lessler J. Epidemic Risk from Cholera Introductions into Mexico. PLoS Currents 2014 Feb; 6:ecurrents.outbreaks.c04478c7fbd9854ef6ba923cc81eb799. DOI: 10.1371/currents.outbreaks.c04478c7fbd9854ef6ba923cc81eb799. Available from: https://pmc.ncbi.nlm.nih.gov/articles/PMC3933092/ [Accessed on: 2026 May 20]
23. Du Z, Liu C, Wang C, Xu L, Xu M, Wang L, Bai Y, Xu X, Lau EHY, Wu P, and Cowling BJ. Reproduction Numbers of Severe Acute Respiratory Syndrome Coronavirus 2 (SARS-CoV-2) Variants: A Systematic Review and Meta-analysis. Clinical Infectious Diseases 2022 Jul; 75:e293–e295. DOI: 10.1093/cid/ciac137. Available from: https://doi.org/10.1093/cid/ciac137 [Accessed on: 2026 May 18]
24. Truelove SA, Keegan LT, Moss WJ, Chaisson LH, Macher E, Azman AS, and Lessler J. Clinical and Epidemiological Aspects of Diphtheria: A Systematic Review and Pooled Analysis. en. Clinical Infectious Diseases 2020 Jun; 71:89–97. DOI: 10.1093/cid/ciz808. Available from: https://academic.oup.com/cid/article/71/1/89/5551532 [Accessed on: 2026 Apr 27]
25. Nash RK, Bhatia S, Morgenstern C, Doohan P, Jorgensen D, McCain K, McCabe R, Nikitin D, Forna A, Cuomo-Dannenburg G, Hicks JT, Sheppard RJ, Naidoo T, Van Elsland S, Geismar C, Rawson T, Leuba SI, Wardle J, Routledge I, Fraser K, Imai-Eaton N, Cori A, and Unwin HJT. Ebola virus disease mathematical models and epidemiological parameters: a systematic review. en. The Lancet Infectious Diseases 2024 Dec; 24:e762–e773. DOI: 10.1016/S1473-3099(24)00374-8. Available from: https://linkinghub.elsevier.com/retrieve/pii/S1473309924003748 [Accessed on: 2026 Apr 27]
26. Martínez VP, Di Paola N, Alonso DO, Pérez-Sautu U, Bellomo CM, Iglesias AA, Coelho RM, López B, Periolo N, Larson PA, Nagle ER, Chitty JA, Pratt CB, Díaz J, Cisterna D, Campos J, Sharma H, Dighero-Kemp B, Biondo E, Lewis L, Anselmo C, Olivera CP, Pontoriero F, Lavarra E, Kuhn JH, Strella T, Edelstein A, Burgos MI, Kaler M, Rubinstein A, Kugelman JR, Sanchez-Lockhart M, Perandones C, and Palacios G. "Super-Spreaders" and Person-to-Person Transmission of Andes Virus in Argentina. en. New England Journal of Medicine 2020 Dec; 383:2230–41. DOI: 10.1056/NEJMoa2009040. Available from: http://www.nejm.org/doi/10.1056/NEJMoa2009040 [Accessed on: 2026 Apr 28]
27. Regan DG, Wood JG, Benevent C, Ali H, Smith LW, Robertson PW, Ferson MJ, Fairley CK, Donovan B, and Law MG. Estimating the critical immunity threshold for preventing hepatitis A outbreaks in men who have sex with men. en. Epidemiology and Infection 2016 May; 144:1528–37. DOI: 10.1017/S0950268815002605. Available from: https://www.cambridge.org/core/product/identifier/S0950268815002605/type/journal_article [Accessed on: 2026 Apr 27]
28. Dankwa EA, Donnelly CA, Brouwer AF, Zhao R, Montgomery MP, Weng MK, and Martin NK. Estimating vaccination threshold and impact in the 2017–2019 hepatitis A virus outbreak among persons experiencing homelessness or who use drugs in Louisville, Kentucky, United States. en. Vaccine 2021 Dec; 39:7182–90. DOI: 10.1016/j.vaccine.2021.10.001. Available from: https://linkinghub.elsevier.com/retrieve/pii/S0264410X21013086 [Accessed on: 2026 Apr 27]
29. Zhang XS and Iacono GL. Estimating human-to-human transmissibility of hepatitis A virus in an outbreak at an elementary school in China, 2011. en. PLOS ONE 2018 Sep; 13. Ed. by Lau EH:e0204201. DOI: 10.1371/journal.pone.0204201. Available from: https://dx.plos.org/10.1371/journal.pone.0204201 [Accessed on: 2026 Apr 27]

30. Mann J and Roberts M. Modelling the epidemiology of hepatitis B in New Zealand. en. Journal of Theoretical Biology 2011 Jan; 269:266–72. DOI: 10.1016/j.jtbi.2010.10.028. Available from: https://linkinghub.elsevier.com/retrieve/pii/S002251931000562X [Accessed on: 2026 Apr 27]
31. Williams JR, Nokes DJ, Medley GF, and Anderson RM. The transmission dynamics of hepatitis B in the UK: a mathematical model for evaluating costs and effectiveness of immunization programmes. eng. Epidemiology and Infection 1996 Feb; 116:71–89. DOI: 10.1017/s0950268800058970
32. Kretzschmar M, Wit GA de, Smits LJM, and Laar MJW van de. Vaccination against hepatitis B in low endemic countries. eng. Epidemiology and Infection 2002 Apr; 128:229–44. DOI: 10.1017/s0950268801006562
33. Zou L, Zhang W, and Ruan S. Modeling the transmission dynamics and control of hepatitis B virus in China. Journal of Theoretical Biology 2010 Jan; 262:330–8. DOI: 10.1016/j.jtbi.2009.09.035. Available from: https://www.sciencedirect.com/science/article/pii/S0022519309004731 [Accessed on: 2026 May 20]
34. Pang J, Cui Ja, and Zhou X. Dynamical behavior of a hepatitis B virus transmission model with vaccination. Journal of Theoretical Biology 2010 Aug; 265:572–8. DOI: 10.1016/j.jtbi.2010.05.038. Available from: https://www.sciencedirect.com/science/article/pii/S002251931000281X [Accessed on: 2026 May 20]
35. Thornley S, Bullen C, and Roberts M. Hepatitis B in a high prevalence New Zealand population: A mathematical model applied to infection control policy. Journal of Theoretical Biology 2008 Oct; 254:599–603. DOI: 10.1016/j.jtbi.2008.06.022. Available from: https://www.sciencedirect.com/science/article/pii/S0022519308003342 [Accessed on: 2026 May 20]
36. Nannyonga B, Sumpter DJT, Mugisha JYT, and Luboobi LS. The Dynamics, Causes and Possible Prevention of Hepatitis E Outbreaks. en. PLOS ONE 2012 Jul; 7:e41135. DOI: 10.1371/journal.pone.0041135. Available from: https://journals.plos.org/plosone/article?id=10.1371/journal.pone.0041135 [Accessed on: 2026 May 20]
37. Morley M, Majumder MS, Gallanis T, and Wilson J. Using Non-traditional Data Sources for Near Real-Time Estimation of Transmission Dynamics in the Hepatitis-E Outbreak in Namibia, 2017–2018. en. *Leveraging Data Science for Global Health*. Ed. by Celi LA, Majumder MS, Ordóñez P, Osorio JS, Paik KE, and Somai M. Cham: Springer International Publishing, 2020 :443–52. DOI: 10.1007/978-3-030-47994-7_28. Available from: https://doi.org/10.1007/978-3-030-47994-7_28 [Accessed on: 2026 May 20]
38. Cooper BS, White LJ, and Siddiqui R. Reactive and pre-emptive vaccination strategies to control hepatitis E infection in emergency and refugee settings: A modelling study. en. PLOS Neglected Tropical Diseases 2018 Sep; 12:e0006807. DOI: 10.1371/journal.pntd.0006807. Available from: https://journals.plos.org/plosntds/article?id=10.1371/journal.pntd.0006807 [Accessed on: 2026 May 20]
39. Wang J, Zhao S, Chen X, Huang Z, Chong MKC, Guo Z, Javanbakht M, and Ran J. The reproductive number of Lassa fever: a systematic review. en. 2021. Available from: https://dx.doi.org/10.1093/jtm/taab029 [Accessed on: 2026 May 21]
40. Ajelli M and Merler S. Transmission Potential and Design of Adequate Control Measures for Marburg Hemorrhagic Fever. en. PLoS ONE 2012 Dec; 7. Ed. by Bausch DG:e50948. DOI: 10.1371/journal.pone.0050948. Available from: https://dx.plos.org/10.1371/journal.pone.0050948 [Accessed on: 2026 Apr 27]
41. Guerra FM, Bolotin S, Lim G, Heffernan J, Deeks SL, Li Y, and Crowcroft NS. The basic reproduction number (R0) of measles: a systematic review. en. The Lancet Infectious Diseases 2017 Dec; 17:e420–e428. DOI: 10.1016/S1473-3099(17)30307-9. Available from: https://linkinghub.elsevier.com/retrieve/pii/S1473309917303079 [Accessed on: 2026 Apr 27]
42. Lo Presti A, Vacca P, Neri A, Fazio C, Ambrosio L, Rezza G, and Stefanelli P. Estimates of the reproductive numbers and demographic reconstruction of outbreak associated with C:P1.5–1,10–8:F3–6:ST–11(cc11) Neisseria meningitidis strains. en. Infection, Genetics and Evolution 2020 Oct; 84:104360. DOI: 10.1016/j.meegid.2020.104360. Available from: https://linkinghub.elsevier.com/retrieve/pii/S156713482030191X [Accessed on: 2026 Apr 27]
43. Chowell G, Abdirizak F, Lee S, Lee J, Jung E, Nishiura H, and Viboud C. Transmission characteristics of MERS and SARS in the healthcare setting: a comparative study. BMC Medicine 2015 Sep; 13:210.

DOI: 10.1186/s12916-015-0450-0. Available from: https://pmc.ncbi.nlm.nih.gov/articles/PMC4558759/ [Accessed on: 2026 May 11]
44. Majumder MS, Rivers C, Lofgren E, and Fisman D. Estimation of MERS-Coronavirus Reproductive Number and Case Fatality Rate for the Spring 2014 Saudi Arabia Outbreak: Insights from Publicly Available Data. PLoS Currents 2014 Dec; 6:ecurrents.outbreaks.98d2f8f3382d84f390736cd5f5fe133c. DOI: 10.1371/currents.outbreaks.98d2f8f3382d84f390736cd5f5fe133c. Available from: https://pmc.ncbi.nlm.nih.gov/articles/PMC4322060/ [Accessed on: 2026 Apr 27]
45. Kamadjeu R, Nsengimana F, Nimpagaritse M, Moturi E, Kezakarayagwa E, Nkiko R, Ngendakumana J, Nimubona R, Nkengurutse L, Ba H, Nizigiyimana D, Ngwakum P, Diouf MS, Bégin F, Nyandwi J, and Noble DJ. Characterising household transmission dynamics of clade Ib mpox in Burundi: a prospective cohort study. The Lancet Infectious Diseases 2026 Feb; 26:182–9. DOI: 10.1016/S1473-3099(25)00483-9. Available from: https://www.sciencedirect.com/science/article/pii/S1473309925004839 [Accessed on: 2026 May 20]
46. Li Y, Liu X, and Wang L. Modelling the Transmission Dynamics and Control of Mumps in Mainland China. en. International Journal of Environmental Research and Public Health 2017 Dec; 15:33. DOI: 10.3390/ijerph15010033. Available from: https://www.mdpi.com/1660-4601/15/1/33 [Accessed on: 2026 Apr 27]
47. Nikolay B, Salje H, Hossain MJ, Khan AD, Sazzad HM, Rahman M, Daszak P, Ströher U, Pulliam JR, Kilpatrick AM, Nichol ST, Klena JD, Sultana S, Afroj S, Luby SP, Cauchemez S, and Gurley ES. Transmission of Nipah Virus — 14 Years of Investigations in Bangladesh. The New England journal of medicine 2019 May; 380:1804–14. DOI: 10.1056/NEJMoa1805376. Available from: https://pmc.ncbi.nlm.nih.gov/articles/PMC6547369/ [Accessed on: 2026 Apr 28]
48. Wang L, Ji L, Li H, Xu D, Chen L, Zhang P, and Wang W. Early evolution and transmission of GII.P16-GII.2 norovirus in China. en. G3 Genes|Genomes|Genetics 2022 Nov; 12. Ed. by Kulathinal R:jkac250. DOI: 10.1093/g3journal/jkac250. Available from: https://academic.oup.com/g3journal/article/doi/10.1093/g3journal/jkac250/6705237 [Accessed on: 2026 May 19]
49. Steele MK, Wikswo ME, Hall AJ, Koelle K, Handel A, Levy K, Waller LA, and Lopman BA. Characterizing Norovirus Transmission from Outbreak Data, United States - Volume 26, Number 8—August 2020 - Emerging Infectious Diseases journal - CDC. en-us. 2020. DOI: 10.3201/eid2608.191537. Available from: https://wwwnc.cdc.gov/eid/article/26/8/19-1537_article [Accessed on: 2026 May 1]
50. Schulz C, Wyatt A, Walker J, Smoll N, Field E, and Khandaker G. Outbreak investigation of norovirus gastroenteritis in a childcare facility in Central Queensland, Australia: a household level case series analysis. en. Communicable Diseases Intelligence 2024 Aug; 48. DOI: 10.33321/cdi.2024.48.46. Available from: https://ojs.cdi.cdc.gov.au/index.php/cdi/article/view/2705 [Accessed on: 2026 May 19]
51. Solano R, Alseda M, Godoy P, Sanz M, Bartolomé R, Manzanares-Laya S, Domínguez À, Caylà JA, and Catalonia atWGftSoAGi. Person-to-person transmission of norovirus resulting in an outbreak of acute gastroenteritis at a summer camp. en-US. European Journal of Gastroenterology & Hepatology 2014 Oct; 26:1160. DOI: 10.1097/MEG.0000000000000179. Available from: https://journals.lww.com/eurojgh/abstract/2014/10000/person_to_person_transmission_of_norovirus.14.aspx [Accessed on: 2026 May 19]
52. Vesga JF, Douglas A, Celma C, Knock ES, Baguelin M, and Edmunds WJ. The transmission dynamics of Norovirus in England: A genotype-specific modelling study. Epidemics 2025 Dec; 53:100875. DOI: 10.1016/j.epidem.2025.100875. Available from: https://www.sciencedirect.com/science/article/pii/S1755436525000635 [Accessed on: 2026 May 19]
53. Pitzer VE, Bowles CC, Baker S, Kang G, Balaji V, Farrar JJ, and Grenfell BT. Predicting the Impact of Vaccination on the Transmission Dynamics of Typhoid in South Asia: A Mathematical Modeling Study. PLoS Neglected Tropical Diseases 2014 Jan; 8:e2642. DOI: 10.1371/journal.pntd.0002642. Available from: https://pmc.ncbi.nlm.nih.gov/articles/PMC3886927/ [Accessed on: 2026 May 20]
54. Kretzschmar M, Teunis PFM, and Pebody RG. Incidence and Reproduction Numbers of Pertussis: Estimates from Serological and Social Contact Data in Five European Countries. en. PLoS Medicine 2010 Jun; 7. Ed. by Murray M:e1000291. DOI: 10.1371/journal.pmed.1000291. Available from: https://dx.plos.org/10.1371/journal.pmed.1000291 [Accessed on: 2026 Apr 27]

55. Poletti P, Merler S, Ajelli M, Manfredi P, Munywoki PK, Nokes DJ, and Melegaro A. Evaluating vaccination strategies for reducing infant respiratory syncytial virus infection in low-income settings. en. BMC Medicine 2015 Mar; 13:49. DOI: 10.1186/s12916-015-0283-x. Available from: https://doi.org/10.1186/s12916-015-0283-x [Accessed on: 2026 May 11]
56. Reis J and Shaman J. Retrospective Parameter Estimation and Forecast of Respiratory Syncytial Virus in the United States. en. PLOS Computational Biology 2016 Oct; 12:e1005133. DOI: 10.1371/journal.pcbi.1005133. Available from: https://journals.plos.org/ploscompbiol/article?id=10.1371/journal.pcbi.1005133 [Accessed on: 2026 May 11]
57. Pitzer VE, Viboud C, Alonso WJ, Wilcox T, Metcalf CJ, Steiner CA, Haynes AK, and Grenfell BT. Environmental Drivers of the Spatiotemporal Dynamics of Respiratory Syncytial Virus in the United States. en. PLOS Pathogens 2015 Jan; 11:e1004591. DOI: 10.1371/journal.ppat.1004591. Available from: https://journals.plos.org/plospathogens/article?id=10.1371/journal.ppat.1004591 [Accessed on: 2026 May 11]
58. Weber A, Weber M, and Milligan P. Modeling epidemics caused by respiratory syncytial virus (RSV). Mathematical Biosciences 2001 Aug; 172:95–113. DOI: 10.1016/S0025-5564(01)00066-9. Available from: https://www.sciencedirect.com/science/article/pii/S0025556401000669 [Accessed on: 2026 May 21]
59. Papadopoulos T and Vynnycky E. Estimates of the basic reproduction number for rubella using seroprevalence data and indicator-based approaches. en. PLOS Computational Biology 2022 Mar; 18. Ed. by Althouse B:e1008858. DOI: 10.1371/journal.pcbi.1008858. Available from: https://dx.plos.org/10.1371/journal.pcbi.1008858 [Accessed on: 2026 Apr 27]
60. Edmunds WJ, Heijden OGVD, Eerola M, and Gay NJ. Modelling rubella in Europe. en. Epidemiology & Infection 2000 Dec; 125:617–34. DOI: 10.1017/S0950268800004660. Available from: https://www.cambridge.org/core/journals/epidemiology-and-infection/article/modelling-rubella-in-europe/ED097142632D66A21F8876A009746F4A [Accessed on: 2026 May 18]
61. Lessler J and Metcalf CJE. Balancing Evidence and Uncertainty when Considering Rubella Vaccine Introduction. en. PLOS ONE 2013 Jul; 8:e67639. DOI: 10.1371/journal.pone.0067639. Available from: https://journals.plos.org/plosone/article?id=10.1371/journal.pone.0067639 [Accessed on: 2026 May 18]
62. Biggerstaff M, Cauchemez S, Reed C, Gambhir M, and Finelli L. Estimates of the reproduction number for seasonal, pandemic, and zoonotic influenza: a systematic review of the literature. en. BMC Infectious Diseases 2014 Sep; 14:480. DOI: 10.1186/1471-2334-14-480. Available from: https://doi.org/10.1186/1471-2334-14-480 [Accessed on: 2026 Apr 27]
63. Stadler T. Inferring Epidemiological Parameters on the Basis of Allele Frequencies. Genetics 2011 Jul; 188:663–72. DOI: 10.1534/genetics.111.126466. Available from: https://pmc.ncbi.nlm.nih.gov/articles/PMC3176535/ [Accessed on: 2026 May 20]
64. Aandahl RZ, Stadler T, Sisson SA, and Tanaka MM. Exact vs. approximate computation: reconciling different estimates of Mycobacterium tuberculosis epidemiological parameters. eng. Genetics 2014 Apr; 196:1227–30. DOI: 10.1534/genetics.113.158808
65. Liu Y and Rocklöv J. What is the reproductive number of yellow fever? Journal of Travel Medicine 2020 Nov; 27:taaa156. DOI: 10.1093/jtm/taaa156. Available from: https://doi.org/10.1093/jtm/taaa156 [Accessed on: 2026 May 1]
66. Lyman M, Forsberg K, Reuben J, Dang T, Free R, Seagle EE, Sexton DJ, Soda E, Jones H, Hawkins D, Anderson A, Bassett J, Lockhart SR, Merengwa E, Iyengar P, Jackson BR, and Chiller T. Notes from the Field: Transmission of Pan-Resistant and Echinocandin-Resistant Candida auris in Health Care Facilities Texas and the District of Columbia, January–April 2021. Morbidity and Mortality Weekly Report 2021 Jul; 70:1022–3. DOI: 10.15585/mmwr.mm7029a2. Available from: https://pmc.ncbi.nlm.nih.gov/articles/PMC8297693/ [Accessed on: 2026 May 20]
67. Prestel C, Anderson E, Forsberg K, Lyman M, Perio MA de, Kuhar D, Edwards K, Rivera M, Shugart A, Walters M, and Dotson NQ. Candida auris Outbreak in a COVID-19 Specialty Care Unit — Florida, July–August 2020. Morbidity and Mortality Weekly Report 2021 Jan; 70:56–7. DOI: 10.15585/mmwr.mm7002e3. Available from: https://pmc.ncbi.nlm.nih.gov/articles/PMC7808709/ [Accessed on: 2026 May 20]

68. Wegehaupt O, Endo A, and Vassall A. Superspreading, overdispersion and their implications in the SARS-CoV-2 (COVID-19) pandemic: a systematic review and meta-analysis of the literature. en. BMC Public Health 2023 May; 23:1003. DOI: 10.1186/s12889-023-15915-1. Available from: https://doi.org/10.1186/s12889-023-15915-1 [Accessed on: 2026 Apr 27]
69. Lanini S, Puro V, Lauria FN, Fusco FM, Nisii C, and Ippolito G. Patient to patient transmission of hepatitis B virus: a systematic review of reports on outbreaks between 1992 and 2007. en. BMC Medicine 2009 Apr; 7:15. DOI: 10.1186/1741-7015-7-15. Available from: https://doi.org/10.1186/1741-7015-7-15 [Accessed on: 2026 May 18]
70. Sacuk AV, , Solopova GG, , Ploskireva AA, and . A systematic review of outbreaks of bloodborne infections (hepatitis B and C, HIV) transmitted from patient to patient in healthcare settings. ru; en. Journal of microbiology, epidemiology and immunobiology 2021 Jul; 98:319–30. DOI: 10.36233/0372-9311-112. Available from: https://doi.org/10.36233/0372-9311-112 [Accessed on: 2026 May 18]
71. Fabrizi F, Dixit V, Messa P, and Martin P. Transmission of Hepatitis B Virus in Dialysis Units: A Systematic Review of Reports on Outbreaks. EN. The International Journal of Artificial Organs 2015 Jan; 38:1–7. DOI: 10.5301/ijao.5000376. Available from: https://doi.org/10.5301/ijao.5000376 [Accessed on: 2026 May 18]
72. Hakim MS, Wang W, Bramer WM, Geng J, Huang F, Man RA de, Peppelenbosch MP, and Pan Q. The global burden of hepatitis E outbreaks: a systematic review. eng. Liver International: Official Journal of the International Association for the Study of the Liver 2017 Jan; 37:19–31. DOI: 10.1111/liv.13237
73. Lo Iacono G, Cunningham AA, Fichet-Calvet E, Garry RF, Grant DS, Khan SH, Leach M, Moses LM, Schieffelin JS, Shaffer JG, Webb CT, and Wood JLN. Using Modelling to Disentangle the Relative Contributions of Zoonotic and Anthroponotic Transmission: The Case of Lassa Fever. en. PLoS Neglected Tropical Diseases 2015 Jan; 9. Ed. by McElroy AK:e3398. DOI: 10.1371/journal.pntd.0003398. Available from: https://dx.plos.org/10.1371/journal.pntd.0003398 [Accessed on: 2026 Apr 29]
74. Hiroshi Nishiura, Kenji Mizumoto, and Yusuke Asai. Assessing the transmission dynamics of measles in Japan, 2016. Epidemics 2017; 20:67–72. DOI: 10.1016/j.epidem.2017.03.005
75. Adebanji AO, Aschl F, Chumo EC, Owiredu EO, Müller J, and Mbegalo T. Social Response and Measles Dynamics. en. Stats 2023 Nov; 6:1280–97. DOI: 10.3390/stats6040079. Available from: https://www.mdpi.com/2571-905X/6/4/79 [Accessed on: 2026 Apr 27]
76. Mbaeyi SA, Blain A, Whaley MJ, Wang X, Cohn AC, and MacNeil JR. Epidemiology of Meningococcal Disease Outbreaks in the United States, 2009–2013. Clinical Infectious Diseases 2019 Feb; 68:580–5. DOI: 10.1093/cid/ciy548. Available from: https://doi.org/10.1093/cid/ciy548 [Accessed on: 2026 May 18]
77. Atkinson B, Gandhi A, and Balmer P. History of Meningococcal Outbreaks in the United States: Implications for Vaccination and Disease Prevention. en. Pharmacotherapy: The Journal of Human Pharmacology and Drug Therapy 2016; 36. _eprint: https://accpjournals.onlinelibrary.wiley.com/doi/pdf/10.1002/phar.1790:8 92. DOI: 10.1002/phar.1790. Available from: https://onlinelibrary.wiley.com/doi/abs/10.1002/phar.1790 [Accessed on: 2026 May 18]
78. Kucharski AJ and Althaus CL. The role of superspreading in Middle East respiratory syndrome coronavirus (MERS-CoV) transmission. en. Eurosurveillance 2015 Jun; 20:21167. DOI: 10.2807/1560-7917.ES2015.20.25.21167. Available from: https://www.eurosurveillance.org/content/10.2807/1560-7917.ES2015.20.25.21167 [Accessed on: 2026 May 19]
79. Choe S, Kim HS, and Lee S. Exploration of Superspreading Events in 2015 MERS-CoV Outbreak in Korea by Branching Process Models. International Journal of Environmental Research and Public Health 2020 Sep; 17:6137. DOI: 10.3390/ijerph17176137. Available from: https://pmc.ncbi.nlm.nih.gov/articles/PMC7504499/ [Accessed on: 2026 May 11]
80. Whittles LK, White PJ, and Didelot X. A dynamic power-law sexual network model of gonorrhoea outbreaks. en. PLOS Computational Biology 2019 Mar; 15:e1006748. DOI: 10.1371/journal.pcbi.1006748. Available from: https://journals.plos.org/ploscompbiol/article?id=10.1371/journal.pcbi.1006748 [Accessed on: 2026 May 20]
81. De Bellis A, Bizzotto A, Anagnostopoulos L, Kourentis L, Marziano V, Mouchtouri V, Merler S, and Guzzetta G. Mitigating norovirus spread on cruise ships: a model-based assessment of diagnostic

timing and isolation. Journal of Travel Medicine 2025 Oct; 32:taaf059. DOI: 10.1093/jtm/taaf059. Available from: https://doi.org/10.1093/jtm/taaf059 [Accessed on: 2026 May 1]

82. Kim S, Lee KS, Pak GD, Excler JL, Sahastrabuddhe S, Marks F, Kim JH, and Mogasale V. Spatial and Temporal Patterns of Typhoid and Paratyphoid Fever Outbreaks: A Worldwide Review, 1990–2018. Clinical Infectious Diseases: An Official Publication of the Infectious Diseases Society of America 2019 Nov; 69:S499–S509. DOI: 10.1093/cid/ciz705. Available from: https://pmc.ncbi.nlm.nih.gov/articles/PMC6821269/ [Accessed on: 2026 May 6]
83. Cho UJ, Cho S, Lee H, Kang SK, Kim BI, Nam Y, Achangwa C, Lim JS, Lee DH, and Ryu S. Transmission Dynamics and Parameters for Pertussis during School-Based Outbreak, South Korea, 2024. en. Emerging Infectious Diseases 2025 Jul; 31. DOI: 10.3201/eid3107.241643. Available from: https://wwwnc.cdc.gov/eid/article/31/7/24-1643_article [Accessed on: 2026 Apr 27]
84. Fraser C, Cummings DAT, Klinkenberg D, Burke DS, and Ferguson NM. Influenza Transmission in Households During the 1918 Pandemic. en. 2011. Available from: https://dx.doi.org/10.1093/aje/kwr122 [Accessed on: 2026 Apr 27]
85. Melsew YA, Gambhir M, Cheng AC, McBryde ES, Denholm JT, Tay EL, and Trauer JM. The role of super-spreading events in Mycobacterium tuberculosis transmission: evidence from contact tracing. en. BMC Infectious Diseases 2019 Mar; 19:244. DOI: 10.1186/s12879-019-3870-1. Available from: https://doi.org/10.1186/s12879-019-3870-1 [Accessed on: 2026 Apr 27]
86. Kappel D, Gifford H, Brackin A, Abdolrasouli A, Eyre DW, Jeffery K, Schlenz S, Aanensen DM, Brown CS, Borman A, Johnson E, Holmes A, Armstrong-James D, Fisher MC, and Rhodes J. Genomic epidemiology describes introduction and outbreaks of antifungal drug-resistant Candida auris. en. npj Antimicrobials and Resistance 2024 Sep; 2:26. DOI: 10.1038/s44259-024-00043-6. Available from: https://www.nature.com/articles/s44259-024-00043-6 [Accessed on: 2026 May 20]
87. Vink MA, Bootsma MCJ, and Wallinga J. Serial Intervals of Respiratory Infectious Diseases: A Systematic Review and Analysis. American Journal of Epidemiology 2014 Nov; 180:865–75. DOI: 10.1093/aje/kwu209. Available from: https://doi.org/10.1093/aje/kwu209 [Accessed on: 2026 May 18]
88. Simpson REH. STUDIES ON SHINGLES: IS THE VIRUS ORDINARY CHICKENPOX VIRUS ? The Lancet. Originally published as Volume 2, Issue 6852 1954 Dec; 264:1299–302. DOI: 10.1016/S0140-6736(54)90708-4. Available from: https://www.sciencedirect.com/science/article/pii/S0140673654907084 [Accessed on: 2026 May 18]
89. Azman AS, Rumunu J, Abubakar A, West H, Ciglenecki I, Helderman T, Wamala JF, Vázquez OdlR, Perea W, Sack DA, Legros D, Martin S, Lessler J, and Luquero FJ. Population-Level Effect of Cholera Vaccine on Displaced Populations, South Sudan, 2014. eng. Emerging Infectious Diseases 2016 Jun; 22:1067–70. DOI: 10.3201/eid2206.151592
90. Weil AA, Khan AI, Chowdhury F, LaRocque RC, Faruque A, Ryan ET, Calderwood SB, Qadri F, and Harris JB. Clinical Outcomes in Household Contacts of Patients with Cholera in Bangladesh. Clinical Infectious Diseases 2009 Nov; 49:1473–9. DOI: 10.1086/644779. Available from: https://doi.org/10.1086/644779 [Accessed on: 2026 May 20]
91. Xu X, Wu Y, Kummer AG, Zhao Y, Hu Z, Wang Y, Liu H, Ajelli M, and Yu H. Assessing changes in incubation period, serial interval, and generation time of SARS-CoV-2 variants of concern: a systematic review and meta-analysis. en. BMC Medicine 2023 Sep; 21:374. DOI: 10.1186/s12916-023-03070-8. Available from: https://doi.org/10.1186/s12916-023-03070-8 [Accessed on: 2026 May 21]
92. Richardson M, Elliman D, Maguire H, Simpson J, and Nicoll A. Evidence base of incubation periods, periods of infectiousness and exclusion policies for the control of communicable diseases in schools and preschools. en-US. The Pediatric Infectious Disease Journal 2001 Apr; 20:380. Available from: https://journals.lww.com/pidj/abstract/2001/04000/evidence_base_of_incubation_periods,_periods_of.4.aspx [Accessed on: 2026 May 18]
93. Vink A. The generation interval of infectious diseases. 2011 Mar. Available from: https://www.semanticscholar.org/paper/The-generation-interval-of-infectious-diseases.-Vink/9f40de5730dcb300eae69f9df2e50db332db8021 [Accessed on: 2026 May 19]
94. CDC. Chapter 4: Hepatitis B. en-us. 2026 Apr. Available from: https://www.cdc.gov/surv-manual/php/table-of-contents/chapter-4-hepatitis-b.html [Accessed on: 2026 May 26]
95. Ballegooijen WM van, Houdt R van, Bruisten SM, Boot HJ, Coutinho RA, and Wallinga J. Molecular Sequence Data of Hepatitis B Virus and Genetic Diversity After Vaccination. American Journal of

Epidemiology 2009 Dec; 170:1455–63. DOI: 10.1093/aje/kwp375. Available from: https://doi.org/10.1093/aje/kwp375 [Accessed on: 2026 May 18]
96. Heymann DL and American Public Health Association, eds. Control of communicable diseases manual: an official report of the American Public Health Association. eng. 18th ed. Washington, DC: The American Public Health Association, 2004
97. Azman AS, Ciglenecki I, Oeser C, Said B, Tedder RS, and Ijaz S. The incubation period of hepatitis E genotype 1: insights from pooled analyses of travellers. Epidemiology and Infection 2018 Sep; 146:1533–6. DOI: 10.1017/S0950268818001097. Available from: https://pmc.ncbi.nlm.nih.gov/articles/PMC6090710/ [Accessed on: 2026 May 20]
98. Zhuang H, Cao XY, Liu CB, and Wang GM. Epidemiology of hepatitis E in China. en. Gastroenterologia Japonica 1991 Jul; 26:135–8. DOI: 10.1007/BF02779283. Available from: https://doi.org/10.1007/BF02779283 [Accessed on: 2026 May 20]
99. Zhao S, Musa SS, Fu H, He D, and Qin J. Large-scale Lassa fever outbreaks in Nigeria: quantifying the association between disease reproduction number and local rainfall. en. Epidemiology & Infection 2020 Jan; 148:e4. DOI: 10.1017/S0950268819002267. Available from: https://www.cambridge.org/core/journals/epidemiology-and-infection/article/largescale-lassa-fever-outbreaks-in-nigeria-quantifying-the-association-between-disease-reproduction-number-and-local-rainfall/CDE6E940E9C14D3530E941D58C53A727 [Accessed on: 2026 May 26]
100. Klinkenberg D and Nishiura H. The correlation between infectivity and incubation period of measles, estimated from households with two cases. Journal of Theoretical Biology 2011 Sep; 284:52–60. DOI: 10.1016/j.jtbi.2011.06.015. Available from: https://www.sciencedirect.com/science/article/pii/S0022519311003146 [Accessed on: 2026 May 20]
101. Marziano V, Bella A, Menegale F, Manso MD, Petrone D, Palamara AT, Pezzotti P, Merler S, Filia A, and Poletti P. Estimating measles susceptibility and transmission patterns in Italy: an epidemiological assessment. English. The Lancet Infectious Diseases 2025 Dec; 25:1303–13. DOI: 10.1016/S1473-3099(25)00293-2. Available from: https://www.thelancet.com/journals/laninf/article/PIIS1473-3099(25)00293-2/fulltext [Accessed on: 2026 May 20]
102. Hoebe CJ, Melker H de, Spanjaard L, Dankert J, and Nagelkerke N. Space-Time Cluster Analysis of Invasive Meningococcal Disease. Emerging Infectious Diseases 2004 Sep; 10:1621–6. DOI: 10.3201/eid1009.030992. Available from: https://pmc.ncbi.nlm.nih.gov/articles/PMC3320315/ [Accessed on: 2026 May 18]
103. Park SH, Kim YS, Jung Y, Choi Sy, Cho NH, Jeong HW, Heo JY, Yoon JH, Lee J, Cheon S, and Sohn KM. Outbreaks of Middle East Respiratory Syndrome in Two Hospitals Initiated by a Single Patient in Daejeon, South Korea. Infection & Chemotherapy 2016 Jun; 48:99–107. DOI: 10.3947/ic.2016.48.2.99. Available from: https://pmc.ncbi.nlm.nih.gov/articles/PMC4945733/ [Accessed on: 2026 May 19]
104. Althobaity Y, Wu J, and Tildesley MJ. A comparative analysis of epidemiological characteristics of MERS-CoV and SARS-CoV-2 in Saudi Arabia. Infectious Disease Modelling 2022 Sep; 7:473–85. DOI: 10.1016/j.idm.2022.07.002. Available from: https://www.sciencedirect.com/science/article/pii/S2468042722000537 [Accessed on: 2026 May 19]
105. Assiri A, McGeer A, Perl TM, Price CS, Al Rabeeah AA, Cummings DA, Alabdullatif ZN, Assad M, Almulhim A, Makhdoom H, Madani H, Alhakeem R, Al-Tawfiq JA, Cotten M, Watson SJ, Kellam P, Zumla AI, and Memish ZA. Hospital Outbreak of Middle East Respiratory Syndrome Coronavirus. The New England journal of medicine 2013 Aug; 369:407–16. DOI: 10.1056/NEJMoa1306742. Available from: https://pmc.ncbi.nlm.nih.gov/articles/PMC4029105/ [Accessed on: 2026 May 19]
106. Kremer C, Nundu SS, Vakaniaki EH, Brosius I, Mukari G, Munganga P, Bangwen E, Tshomba JC, Mujula Y, De Vos E, Van Dijck C, Houben S, Gressani O, Kacita C, Mukadi-Bamuleka D, Wawina-Bokalanga T, Kinganda-Lusamaki E, Amuri-Aziza A, Lupola PM, Muyembe-Tamfum JJ, Mbala-Kingebeni P, Liesenborghs L, Hens N, and Torneri A. Epidemiological characteristics of monkeypox virus Clade Ib in the Democratic Republic of the Congo. en. Nature Communications 2025 Dec; 17:180. DOI: 10.1038/s41467-025-66875-6. Available from: https://www.nature.com/articles/s41467-025-66875-6 [Accessed on: 2026 May 20]
107. MEYER MB. AN EPIDEMIOLOGIC STUDY OF MUMPS; ITS SPREAD IN SCHOOLS AND FAMILIES. American Journal of Epidemiology 1962 Mar; 75:259–81. DOI: 10.1093/oxfordjournals.aje.a1

20248. Available from: https://doi.org/10.1093/oxfordjournals.aje.a120248 [Accessed on: 2026 May 18]
108. CDC. Clinical Overview of Typhoid Fever and Paratyphoid Fever. en-us. 2024 May. Available from: https://www.cdc.gov/typhoid-fever/hcp/clinical-overview/index.html [Accessed on: 2026 May 20]
109. Beest DE te, Henderson D, Maas NAT van der, Greeff SC de, Wallinga J, Mooi FR, and Boven M van. Estimation of the serial interval of pertussis in Dutch households. Epidemics 2014 Jun; 7:1–6. DOI: 10.1016/j.epidem.2014.02.001. Available from: https://www.sciencedirect.com/science/article/pii/S1755436514000048 [Accessed on: 2026 May 18]
110. Su Y, Dai R, Luo F, Zheng S, Hua C, He H, and Zhang H. Household transmission patterns and serial interval of pertussis in China. English. Journal of Infection 2024 Dec; 89. DOI: 10.1016/j.jinf.2024.106322. Available from: https://www.journalofinfection.com/article/S0163-4453(24)00256-1/fulltext [Accessed on: 2026 May 18]
111. Full article: Transmission patterns of pertussis in school-based clustered outbreaks: A retrospective analysis in Zhejiang Province, China, 2024. Available from: https://www.tandfonline.com/doi/full/10.1080/21645515.2026.2654305 [Accessed on: 2026 May 20]
112. Chan LYH, Morris SE, Stockwell MS, Bowman NM, Asturias E, Rao S, Lutrick K, Ellingson KD, Nguyen HQ, Maldonado Y, McLaren SH, Sano E, Biddle JE, Smith-Jeffcoat SE, Biggerstaff M, Rolfes MA, Talbot HK, Grijalva CG, Borchering RK, and Mellis AM. Estimating the generation time for influenza transmission using household data in the United States. Epidemics 2025 Mar; 50:100815. DOI: 10.1016/j.epidem.2025.100815. Available from: https://www.sciencedirect.com/science/article/pii/S1755436525000039 [Accessed on: 2026 May 18]
113. Carrat F, Vergu E, Ferguson NM, Lemaitre M, Cauchemez S, Leach S, and Valleron AJ. Time Lines of Infection and Disease in Human Influenza: A Review of Volunteer Challenge Studies. American Journal of Epidemiology 2008 Apr; 167:775–85. DOI: 10.1093/aje/kwm375. Available from: https://doi.org/10.1093/aje/kwm375 [Accessed on: 2026 May 18]
114. Cowling BJ, Fang VJ, Riley S, Malik Peiris JS, and Leung GM. Estimation of the Serial Interval of Influenza. en-US. Epidemiology 2009 May; 20:344. DOI: 10.1097/EDE.0b013e31819d1092. Available from: https://journals.lww.com/epidem/abstract/2009/05000/estimation_of_the_serial_interval_of_influenza.7.aspx [Accessed on: 2026 May 18]
115. Petrie JG, Ohmit SE, Cowling BJ, Johnson E, Cross RT, Malosh RE, Thompson MG, and Monto AS. Influenza Transmission in a Cohort of Households with Children: 2010-2011. en. PLOS ONE 2013 Sep; 8:e75339. DOI: 10.1371/journal.pone.0075339. Available from: https://journals.plos.org/plosone/article?id=10.1371/journal.pone.0075339 [Accessed on: 2026 May 18]
116. Cowling BJ, Chan KH, Fang VJ, Lau LLH, So HC, Fung ROP, Ma ESK, Kwong ASK, Chan CW, Tsui WWS, Ngai HY, Chu DWS, Lee PWY, Chiu MC, Leung GM, and Peiris JSM. Comparative Epidemiology of Pandemic and Seasonal Influenza A in Households. New England Journal of Medicine 2010 Jun; 362. _eprint: https://www.nejm.org/doi/pdf/10.1056/NEJMoa0911530:2175–84. DOI: 10.1056/NEJMoa0911530. Available from: https://www.nejm.org/doi/full/10.1056/NEJMoa0911530 [Accessed on: 2026 May 18]
117. Borgdorff MW, Sebek M, Geskus RB, Kremer K, Kalisvaart N, and Soolingen D van. The incubation period distribution of tuberculosis estimated with a molecular epidemiological approach. en. Available from: https://dx.doi.org/10.1093/ije/dyr058 [Accessed on: 2026 May 12]
118. Brooks-Pollock E, Becerra MC, Goldstein E, Cohen T, and Murray MB. Epidemiologic Inference From the Distribution of Tuberculosis Cases in Households in Lima, Peru. The Journal of Infectious Diseases 2011 Jun; 203:1582–9. DOI: 10.1093/infdis/jir162. Available from: https://pmc.ncbi.nlm.nih.gov/articles/PMC3096792/ [Accessed on: 2026 May 12]
119. Asbroek AH ten, Borgdorff MW, Nagelkerke NJ, Sebek MM, Devillé W, Embden JD van, and Soolingen D van. Estimation of serial interval and incubation period of tuberculosis using DNA fingerprinting. eng. The International Journal of Tuberculosis and Lung Disease: The Official Journal of the International Union Against Tuberculosis and Lung Disease 1999 May; 3:414–20
120. Ferreira FCdSL, Camacho LAB, and Villela DAM. Occurrence of yellow fever outbreaks in a partially vaccinated population: An analysis of the effective reproduction number. en. PLOS Neglected Tropical Diseases 2022 Sep; 16:e0010741. DOI: 10.1371/journal.pntd.0010741. Available from: https://journals.plos.org/plosntds/article?id=10.1371/journal.pntd.0010741 [Accessed on: 2026 May 18]

121. Wu JT, Peak CM, Leung GM, and Lipsitch M. Fractional dosing of yellow fever vaccine to extend supply: a modelling study. English. The Lancet 2016 Dec; 388:2904–11. DOI: 10.1016/S0140-6736(16)31838-4. Available from: https://www.thelancet.com/journals/lancet/article/PIIS0140-6736(16)31838-4/abstract [Accessed on: 2026 May 18]
122. Hoffmann S, White AE, McQueen RB, Ahn JW, Gunn-Sandell LB, and Scallan Walter EJ. Economic Burden of Foodborne Illnesses Acquired in the United States. EN. Foodborne Pathogens and Disease 2025 Jan; 22:4–14. DOI: 10.1089/fpd.2023.0157. Available from: https://journals.sagepub.com/action/showAbstract [Accessed on: 2026 May 20]
123. Bozkurt I, Sunbul M, Yilmaz H, Esen S, Leblebicioglu H, and Beeching NJ. Direct healthcare costs for patients hospitalized with Crimean-Congo haemorrhagic fever can be predicted by a clinical illness severity scoring system. Pathogens and Global Health 2016 Feb; 110:9–13. DOI: 10.1080/20477724.2015.1136130. Available from: https://pmc.ncbi.nlm.nih.gov/articles/PMC4870027/ [Accessed on: 2026 May 20]
124. Zhou F, Ortega-Sanchez IR, Guris D, Shefer A, Lieu T, and Seward JF. An Economic Analysis of the Universal Varicella Vaccination Program in the United States. The Journal of Infectious Diseases 2008 Mar; 197:S156–S164. DOI: 10.1086/522135. Available from: https://doi.org/10.1086/522135 [Accessed on: 2026 May 20]
125. CDC. Cholera in the United States. en-us. 2025 Sep. Available from: https://www.cdc.gov/cholera/about/about-cholera-in-the-united-states.html [Accessed on: 2026 May 20]
126. Kapinos KA, Peters RM, Murphy RE, Hohmann SF, Podichetty A, and Greenberg RS. Inpatient Costs of Treating Patients With COVID-19. JAMA Network Open 2024 Jan; 7. Available from: https://jamanetwork.com/journals/jamanetworkopen/fullarticle/2813457 [Accessed on: 2026 May 20]
127. IHME. Global Burden of Disease (GBD). en. Available from: https://www.healthdata.org/research-analysis/gbd [Accessed on: 2026 May 26]
128. Yu W, Guo Y, Zhang S, Kong Y, Shen Z, and Zhang J. Proportion of asymptomatic infection and nonsevere disease caused by SARS-CoV-2 Omicron variant: A systematic review and analysis. Journal of Medical Virology 2022 Aug :10.1002/jmv.28066. DOI: 10.1002/jmv.28066. Available from: https://pmc.ncbi.nlm.nih.gov/articles/PMC9538850/ [Accessed on: 2026 May 26]
129. Ekwueme DU, Strebel PM, Hadler SC, Meltzer MI, Allen JW, and Livengood JR. Economic Evaluation of Use of Diphtheria, Tetanus, and Acellular Pertussis Vaccine or Diphtheria, Tetanus, and Whole-Cell Pertussis Vaccine in the United States, 1997. Archives of Pediatrics & Adolescent Medicine 2000 Aug; 154:797–803. DOI: 10.1001/archpedi.154.8.797. Available from: https://doi.org/10.1001/archpedi.154.8.797 [Accessed on: 2026 May 20]
130. Reich NG, Lessler J, Varma JK, and Vora NM. Quantifying the Risk and Cost of Active Monitoring for Infectious Diseases. Scientific Reports 2018 Jan; 8:1093. DOI: 10.1038/s41598-018-19406-x. Available from: https://pmc.ncbi.nlm.nih.gov/articles/PMC5773605/ [Accessed on: 2026 May 26]
131. Fox M. Cost to Treat Ebola: $1 Million For Two Patients. en. 2014 Nov. Available from: https://www.nbcnews.com/storyline/ebola-virus-outbreak/cost-treat-ebola-1-million-two-patients-n250986 [Accessed on: 2026 May 26]
132. Nelson NP, Easterbrook PJ, and McMahon BJ. Epidemiology of Hepatitis B Virus Infection and Impact of Vaccination on Disease. Clinics in liver disease 2016 Nov; 20:607–28. DOI: 10.1016/j.cld.2016.06.006. Available from: https://pmc.ncbi.nlm.nih.gov/articles/PMC5582972/ [Accessed on: 2026 May 20]
133. Nguyen MH, Ozbay AB, Liou I, Meyer N, Gordon SC, Dusheiko G, and Lim JK. Healthcare resource utilization and costs by disease severity in an insured national sample of US patients with chronic hepatitis B. English. Journal of Hepatology 2019 Jan; 70:24–32. DOI: 10.1016/j.jhep.2018.09.021. Available from: https://www.journal-of-hepatology.eu/article/S0168-8278(18)32440-1/fulltext [Accessed on: 2026 May 20]
134. Talbird SE, Anderson SA, Nossov M, Beattie N, Rak AT, and Diaz-Mitoma F. Cost-effectiveness of a 3-antigen versus single-antigen vaccine for the prevention of hepatitis B in adults in the United States. Vaccine 2023 May; 41:3506–17. DOI: 10.1016/j.vaccine.2023.04.022. Available from: https://www.sciencedirect.com/science/article/pii/S0264410X23004140 [Accessed on: 2026 May 20]

135. Sriudomporn S and Patenaude B. Quantifying the Cost of Measles Outbreak in the U.S. and How Costs Scale with Outbreak Size. en. ISSN: 3067-2007 Pages: 2025.10.24.25338724. 2025 Oct. DOI: 10.1101/2025.10.24.25338724. Available from: https://www.medrxiv.org/content/10.1101/2025.10.24.25338724v2 [Accessed on: 2026 May 20]
136. Anonychuk A, Woo G, Vyse A, Demarteau N, and Tricco AC. The Cost and Public Health Burden of Invasive Meningococcal Disease Outbreaks: A Systematic Review. Pharmacoeconomics 2013; 31:563–76. DOI: 10.1007/s40273-013-0057-2. Available from: https://pmc.ncbi.nlm.nih.gov/articles/PMC3691489/ [Accessed on: 2026 May 20]
137. Herrera-Restrepo O, Kwiatkowska M, Huse S, Kocaata Z, and Ganz ML. Retrospective analysis of health and economic burden among commercially-insured individuals diagnosed with invasive meningococcal disease in the United States. Human Vaccines & Immunotherapeutics. 20:2436039. DOI: 10.1080/21645515.2024.2436039. Available from: https://pmc.ncbi.nlm.nih.gov/articles/PMC11651274/ [Accessed on: 2026 May 20]
138. Zhang XS, Niyomsri S, Mandal S, Mohammed H, Mindlin M, Dugbazah B, Adjei S, Owoseni B, Charlett A, I'Anson J, Sugars E, Kliner M, Mannes T, Jewitt E, Gilbert L, Moazam S, Dewsnap C, Phillips D, Amirthalingam G, Ramsay ME, Vickerman P, and Walker JG. Cost-effectiveness of vaccination strategies to control future mpox outbreaks in England: a modelling study. eng. The Lancet Regional Health. Europe 2025 Aug; 55:101364. DOI: 10.1016/j.lanepe.2025.101364
139. Pike J, Schwartz S, Kay M, Perez-Osorio A, Marin M, Jenkins M, Routh J, Duchin J, DeBolt C, and Zhou F. Cost of responding to the 2017 University of Washington Mumps Outbreak: A Prospective Analysis. Journal of public health management and practice : JPHMP 2020; 26:116–23. DOI: 10.1097/PHH.0000000000000957. Available from: https://pmc.ncbi.nlm.nih.gov/articles/PMC6733677/ [Accessed on: 2026 May 20]
140. Tucker C. As norovirus outbreaks grow, health departments respond: Millions infected each year with virus. en. 2013 Mar. Available from: https://www.thenationshealth.org/content/43/2/1.3 [Accessed on: 2026 May 20]
141. Wang J, Gao Z, Yang Zr, Liu K, and Zhang H. Global prevalence of asymptomatic norovirus infection in outbreaks: a systematic review and meta-analysis. BMC Infectious Diseases 2023 Sep; 23:595. DOI: 10.1186/s12879-023-08519-y. Available from: https://pmc.ncbi.nlm.nih.gov/articles/PMC10496210/ [Accessed on: 2026 May 26]
142. Roo AMd, Vondeling GT, Boer M, Murray K, and Postma MJ. The global health and economic burden of chikungunya from 2011 to 2020: a model-driven analysis on the impact of an emerging vector-borne disease. en. BMJ Global Health 2024 Dec; 9. DOI: 10.1136/bmjgh-2024-016648. Available from: https://gh.bmj.com/content/9/12/e016648 [Accessed on: 2026 May 27]
143. CDC. 2025 Provisional Pertussis Surveillance Report. Tech. rep. 2026 Jan. Available from: https://www.cdc.gov/pertussis/media/pdfs/2026/02/363295-A_FS_PertussisSurveillanceReport_011626_508pass.pdf
144. O'Brien JA and Caro JJ. Hospitalization for pertussis: profiles and case costs by age. BMC Infectious Diseases 2005 Jul; 5:57. DOI: 10.1186/1471-2334-5-57. Available from: https://pmc.ncbi.nlm.nih.gov/articles/PMC1184075/ [Accessed on: 2026 May 20]
145. McGarry LJ, Krishnarajah G, Hill G, Skornicki M, Pruttivarasin N, Masseria C, Arondekar B, Pelton SI, and Weinstein MC. Cost-Effectiveness Analysis of Tdap in the Prevention of Pertussis in the Elderly. en. PLOS ONE 2013 Sep; 8:e67260. DOI: 10.1371/journal.pone.0067260. Available from: https://journals.plos.org/plosone/article?id=10.1371/journal.pone.0067260 [Accessed on: 2026 May 26]
146. CDC. Local Health Department Costs Associated with Response to a School-Based Pertussis Outbreak — Omaha, Nebraska, September–November 2008. eng. 2011 Jan. Available from: https://www.cdc.gov/mmwr/preview/mmwrhtml/mm6001a2.htm [Accessed on: 2026 May 20]
147. Huang SS, Johnson KM, Ray GT, Wroe P, Lieu TA, Moore MR, Zell ER, Linder JA, Grijalva CG, Metlay JP, and Finkelstein JA. Healthcare utilization and cost of pneumococcal disease in the United States. Vaccine 2011 Apr; 29:3398–412. DOI: 10.1016/j.vaccine.2011.02.088. Available from: https://www.sciencedirect.com/science/article/pii/S0264410X11003367 [Accessed on: 2026 May 20]
148. Du Y, Yan R, Wu X, Zhang X, Chen C, Jiang D, Yang M, Cao K, Chen M, You Y, Zhou W, Chen D, Xu G, and Yang S. Global burden and trends of respiratory syncytial virus infection across different

age groups from 1990 to 2019: A systematic analysis of the Global Burden of Disease 2019 Study. International Journal of Infectious Diseases 2023 Oct; 135:70–6. DOI: 10.1016/j.ijid.2023.08.008. Available from: https://www.sciencedirect.com/science/article/pii/S1201971223006938 [Accessed on: 2026 May 20]

149. CDC. Preliminary Estimates of RSV Burden. en-us. 2026 Feb. Available from: https://www.cdc.gov/rsv/php/surveillance/burden-estimates.html [Accessed on: 2026 May 20]

150. Carrico J, Hicks KA, Wilson E, Panozzo CA, and Ghaswalla P. The Annual Economic Burden of Respiratory Syncytial Virus in Adults in the United States. The Journal of Infectious Diseases 2023 Dec; 230:e342–e352. DOI: 10.1093/infdis/jiad559. Available from: https://pmc.ncbi.nlm.nih.gov/articles/PMC11326840/ [Accessed on: 2026 May 20]

151. Hall C, Geiman J, Biggar R, Kotok D, Hogan P, and Jr DG. Respiratory syncytial virus infections within families. N Engl J Med 1976; 294:414–9. DOI: 10.1056/NEJM197602192940803. Available from: https://pubmed.ncbi.nlm.nih.gov/173995/

152. Cassini A, Colzani E, Pini A, Mangen MJJ, Plass D, McDonald SA, Maringhini G, Lier A van, Haagsma JA, Havelaar AH, Kramarz P, and Kretzschmar ME. Impact of infectious diseases on population health using incidence-based disability-adjusted life years (DALYs): results from the Burden of Communicable Diseases in Europe study, European Union and European Economic Area countries, 2009 to 2013. Eurosurveillance 2018 Apr; 23:17–454. DOI: 10.2807/1560-7917.ES.2018.23.16.17-00454. Available from: https://pmc.ncbi.nlm.nih.gov/articles/PMC5915974/ [Accessed on: 2026 May 20]

153. Thompson KM and Odahowski CL. The Costs and Valuation of Health Impacts of Measles and Rubella Risk Management Policies. en. Risk Analysis 2015 Aug; 36. DOI: doi.org/10.1111/risa.12459. Available from: https://onlinelibrary.wiley.com/doi/abs/10.1111/risa.12459 [Accessed on: 2026 May 20]

154. Iezzoni LI, Yu J, Wint AJ, Smeltzer SC, and Ecker JL. Prevalence of Current Pregnancy Among U.S. Women with and without Chronic Physical Disabilities. Medical care 2013 Jun; 51:555–62. DOI: 10.1097/MLR.0b013e318290218d. Available from: https://pmc.ncbi.nlm.nih.gov/articles/PMC3733491/ [Accessed on: 2026 May 26]

155. U.S. Census Bureau. U.S. and World Population Clock. 2026 May. Available from: https://www.census.gov/popclock/ [Accessed on: 2026 May 26]

156. Popovian R and Winegarden W. Influenza's Economic Burden and the Impact of Adult Vaccination. en. ISSN: 3067-2007 Pages: 2025.12.23.25342685. 2025 Dec. DOI: 10.64898/2025.12.23.25342685. Available from: https://www.medrxiv.org/content/10.64898/2025.12.23.25342685v1 [Accessed on: 2026 May 20]

157. Furuya-Kanamori L, Cox M, Milinovich GJ, Magalhaes RJS, Mackay IM, and Yakob L. Heterogeneous and Dynamic Prevalence of Asymptomatic Influenza Virus Infections. Emerging Infectious Diseases 2016 Jun; 22:1052–6. DOI: 10.3201/eid2206.151080. Available from: https://pmc.ncbi.nlm.nih.gov/articles/PMC4880086/ [Accessed on: 2026 May 21]

158. Castro KG, Marks SM, Chen MP, Hill AN, Becerra JE, Miramontes R, Winston CA, Navin TR, Pratt RH, Young KH, and LoBue PA. Estimating tuberculosis cases and their economic costs averted in the United States over the past two decades. The international journal of tuberculosis and lung disease : the official journal of the International Union against Tuberculosis and Lung Disease 2016 Jul; 20:926–33. DOI: 10.5588/ijtld.15.1001. Available from: https://pmc.ncbi.nlm.nih.gov/articles/PMC4992985/ [Accessed on: 2026 May 20]

159. Shrestha S, Cilloni L, Asay GRB, Kammerer JS, Raz K, Shaw T, Cilnis M, Wortham J, Marks SM, and Dowdy D. Model-Based Analysis of Impact, Costs, and Cost-effectiveness of Tuberculosis Outbreak Investigations, United States. en-us. Emerging Infectious Diseases 2025 Mar; 31. DOI: 10.3201/eid3103.240633. Available from: https://wwwnc.cdc.gov/eid/article/31/3/24-0633_article [Accessed on: 2026 May 26]

160. Emery JC, Dodd PJ, Banu S, Frascella B, Garden FL, Horton KC, Hossain S, Law I, Leth F van, Marks GB, Nguyen HB, Nguyen HV, Onozaki I, Quelapio MID, Richards AS, Shaikh N, Tiemersma EW, White RG, Zaman K, Cobelens F, and Houben RM. Estimating the contribution of subclinical tuberculosis disease to transmission: An individual patient data analysis from prevalence surveys. eLife 2023 Dec; 12. Ed. by Kana BD:e82469. DOI: 10.7554/eLife.82469. Available from: https://doi.org/10.7554/eLife.82469 [Accessed on: 2026 May 20]

161. Feng H, Liang H, Xu H, Huang S, Li J, Wen W, Chen Y, and Zhou F. The hidden burden: a regional study revealing the substantial burden and distinct epidemiology of asymptomatic pulmonary tuberculosis in Southern China. en. BMC Public Health 2026 Mar; 26:1129. DOI: 10.1186/s12889-026-26848-w. Available from: https://doi.org/10.1186/s12889-026-26848-w [Accessed on: 2026 May 26]
162. Shepard DS, Coudeville L, Halasa YA, Zambrano B, and Dayan GH. Economic Impact of Dengue Illness in the Americas. The American Journal of Tropical Medicine and Hygiene 2011 Feb; 84:200–7. DOI: 10.4269/ajtmh.2011.10-0503. Available from: https://pmc.ncbi.nlm.nih.gov/articles/PMC3029168/ [Accessed on: 2026 May 20]
163. Bureau of Labor Statistics. CPI Inflation Calculator. en. Available from: https://www.bls.gov/data/inflation_calculator.htm [Accessed on: 2026 May 20]
164. IRS. Yearly average currency exchange rates. en. 2026 Feb. Available from: https://www.irs.gov/individuals/international-taxpayers/yearly-average-currency-exchange-rates [Accessed on: 2026 May 26]
