## Supplemental Table 1 for "Why epidemic risk at the 2026 World Cup may not be what you think"

### Summary of WC threat-assessment decisions

Generated May 27, 2026

Supplemental table S1 to "Why epidemic risk at the 2026 World Cup may not be what you think."

| Pathogen | Decision & details |
|---|---|
| **PRIORITY PATHOGENS - Seasonal Respiratory Viruses** | |
| **COVID-19** | **Inclusion Criteria: Endemic seasonal respiratory virus with potential for significant importations for countries where disease is in season and elevated spread at World Cup related activities.**<br>• *Notes and references:* Excess importations are estimated based on CDC estimates of illness for 2024-25 season and assuming 40% symptomatic proportion. Incidence assigned to epi weeks using COVID-NET data. Assign June incidence to all countries in Europe, North Africa, Middle East, North America (excl. Panama and Curacao) and Japan. Assign peak incidence to South America (incl. Panama and Curacao), Sub-Saharan Africa, and Australia. Assume 10 day infectious period per CDC.[1] [23] [4] Transmission in June assumed to follow latest $R_t$ estimates from CDC[5].<br>• *Expected Excess Importations:* 1,920<br>• *Local Presence at World Cup Related Activites:* 17,388<br>• *Additional Infections due to Transmission (per imported case):* 4.9 (World Cup), 3.1 (normal)<br> ◦ simulation parameters $R_t = 1$ (community), $R_t = 1.2$ (World Cup), $k = 0.41$, $g = 2.99$, susceptibility N/A since $R_t$ based simulation<br>• *Total Additional Infections:* 44,300<br>• *Cost per case:* $488<br>• *Expected Impact:* $16,200,000 |
| **RSV** | **Inclusion Criteria: Endemic seasonal respiratory virus with potential for significant importations for countries where disease is in season and elevated spread at World Cup related activities.**<br>• *Notes and references:* June incidence for each country determined by applying proportion of flu cases that occur in June for each country as reported to FluNet[6] and assuming RSV will follow similar patterns (apportioned based on annual US incidence rates). US incidence based on seasonal allocation of a 20% attack rate (in line with estimates for adults)[7]. Transmission in June assumed to follow latest $R_t$ estimates from CDC[5].<br>• *Expected Excess Importations:* 1,174<br>• *Local Presence at World Cup Related Activities:* 217<br>• *Additional Infections due to Transmission (per imported case):* 1.1 (World Cup), 0.9 (normal)<br> ◦ simulation parameters $R_t = 0.78$ (community), $R_t = 0.94$ (World Cup), $k = 0.94$, $g = 8.8$, susceptibility N/A since $R_t$ based simulation<br>• *Total Additional Infections:* 2,564<br>• *Cost per case:* $688<br>• *Expected Impact:* $1,763,707 |
| **Seasonal Influenza (A/B)** | **Inclusion Criteria: Endemic seasonal respiratory virus with potential for significant importations for countries where disease is in season and elevated spread at World Cup related activities.**<br>• *Notes and references:* June incidence for each country determined by applying proportion of flu cases that occur in June for each country as reported to FluNet[6] to annual US incidence. Specifically, 51 million sympathetic illnesses in 2024-25 season. Assume 84% of all illnesses are symptomatic. Using NHSN |

hospitalization data estimate that ~1% of infections occur in June[8 91]. Implies incidence of 174.8/100k in June. US incidence based on seasonal allocation of a 20% attack rate (in line with estimates for adults)[7]. Transmission in June assumed to follow latest $R_t$ estimates from CDC[5].

- *Expected Excess Importations:* 946
- *Local Presence at World Cup Related Activites:* 7,187
- *Additional Infections due to Transmission (per imported case):* 3.8 (World Cup), 2.5 (normal)
    - simulation parameters $R_t = 0.91$ (community), $R_t = 1.1$ (World Cup), $k = 0.94$, $g = 3.1$, susceptibility N/A since $R_t$ based simulation
- *Total Additional Infections:* 14,491
- *Cost per case:* $1,230
- *Expected Impact:* $21,943,257

#### PRIORITY PATHOGENS - Mosquito-Borne Pathogens

**Chikungunya**

**Inclusion Criteria: Non-endemic pathogen with potential to establish endemic transmission in some World Cup host locations.**

- *Notes and references:* Importation analysis suggests an expected 4.7 importations of Chikungunya over the course of the tournament. Given documented cases of US transmission[10] and year round presence of transmission conditions in some World Cup host cities, established endemic transmission is plausible.

**Dengue**

**Inclusion Criteria: Non-endemic pathogen with potential to establish endemic transmission in some World Cup host locations.**

- *Notes and references:* Importation analysis suggests an expected 3.4 importations of dengue over the course of the tournament. Given documented cases of US transmission[11 1213] and year round presence of transmission conditions in some World Cup host cities, established endemic transmission is plausible.

**Malaria**

**Inclusion Criteria: Non-endemic pathogen with potential to establish endemic transmission in some World Cup host locations.**

- *Notes and references:* Malaria importations are common, approximately 2000 per year[14]. Given documented cases of US transmission[14] and year round presence of transmission conditions in some World Cup host cities, established endemic transmission is plausible, if unlikely.

**Zika**

**Inclusion Criteria: Non-endemic pathogen with potential to establish endemic transmission in some World Cup host locations.**

- *Notes and references:* Zika importations have occurred at low rates, but largely from World Cup countries. Given documented cases of US transmission[15] and year round presence of transmission conditions in some World Cup host cities, established endemic transmission is plausible.

#### PRIORITY PATHOGENS - High Importation Risk Pathogens

**Hepatitis B**

**Inclusion criteria: Endemic pathogen with no increased transmission risk at World Cup related activities but importations likely to increase local prevalence by more than 5%.**

- *Notes and references: Estimated chronic HBV prevalence in US adults (2023): 640,000 cases Chronic HBV prevalence among US adults as a proportion = 640,000/299,300,000 = 0.002 Estimated number of prevalent HBV cases, WC cities = 0.002*84.6 million = 169,200 Number of imported cases needed to meet 5% local prevalence threshold: 0.05 * 169,200 = 8,460 imported cases Global HBV prevalence (2024) = 2.9% Estimated international WC travelers = 1 million Total importations = 0.029 * 640,000 = 18,400 imported cases Key references (US & global prevalence):[16 17]. Susceptibility estimates based on[18] **Note that none of the per case impact will occur during the World Cup itself.** Estimated cost per case of $25742 calculated from estimated cost of acute and chronic cases[19,20] combined with the assumption that 10% of acute cases will become chronic[21].

[NEED TO UPDATE THIS] *WC Per case outbreak size:* Different than others because of chronic infections not counting unless become hepatic.; General calculation…needs to be updated: To make an attempt using AI summary numbers w/o interrogating up for a moment, it looks like chronic cases cause 4.9-7 secondary cases over the course of their illness. Let's say most people are living 40 years with chronic HBV infection, that would give us something like 17/(40 * 365) = 0.0012 expected infections at the World Cup…assuming complete susceptibility. But vax rates among US adults are around 35%, so we should see 0.0012 x 0.65 = 0.00076 secondary infections per chronic import…or 18,386 * 0.00076 = 13.9 infections causes from the imported cases

- *Expected Excess Importations:* 18,400
- *Additional Infections due to Transmission (per imported case):* 0.00076
  - simulation parameters $R_0 = 0.00076$, $k = 0.23$, $g = 85$, 73% susceptibility
- *Total Additional Infections:* 10
- *Cost per Case:* $25,742
- *Expected Impact:* $102,479

---

**Tuberculosis (TB)**

**Inclusion criteria: Endemic pathogen with some increased transmission risk at World Cup related activities and importations likely to increase active disease by measurable levels.**

- *Notes and references:* Results reflect *active* TB cases only. Data from 2014 on direct medical cost of non-MDR TB ($24110) + societal cost including productivity lost ($62400)[22] Adjusted for inflation Public health response cost?[22]; Actually for this paper the societal cost includes the direct costs so the direct + indirect costs is $62,400. Extracted a public health estimate of $11,320 from here:[23] Total is $73,720[23] Transmission increases 1at World Cup appear possible
- *Expected Excess Importations:* 14.8
- *Local Presence at World Cup Related Activities:* 9
- *Additional Infections due to Transmission (per imported case):* 1.8 (World Cup), 1.5 (normal)
  - simulation parameters $R_0 = 1.5$ (community), $R_0 = 1.8$ (World Cup), $k = 0.16$, $g = 653$, 100% susceptibility
- *Total Additional Infections:* 44
- *Cost per Case:* $73,720
- *Expected Impact:* $3,316,711

---

#### PRIORITY PATHOGENS - Locally Present with Excess Transmission Risk

**Measles**

**Inclusion criteria: Locally present pathogen with increased transmission risk at World Cup related activities and potential impact from importations.**

- *Notes and references:* Measles outbreaks are known to occur at sporting events and other larger outdoor gatherings[24 25]. The exceptionally high transmissibility of measles means that large transmission events are possible even at relatively short gatherings [CITE] Per case costs based on[26]*
- *Expected Excess Importations:* 0.212
- *Local Presence at World Cup Related Activities:* 1.3
- *Additional Infections due to Transmission (per imported case):* 2.1 (World Cup), 1.7 (normal)
  - simulation parameters $R_0 = 14.8$ (community), $R_0 = 17.7$ (World Cup), $k = 0.63$, $g = 1.6$, 11.9% susceptiblity
- *Total Additional Infections:* 1.1
- *Cost per Case:* $43,000
- *Expected Impact:* $50,643

---

**Norovirus**

**Inclusion criteria: Locally present pathogen with increased transmission risk at World Cup related activities and potential impact from importations.**

- *Notes and references: Compared to other enteric viruses, there is more evidence to support elevated transmission at sporting events for norovirus. Carriage in staff and recovered from cases occurred 2018*

*Olympics and the Qatar World Cup. When factors associated with outbreaks are analyzed, increased $R_0$ is associated with the winter months and university/school settings[27], but little else. Food service as the setting is related substantially increased outbreak size [2827]; however norovirus outbreaks much less common in the summer: [29] Importations were calculated assuming similar incidence rates in travelers as the US population. Local prevalence based on seasonal incidence rates applied to expected numbers of participants in World Cup activities, divided by the average outbreak size (to adjust for clustering). Cost calculations based on Medical and productivity costs[30] Response costs based on an outbreak of 307 which cost $10,000 total.[31] Though overall projected attack rates seem large, they are relatively small increases at the per-capital basis compared to the already high projected norivirus incidence.*

- *Expected Excess Importations:* 60.3
- *Local Presence at World Cup Related Activities:* 222 (indepdent introductions)
- *Additional Infections due to Transmission (per imported case):* 2.3 (World Cup), 1.8 (normal)
  - simulation parameters $R_0 = 2.45$ (community), $R_0 = 3.25$ (World Cup), $k = 0.42$, $g = 3.77$, 80% susceptiblity
- *Total Additional Infections:* 6,568
- *Cost per Case:* $496
- *Expected Impact:* $4,177,576

#### PRIORITY PATHOGENS - Critical Scenarios and High Consequence, Low Proabability Pathogens

**Hantavirus (Andes Virus) [scenario]**

**Inclusion Criteria: High impact pathogen without ability to establish local transmission ($R_0 < 1$) and uncertain risk landscape due to recent events.**

- *Notes and references:* Based on uncertainty induced by the recent cruise ship outbreak, we analyzed a scenario assuming one introduction of Andes Hantavirus to the US during the World Cup, noting that ***this scenario remains highly unlikely***. Costs assumed to be $500,000 cases based on the low end of costs for other rare, high consequence pathogen responses (e.g., Ebola).
- *Expected Excess Importations:* 1 (hypothetical scenario)
- *Additional Infections due to Transmission (per cases):* 2.12
  - simulation parameters $R_0 = 2.12$, $k = 0.16$, $g = 23$, 100% susceptibility
- *Total Additional Infections:* 3.12
- *Cost per case:* $500,000 (scenario assumption)
- *Expected Impact:* $1,562,000

**Lassa Fever**

**Inclusion Criteria: A non-endemic pathogen with no ability to establish local transmission (no documented extended transmission in high income countries) and a very low probability of importation, but a high enough per case impact to warrant inclusion on the list.**

- *Notes and references:* Lassa fever is far more common than Ebola and Marburg. Importation estimates based on a 20% increase over historic rates (9 cases imported into the US since 1969[32]). Per introduction costs assumed to be the same as Ebola and based on introductions in 2014[33].
- *Expected Excess Importations:* 0.00337
- *Additional Infections due to Transmission (per cases):* 2.5
  - simulation parameters $R_0 = 1.6$, $k = 0.14$, $g = 7.8$, 100% susceptibility
- *Total Additional Infections:* 0.012
- *Cost per case:* $4,300,000
- *Expected Impact:* $50,177

**Novel Resp. Virus (including novel influenza)**

**Inclusion Criteria: Hypothetical high impact pathogen with potential ability to establish local transmission. Low probability event with little details, that should always be on watch list.**

- *Notes and references:* This is the proverbial "disease X" and no details are available. Based on simulations, a single introduction with a novel respiratory virus with pandemic influenza like transmission characteristics[34] would be expected to cause about 18 additional cases.

| Excluded pathogens | |
|---|---|
| *C. perfringens* | **Exclusion criteria: Endemic pathogen with no increased transmission risk at World Cup related activities and increased local prevalence due to importations of less than 5%.**<br>• *Notes and references: 1 million people a year get this in the US. Importations will be insignificant.*[35]. |
| **Anaplasmosis** | **Exclusion criteria: Endemic pathogen with no increased transmission risk at World Cup related activities and increased local prevalence due to importations of less than 5%.**<br>• *Notes and references: On average there are ~3,000 cases in just Jun and July alone. Unlikely that importation will be significant.* [36]*; This meta-review found that incidence was highest in North America. Although Africa was not included in the analysis, only ~4% of WC travelers are expected to come from Africa.*[37] *[2]* |
| **Anthrax** | **Exclusion criteria: Endemic pathogen with no increased transmission risk at World Cup related activities and increased local prevalence due to importations of less than 5%.**<br>• *Notes and references: Extremely rare in the US recently. Only 9 cases since 2006 (as of 2025).*[38] *We can find no documented cases of someone in the US having anthrax after travel or a traveler coming with it (importations are associated with animal hides).*[38] evalence |
| **Babesiosis** | **Exclusion criteria: Endemic pathogen with no increased transmission risk at World Cup related activities and increased local prevalence due to importations of less than 5%.**<br>• *Notes and references:* *~3,500 cases in 2023[39]. A meta-review looked at prevalence by continent. The only continent with more than 6.5x the prevalence of North America was Australia (and this was based on one study looking at a population of people referred for suspected Lyme Disease). The weighted average prevalence among expected WC visitors is 2.8x North America prevalence. Therefore excluding on the basis of the 6.5 rule.[40] |
| **Bacillus cereus** | **Exclusion criteria: Endemic pathogen with no increased transmission risk at World Cup related activities and increased local prevalence due to importations of less than 5%.**<br>• *Notes and references: Naturally ubiquitous, so no significant importation risk.*[41] |
| **Brucellosis** | **Exclusion criteria: Endemic pathogen with no increased transmission risk at World Cup related activities and increased local prevalence due to importations of less than 5%.**<br>• *Notes and references: 80-200 people in the US get Brucellosis a year.* [42] *Global estimated incidence is 2.1 million cases per year.*[43] *2-4 week infection period*[44] *with the vast majority of cases are in Africa.*[42] [43][44]*. Assuming incidence rates among World Cup travelers similar to regional rates (almost certainly an overestimate) gives an estimate of 1.1 imports per year. [REFERENCE SPECIAL TABLE FOR MATH]* local prevalence |
| **CPO (Carbapenemase)** | **Exclusion criteria: Endemic pathogen with no increased transmission risk at World Cup related activities and increased local prevalence due to importations of less than 5%.**<br>• *Notes and references: This is globally distributed, and very common in the United States with most cases being healthcare related.*[45] |
| **Candida auris** | **Exclusion criteria: Endemic pathogen with no increased transmission risk at World Cup related activities and increased local prevalence due to importations of less than 5%.**<br>• *Notes and references: Primarily hospital associated with minimal carriage in healthy individuals.*[46] |
| **Chickenpox** | **Exclusion criteria: Endemic pathogen with increased transmission risk at World Cup related activities but not enough anticipated impact to warrant inclusion.**<br>• *Notes and references:* Susceptibility based on current coverage of chickenpox vaccine[47]. Imports calculated as a multiple on measles imports based on ratio of global incidence, 140 million cases per year for chickenpox[48], versus 10.3 million per year for measles[49]. |

- *Expected Excess Importations:* 3.0
- *Local Presence at World Cup Related Activites:* 144
- *Additional Infections due to Transmission (per imported case):* 0.75 (World Cup), 0.62 (normal)
    - simulation parameters $R_0 = 6.2$ (community), $R_0 = 7.4$ (World Cup), $k = 0.93$, $g = 14$, 10% susceptibility
- *Total Additional Infections:* 23.5
- *Cost per Case:* $730
- *Expected Impact:* $17,100

---

**Cholera**

**Exclusion Criteria: A non-endemic pathogen with no ability to establish local transmission (water and sanitation conditions) and minimal impact from imports.**

- *Notes and references: Endemic cholera is absent from countries with adequate water and sanitation infrastructure. Even with highly pessimistic assumptions about secondary cases at the World Cup (assuming endemic country transmission rates) expected impact is low.*
- *Expected Excess Importations:* 0.0546
- *Additional Infections due to Transmission (per cases):* 8.15
    - simulation parameters $R_0 = 1.65$, $k = 4.5$, $g = 4$, 100% susceptibility
- *Total Additional Infections:* 0.445
- *Cost per case:* $1,190[50] [51]
- *Expected Impact:* $530

---

**Crimean-Congo Hemorrhagic Fever (CCHF)**

**Exclusion Criteria: A non-endemic pathogen with no ability to establish local transmission (absent vector) and minimal impact from imports.**

- *Notes and references: Vector is not present in the US[52]. Importations calculated based on excess World Cup travel and rates from[53], under the pessimistic assumption 25% of case occur in June. Per case cost[54] adjusted for inflation.*
- *Expected Excess Importations:* 0.13
- *Additional Infections due to Transmission (per case):* 0
- *Total Additional Infections:* 0.13
- *Cost per case:* $1,830
- *Expected Impact:* $238

---

**Cryptosporidiosis**

**Exclusion criteria: Endemic pathogen with no increased transmission risk at World Cup related activities and increased local prevalence due to importations of less than 5%.**

- *Notes and references: Over 823,000 cases per year in US; US is in highest category for prevalence among general population (Figure 3) in[55]*

---

**Cyclosporiasis**

**Exclusion criteria: Endemic pathogen with no increased transmission risk at World Cup related activities and increased local prevalence due to importations of less than 5%.**

- *Notes and references: About 20% of cases from 2015-2019 reported international travel…about 16 per year. Even a 20% increase in international importations (pessimistic given WC countries) would only increase cases by 20%* 20% = 4% increase in total cases.[56].*

---

**Diphtheria**

**Exclusion Criteria: A non-endemic pathogen with no ability to establish local transmission (high immunity/vaccination rates) and minimal impact from imports.**

- *Notes and references: Because of lack of data we make the conservative assumption that imports will be equal to the total number of cases of diphtheria in the US since 2016 and a 20% increase[57]. Susceptibility estimates are based on 4 dose tdap coverage [47]. $R_0$ assumes an increase in transmission at the World Cup.*
- *Expected Excess Importations:* 0.0187
- *Additional Infections due to Transmission (per cases):* 0.76
    - simulation parameters $R_0 = 3.2$, $k = 0.93$, $g = 7.8$, 20% susceptibility

- *Total Additional Infections:* 0.03
- *Cost per case:* $179,201 (conservative based on high DALY impact)
- *Expected Impact:* $5,800

---

**E. coli (STEC)**

**Exclusion criteria: Endemic pathogen with no increased transmission risk at World Cup related activities and increased local prevalence due to importations of less than 5%.**

- *Notes and references: 15% of cases are associated with international travel normally, which also tends to be to non-WC countries*[58]*. A 20% increase would be a three percent increase in overall prevalence.*

---

**Ebola (EVD) [outbreak/hypothetical]**

**Exclusion Criteria: A non-endemic pathogen with no ability to establish local transmission (no documented extended transmission in high income countries) and exceedingly low importation probability.**

- *Notes and references:* Importation estimates assume the DRC/Uganda outbreak is 1,000 infections and ignore the impact of the travel ban. Rates based on analysis of excess travel. Per introduction costs based on introductions in 2014[33].
- *Expected Excess Importations:* 0.00065
- *Additional Infections due to Transmission (per cases):* 1.87
  - simulation parameters $R_0 = 1.8$, $k = 0.35$, $g = 15.4$, 100% susceptibility
- *Total Additional Infections:* 0.00183
- *Cost per case:* $4,300,000
- *Expected Impact:* $7,852

---

**Ehrlichiosis**

**Exclusion criteria: Endemic pathogen with no increased transmission risk at World Cup related activities and increased local prevalence due to importations of less than 5%.**

- *Notes and references: E. chaffeensis causes majority of human cases and is almost exclusively reported in North America.;*[59]*];*[60]

---

**Giardiasis**

**Exclusion criteria: Endemic pathogen with no increased transmission risk at World Cup related activities and increased local prevalence due to importations of less than 5%.**

- *Notes and references: Data on global incidence is sparse. Over 1 million cases per year in the United States*[61]*. The pathogen is environmentally ubiquitous in the US, with comparable prevalence in non-human animals to the rest of the globe*[62]*.*

---

**Glanders (B. mallei)**

**Exclusion Criteria: A non-endemic pathogen with no ability to establish local transmission (humans are a dead end host) and negligible impact from imports.**

- *Notes and references: Eliminated from North America*[63]*. Only less than 20 cases are reported globally per year, liberal importation estimate based on this.*
- *Expected Excess Importations:* 0.000001
- *Additional Infections due to Transmission (per cases):* 0
  - No transmission in humans
- *Total Additional Infections:* 0.000001
- *Cost per case:* $10,000,000 (high IFR)
- *Expected Impact:* $10

---

**HIV**

**Exclusion criteria: Endemic pathogen with no increased transmission risk at World Cup related activities and increased local prevalence due to importations of less than 5%.**

- *Notes and references: See note under "Syphilis" about evidence indicating no increase in STIs associated with international sporting events. To assess the potential for increased WC-related transmission of bloodborne infections, we conducted a rapid literature review with the following search terms: ("injection drug use" OR "injecting drug use" OR "needle sharing" OR PWID OR bloodborne OR parenteral) AND (sport* OR "mass gathering") AND (outbreak OR increase*) Results did not yield any evidence of increased transmission of bloodborne pathogens surrounding sporting events. NATIONAL ESTIMATES Most recent (2022) CDC* *estimate for HIV prevalence in US: 1.2 million Most recent (2022) CDC estimate for annual US HIV incidence = 31,800 Most recent (2022) CDC estimate for annual new HIV diagnoses in US = 37,981 US Census estimate for US population size (2022) = 333.3 million Estimated US HIV prevalence as proportion of US population = 1.2 / 333.3 = 0.0036 WORLD CUP CITY ESTIMATES Population size, World Cup cities (from "Local Presence" sheet): 84.6 million Estimated number of prevalent HIV cases, World Cup cities: 0.0036* 84.6 million = 304,560 prevalent cases Number of imported cases needed to meet 5% local prevalence threshold: 0.05 * 304,560 = 15,228 imported cases Estimated annual # new HIV infections in World Cup Cities = 31,800 * (84.6/333.3) = 8,072 Estimated annual # new HIV diagnoses in World Cup Cities = 37,981 * (84.6/333.3) = 9,641 COMPARISONS: WC IMPORTATIONS>5% LOCAL PREVALENCE vs. "TYPICAL" IMPORTATIONS One approach: Estimated % of new HIV diagnoses in US among people born outside US = 18.9% Estimated annual # of new HIV diagnoses in World Cup Cities among people born outside the US = 0.189*9641 = 1,822 For World Cup importations of HIV to exceed the 5% local prevalence threshold, we would need 15,228/1,822 = 8 times as many people with HIV to enter the US for World Cup games over 6 weeks as the estimated annual number of new HIV diagnoses in World Cup cities among people born outside the US Another approach: Estimated % of US HIV cases linked to potential transmission partner from outside the US via phylogenetic analysis = 3.5% Proxy estimate of "normal/typical" annual importations in US = 0.035*31,800 = 1,113 Proxy estimate of "normal/typical" annual importations in WC Cities = 0.035*8072 = 283 For WC importations of HIV to exceed 5% local prevalence, we would need 15,228/283 = 54 times as many people w/ HIV to enter US for WC games over 6 wks as the estimated # of annual new HIV infections in WC cities from contacts outside US.; References: [64] [65][66] [67]

**HPAI (H5N1)**

**Exclusion criteria: Endemic pathogen with no increased transmission risk at World Cup related activities and increased local prevalence due to importations of less than 5%.**

- *Notes and references:* While cases are sporadic, H5N1 occurrence in the US in recent years has been not infrequent[68]. In contrast, most recent cases elsewhere in the world are in places without World Cup teams, and US has higher cases than elsewhere in recent years. [69].

**Hand Foot & Mouth**

**Exclusion criteria: Endemic pathogen with no increased transmission risk at World Cup related activities and increased local prevalence due to importations of less than 5%.**

- *Notes and reference:* *Has a high prevalence in the United States, particularly in recent years (e.g., Virginia[70]). The vast majority of cases are in children under 10, making World Cup importations unlikely[71].

**Hendra Virus**

**Exclusion Criteria: A non-endemic pathogen with no ability to establish local transmission (zoonotic host absent) and negligible impact from imports.**

- *Notes and references:* There have been no reported human cases of Hendra virus since 2009[72].

**Hepatitis A**

**Exclusion criteria: Endemic pathogen with no increased transmission risk at World Cup related activities and increased local prevalence due to importations of less than 5%.**

- *Notes and references: Based on numbers in the Global Burden of Disease Study estimates for US, we would expect there to be ~112,000 cases in host cities and only ~935 excess imports during the World Cup*

**Hepatitis C**

**Exclusion criteria: Endemic pathogen with no increased transmission risk at World Cup related activities and no reasonable chance of healthcare outcomes during the World Cup.**

- *Notes and references:* Primary sequelae of Hepatitis C are chronic conditions unlikely to impact the United States during visitors time at the World Cup. Likewise, there is no evidence or reason to believe there will be increase injection drug use during the World Cup.

**Hepatitis E**

**Exclusion criteria: Endemic pathogen with no increased transmission risk at World Cup related activities and increased local prevalence due to importations of less than 5%.**

- *Notes and references: Despite being uncommon, Hepatitis E is endemic to the Unites States[73] and:[74] [73]. However, increased transmission at the World Cup is unlikely as in developed settings mostly spread via consumption of undercooked meat[75]. Based on [REFERENCE] there are 4,621 expected cases in World Cup*

| | |
|---|---|
| | *communities compared to an expected 35 excess imports 4621 expected cases in WC communities (see incidence column) compared to 35 expected excess imports.* |
| **Legionella** | **Exclusion criteria: Endemic pathogen with no increased transmission risk at World Cup related activities and increased local prevalence due to importations of less than 5%.**<br>• *Notes and references: There are around 6,000 cases per year in the US. Real incidence maybe in the 25,000 per year.* [76] *Global incidence is projected to be similar to the US (10-15 per million) so a >5% increase in local incidence is highly unlikely.*[77] *(Prevalence/incidence in the WC population would have to be 6.5 that in the US population in order to hit the 5% mark)*[76] [77]. |
| **Leptospirosis** | **Exclusion criteria: Endemic pathogen with no increased transmission risk at World Cup related activities and increased local prevalence due to importations of less than 5%.**<br>• *Notes and references: About 200 cases a year occur in the US with only ~13% imported. A 20% (or even 35%) increase in importations would not result in significant increase in transmission.*[78]*; Global burden concentrated in South Asia and sub-saharan Africa (areas of low increase in travel during the tournament).* |
| **Listeriosis** | **Exclusion criteria: Endemic pathogen with no increased transmission risk at World Cup related activities and increased local prevalence due to importations of less than 5%.**<br>• *Notes and references: 0.24 cases per 100,000 population in US.*[79] *Highest global incidence about 10 per million, which is less than 6.5 times the US (incidence must be 6.5x higher in WC attendees to achieve a 5% increase)* [80] *[60] [59]* |
| **Lyme Disease** | **Exclusion criteria: Endemic pathogen with no increased transmission risk at World Cup related activities and increased local prevalence due to importations of less than 5%.**<br>• *Notes and references:* While present in other parts of the world, Lyme Disease is massively more prevalent in the United States, with incidence rates 6 or more times higher than other countries[81]. |
| **MERS-CoV** | **Exclusion Criteria: A non-endemic pathogen with no ability to establish local transmission (zoonotic host absent) and negligible impact from imports.**<br>• *Notes and references:* Camels are not common in the Unites States, so MERS-CoV could not establish endemic transmission. Importations are rare, based on 2025 incidence in Saudi Arabia there is a less than 0.2% chance of an importation. Even assuming a high per case response cost, impact would be minimum.<br>• *Expected Excess Importations:* 0.00148<br>• *Additional Infections due to Transmission (per cases):* 2.29<br> ◦ simulation parameters (highly pessimistic) $R_0 = 2.3$, $k = 0.09$, $g : 14.1$, susceptibility = 100%.<br>• *Total Additional Infections:* 0.00487<br>• *Cost per case:* $1,000,000 (pessimistic default)<br>• *Expected Impact:* $4,870 |
| **Marburg (MVD) [Hypothetical Scenario]** | **Exclusion Criteria: A non-endemic pathogen with no ability to establish local transmission (no documented extended transmission in high income countries) and exceedingly low importation probability.**<br>• *Notes and references:* Importation estimates assume a ***hypothetical*** outbreak in Ghana with 50 infections. Rates based on analysis of excess travel. Per introduction costs based on introductions of Ebola in 2014[33].<br>• *Expected Excess Importations:* 0.00182<br>• *Additional Infections due to Transmission (per cases):* 2.19<br> ◦ simulation parameters $R_0 = 1.6$, $k = 0.35$, $g = 9$, 100% susceptibility<br>• *Total Additional Infections:* 0.0058<br>• *Cost per case:* $4,300,000<br>• *Expected Impact:* $24,972 |

**Melioidosis (B. pseudomallei)**

**Exclusion criteria: Endemic pathogen with no increased transmission risk at World Cup related activities and increased local prevalence due to importations of less than 5%.**

- *Notes and references: The disease is present, but rare, in the United States, however, none of the most sources of the disease are in the World Cup.[82] While human to human transmission is possible, it is extremely rare, and contact with environmental sources is more common. In terms of importation risk, melioidosis most commonly occurs in Northern Australia and Southeast Asia[83]. Of countries in these areas Australia is the only country participating in the World Cup. In Australia, the main population centers are not in endemic regions. Additionally, melioidosis in Australia is seasonal, with the peak occurring in January - April[84]. The causative agent of melioidosis is present in the Americas (including in the US), however most human cases in the Americas occur in Brazil where incidence is concentrated in a single state in the northeast which is only about 4% of the Brazilian population[85].*

**Meningococcal**

**Exclusion criteria: Endemic pathogen with no increased transmission risk at World Cup related activities and increased local prevalence due to importations of less than 5%.**

- *Notes and references: Meningitis has been associated with sporting events, but in the context of congregate living settings, and reviews have shown no evidence of elevated risk [CITE przegl.Epidem 3-2009.pdf]. Prevalence/incidence in the WC population would have to be 6.5 that in the US population in order to hit the 5% mark. Carriage in the US is 5-10% according to the CDC. [86] This means we would have to have 32.5-65% carriage among WC attendees to increase incidence by 5%...which is higher than even high risk setting rates, except some studies of University students:[87] [86]*

**Methicillin-resistant Staphylococcus aureus (MRSA)**

**Exclusion criteria: Endemic pathogen with no increased transmission risk at World Cup related activities and increased local prevalence due to importations of less than 5%.**

- *Notes and references: Prevalence of MRSA is high in the US compared to elsewhere in the world. One of the highest rates based on here:[88] And here:[89]*

**Mpox Clade IIb**

**Exclusion criteria: Endemic pathogen with no increased transmission risk at World Cup related activities and increased local prevalence due to importations of less than 5%.**

- *Notes and references:* Incidence of clade IIb mpox is not significantly higher anywhere in the world than the US beside certain parts of Africa for which travel to the United States is exceedingly low[90] [Srivastava2025MpoxClades].*

**Mpox Clade Ib**

**Exclusion Criteria: A non-endemic pathogen with no ability to establish local transmission (no documented extended transmission in high income countries, reproductive number below one) and exceedingly low importation probability.**

- *Notes and references:* Importations based on analysis of travel data and distribution of Mpox Clade Ib case (see importation methods and results). Total cost per case (USD):* Using the Clade II costs from [91]
- *Expected Excess Importations:* 0.0253
- *Additional Infections due to Transmission (per cases):* 1.1
  - simulation parameters $R_0 = 0.9$, $k = 0.26$, $g = 8.59$, 100% susceptibility
- *Total Additional Infections:* 0.0543
- *Cost per case:* $3,840
- *Expected Impact:* $208

**Mumps**

**Exclusion criteria: Endemic pathogen with increased transmission risk at World Cup related activities but not enough anticipated impact to warrant inclusion.**

- *Notes and references:* Local prevalence calculated based on US incidence of ~400 mumps cases per year over the past 3 years[92]. Importations based on travel analysis. Pessimistic susceptibility estimates are used, applying the top susceptibility in any age group from[93] to the overall population. Cost is based of the reported cost of public health response per case.[94]
- *Expected Excess Importations:* 0.52
- *Local Presence at World Cup Related Activities:* 0.4

- *Additional Infections due to Transmission (per imported case):* 3.3 (World Cup), 2.8 (normal)
    - simulation parameters $R_0 = 5.4$ (community), $R_0 = 6.5$ (World Cup), $k = 0.93$, $g = 18.12$, 52.6% susceptibility
- *Total Additional Infections:* 2.5
- *Cost per Case:* $3,950
- *Expected Impact:* $9,827

---

**Nipah Virus**

**Exclusion Criteria: A non-endemic pathogen with no ability to establish local transmission (zoonotic host absent) and zero impact from imports.**

- *Notes and references:* The bat species which is Niaph virus's zoonotic host does not exist in the US. There are no cases in any of the World Cup countries, so expected excess importations is 0.[95]
- *Expected Excess Importations:* 0
- *Expected Impact:* $0

---

**Oropouche**

**Exclusion Criteria: A non-endemic pathogen with no ability to establish local transmission (lack of competent vector).**

- *Notes and references:* Though 108 cases of Oropouche virus were detected in the US from 2024-2025, the risk of efficient transmission of Oropouche in the US is considered low due to the limited abundance and human interaction of the most effective vector, Culicodes Paraensis.[96,97] Other possible vectors present in the US. Other vectors in the US have been evaluated and have been found to have low competence.[98]
- *Expected Excess Importations:* 2.31
- *Additional Infections due to Transmission (per cases):* 1.73
    - simulation parameters $R_0 = 0.995$, $k = 0.02$, $g = 5.5$, 100% susceptibility
- *Total Additional Infections:* 2.3
- *Cost per case:* $2,700 (based on costs for chikungunya)[99]
- *Expected Impact:* $6,231

---

**Paratyphoid Fever**

**Exclusion Criteria: A non-endemic pathogen with no ability to establish local transmission (water and sanitation conditions) and minimal impact from imports.**

- *Notes and references: Paratyphoid is absent from countries with adequate water and sanitation infrastructure. Importations calculated based on CDC data and number of importations each year, assuming a 20% increase concentrate in WC locations.* [100,101]. Lacking any data on industrialized country $R_0$, numbers for Typhoid were used.
- *Expected Excess Importations:* 1.92
- *Additional Infections due to Transmission (per cases):* 0
- *Total Additional Infections:* 5.3
- *Cost per case:* $1,490 (assumed to be same as typhoid)
- *Expected Impact:* $7,830

---

**Pertussis**

**Exclusion criteria: Endemic pathogen with no increased transmission risk at World Cup related activities and increased local prevalence due to importations of less than 5%.**

- *Notes and references:* Travel importation analysis suggests 0.65 excess importations per year, which is <5% of the expected cases in World Cup metro areas given an US incidence rate of ~0.55 per 100,000[102,103,104].

---

**Plague (Y. pestis)**

**Exclusion criteria: Endemic pathogen with no increased transmission risk at World Cup related activities and increased local prevalence due to importations of less than 5%.**

- *Notes and references:* There are ~2000 case of plague globally each year, and ~7 annually in the US[105,106]. Increase travel to the US during the World Cup would increase expected prevalence to 5% even under very pessimistic assumptions about these cases.

**Pneumonia (Bacterial)**

**Exclusion criteria: Endemic pathogen with no increased transmission risk at World Cup related activities and increased local prevalence due to importations of less than 5%.**

- *Notes and references: Using numbers from* [107] *CAP rates are 28.4 per 10,000 in the US, leading to approximately 22,500 expected cases in WC metro areas during the tournament. Even if we assumed all importees had the highest known rates of 14 per 1000, we get 14/1000* 650000 * 39/365 =972 during the tournament, which is less than 5% of 22,500.[107]*

**Polio**

**Exclusion Criteria: A non-endemic pathogen with no ability to establish local transmission (high vaccination rates) and zero impact from imports.**

- *Notes and references:* No wild type polio cases have occurred in World Cup countries in the past year, so excess importation risk is zero[108].
- *Expected Excess Importations:* 0
- *Expected Impact:* $0

**Psittacosis**

**Exclusion criteria: Endemic pathogen with no increased transmission risk at World Cup related activities and increased local prevalence due to importations of less than 5%.**

- *Notes and references:* Among the outbreaks found in this meta-review, 35-45% of cases occurred in the US despite US being only 4-5% of global population.[109]

**Q Fever**

**Exclusion criteria: Endemic pathogen with no increased transmission risk at World Cup related activities and increased local prevalence due to importations of less than 5%.**

- *Notes and references: US Incidence ~200 per year*[110] *Highest incidence reported region is EU with 900-1100 cases annually.*[111] *Disease length ~2 weeks*[112] *Population of the EU: 450,000,000 Based on this you would need roughly (200*.05)/(1100/450000000) = 4090909 travelers from the EU for the World Cup to increase local prevalence by 5%[110][111112]*

**Rabies**

**Exclusion criteria: Endemic pathogen with no increased transmission risk at World Cup related activities and increased local prevalence due to importations of less than 5%.**

- *Notes and references: From 1990 to 2012 there were 16 imported rabies cases in the US. This corresponds to an expected monthly excess importation of 0.0116.*[113]

**Ricin (Toxin)**

**Exclusion criteria: Bioterrorism agent with no probability of natural importation or incidence.**

- *Notes and references: Included based on original list criteria, but pure bioterrorism agents are outside of scope of screening.*

**Rubella**

**Exclusion Criteria: A non-endemic pathogen with no ability to establish local transmission (high vaccination rates) and minimal impact from imports.**

- *Notes and references:* Note that $R_0$ used in analysis is based on elevated world cup transmission. Immunity assumed to be same as measles assumptions. Importations based on analysis of travel data and incidence. Note that $R_0$ is adjusted for World Cup increases.
- *Expected Excess Importations:* 0.22
- *Additional Infections due to Transmission (per cases):* 0.74
  - simulation parameters $R_0 = 6.24$, $k = 0.93$, $g = 18.3$, S: 11.9% susceptiblity
- *Total Additional Infections:* 0.39
- *Cost per case:* $9,339
- *Expected Impact:* $3,600

**Salmonella**

**Exclusion criteria: Endemic pathogen with no increased transmission risk at World Cup related activities and increased local prevalence due to importations of less than 5%.**

- *Notes and references: Given high US incidence a 5% increase due to importations does not seem plausible.; Based on GBD estimate* [114] *and country SDIs, expected incidence in the US is 0.89 versus 0.99 among*

| | |
|---|---|
| | *travelers, which does not reach the 6.5 threshold for inclusion. [96]* |
| **Shigellosis** | **Exclusion criteria: Endemic pathogen with no increased transmission risk at World Cup related activities and increased local prevalence due to importations of less than 5%.**<br>• *Notes and references: Already highly prevalent in the US*[115] |
| **Smallpox** | **Exclusion criteria: Bioterrorism agent with no probability of natural importation or incidence.**<br>• *Notes and references: Included based on original list criteria, but pure bioterrorism agents are outside of scope of screening.* |
| **Spotted Fever (RMSF)** | **Exclusion criteria: Endemic pathogen with no increased transmission risk at World Cup related activities and increased local prevalence due to importations of less than 5%.**<br>• *Notes and references:* Only circulates in the Americas, particularly the United States[116]. |
| **St. Louis Encephalitis** | **Exclusion criteria: Endemic pathogen with no increased transmission risk at World Cup related activities and increased local prevalence due to importations of less than 5%.**<br>• *Notes and references:* Human cases occur pretty much exclusively in the US[117]. |
| **Syphilis** | **Exclusion criteria: Endemic pathogen with no increased transmission risk at World Cup related activities and increased local prevalence due to importations of less than 5%.**<br>• *Notes and references: Calculations focus on "early syphilis," which corresponds approximately to the first year of infection when infectiousness is highest (and is thus the relevant stage of infection for which we must estimate the potential for importations to increase local prevalence). With the duration of early syphilis being approximately one year, estimates of annual incidence approximate prevalence of early syphilis (assuming approximately steady-state conditions). With numerous additional assumptions, simplifications, and caveats, we have: NATIONAL ESTIMATES Most recent estimate for syphilis incidence in US (2018): 146,000 incident infections among persons aged 14-49 CDC estimate for ratio of reported early syphilis cases in US, 2024:2018 = 89,035/73,062 = 1.22 Estimated US prevalence of early syphilis (2024) = 1.22*146,000 = 178,120 cases among persons aged 14-49 US Census estimate for US population size (2024) = 340.1 million % US population who are adults (2024) = 88% Size of adult population in US (2024) = 0.88*340.1 million = 299.3 million Estimated early syphilis prevalence as proportion of US population (2024) = 178,120 / 299,300,000 = 0.0006 WORLD CUP CITY ESTIMATES Population size, World Cup cities (from "Local Presence" sheet): 84.6 million Estimated number of prevalent early syphilis cases, WC cities: 0.0006*84.6 million = 50,760 early cases Number of imported early syphilis cases needed to meet 5% local prevalence threshold: = 0.05*50,760 = 2,538 Estimated travelers coming to US for World Cup = 1 million WHO estimate of global annual syphilis incidence in adults, 2022 = 8 million World adult population size, 2022 = 6 billion Estimated world prevalence of early syphilis = 8 million / 6 billion = 0.001 Total importations of early syphilis = 0.001*1 million = 1,000 (<5% of estimated WC city prevalence of early syphilis); References:[118] [119][120]* |
| **Trichinellosis** | **Exclusion criteria: Endemic pathogen with no increased transmission risk at World Cup related activities and increased local prevalence due to importations of less than 5%.**<br>• *Notes and references: US incidence is about 15 per year and there are an estimated 10,000 cases per year internationally. Importation expectation if these were randomly distributed in WC travelers importations would be about 1 in 10,000,000 (10000/8000000000*39/365 = 10^-7) which is below the 5% increase risk.*[121]* |
| **Tularemia** | **Exclusion criteria: Endemic pathogen with no increased transmission risk at World Cup related activities and increased local prevalence due to importations of less than 5%.**<br>• *Notes and references: [122][123] [116] [117]; About 200 cases per year in the US. Very rare overall, and prevalent in the United States. Most common in Scandinavian countries, but not in rates likely to increase risk in US [124] [118]* |
| **Typhoid Fever** | **Exclusion Criteria: A non-endemic pathogen with no ability to establish local transmission (water and sanitation conditions) and minimal impact from imports.** |

| | |
|---|---|
| | • *Notes and references:* Importation risk based on travel analysis. Reproductive number based on estimates from Taiwan (industrialized setting). Cost based on[30] adjusted for inflation.[30]<br>• *Expected Excess Importations:* 0.0278<br>• *Additional Infections due to Transmission (per cases):* 0.96<br> ◦ simulation parameters $R_0 = 0.995$, $k = 0.003$, $g = 14.5$, 100% susceptibility<br>• *Total Additional Infections:* 0.055<br>• *Cost per case:* $1,490<br>• *Expected Impact:* $81 |
| **VRSA** | **Exclusion criteria: Endemic pathogen with no increased transmission risk at World Cup related activities and increased local prevalence due to importations of less than 5%.**<br>• *Notes and references: Present at very low levels globally and transmission is most common in healthcare settings. Present in US at similar prevalence.*[45]. |
| **Vibriosis** | **Exclusion criteria: Endemic pathogen with no increased transmission risk at World Cup related activities and increased local prevalence due to importations of less than 5%.**<br>• *Notes and references: Globally ubiquitous, even in the US, and there are lot's of cases in the US.*[125] |
| **West Nile Virus** | **Exclusion criteria: Endemic pathogen with no increased transmission risk at World Cup related activities and increased local prevalence due to importations of less than 5%.**<br>• *Notes and references: Incidence is higher in North America than elsewhere in the world. Hence importations are unlikely to increase local incidence by more than 5%.*[126]. |
| **Yellow Fever** | **Exclusion Criteria: A non-endemic pathogen with no ability to establish local transmission (vector conditions and no support of a sylvatic cycle) and minimal impact from imports.**<br>• *Notes and references:* Despite percent of *Aedes* mosquitos there has been no documented human transmission of yellow fever in the United States since 1905, despite introductions.[127] [128] |

#### References


1. S Hirve, LP Newman, J Paget, E Azziz-Baumgartner, J Fitzner, N Bhat, others. Influenza seasonality in the tropics and subtropics - when to vaccinate? *PLoS One*. 2016;11(4):e0153003. doi:10.1371/journal.pone.0153003
2. Preliminary estimates of COVID. Accessed May 24, 2026. https://www.cdc.gov/covid/php/surveillance/burden-estimates.html
3. Infection rates and symptomatic proportion of SARS. Accessed May 24, 2026. https://wwwnc.cdc.gov/eid/article/30/9/24-0065_article
4. COVID. Accessed May 24, 2026. https://www.cdc.gov/yellow-book/hcp/travel-associated-infections-diseases/covid-19.html
5. Current epidemic trends (based on rt) for states. Accessed May 26, 2026. https://www.cdc.gov/cfa-modeling-and-forecasting/rt-estimates/index.html?tab=1
6. Global influenza programme. Accessed May 26, 2026. https://www.who.int/tools/flunet
7. CB Hall, JM Geiman, R Biggar, DI Kotok, PM Hogan, Douglas GR Jr. Respiratory syncytial virus infections within families. *N Engl J Med*. 1976;294(8):414-419. doi:10.1056/NEJM197602192940803
8. NH Leung, C Xu, DK Ip, BJ Cowling. Review article: The fraction of influenza virus infections that are asymptomatic: A systematic review and meta-analysis. *Epidemiology*. 2015;26(6):862-872. doi:10.1097/EDE.0000000000000340
9. About estimated flu burden. Accessed May 24, 2026. https://www.cdc.gov/flu-burden/php/about/index.html
10. Katherine Kendrick, Danielle Stanek, Carina Blackmore. Transmission of chikungunya virus in the continental united states — florida, 2014. *Morbidity and Mortality Weekly Report*. 2014;63(48):1137.
11. Lizis O. Rodriguez, Eli B. Levitt, Nahal Khamisani, Sarah Nickle, Guillermo Izquierdo-Pretel. Local transmission of dengue in south florida: A case report. *Cureus*. 2024;16(7):e65375. doi:10.7759/cureus.65375

12. Lin H. Chen, Carlos Marti, Clemente Diaz Perez, Bianca M. Jackson, Alyssa M. Simon, Mei Lu. Epidemiology and burden of dengue fever in the united states: A systematic review. *Journal of Travel Medicine*. 2023;30(7):taad127. doi:10.1093/jtm/taad127
13. Coreen M. Beaumier, Melissa N. Garcia, Kristy O. Murray. The history of dengue in the united states and its recent emergence. *Current Tropical Medicine Reports*. 2014;1(1):32-35. doi:10.1007/s40475-013-0008-1
14. Nikhil Ranadive, Joel L. N. Barratt, Ellen M. Dotson, John E. Gimnig, Audrey E. Lenhart, Rebecca S. Levine, Kimberly E. Mace, Peter D. McElroy, Brian H. Raphael, Dean Sayre, Laura C. Steinhardt, Alica C. Sutcluffe, Seymour G. Willimas, Alison D. Ridpath. CDC operational guidance for investigating locally acquired mosquito-transmitted malaria — united states, 2026. *Morbidity and Mortality Weekly Report*. 2026;75(1):1-14. https://www.cdc.gov/mmwr/volumes/75/rr/rr7501a1.htm
15. Zika cases in the united states. Accessed May 26, 2026. https://www.cdc.gov/zika/zika-cases-us/index.html
16. Hepatitis b basics. Accessed May 24, 2026. https://www.cdc.gov/hepatitis-b/about/index.html
17. Global hepatitis report 2026. Accessed May 24, 2026. https://www.who.int/teams/global-hiv-hepatitis-and-stis-programmes/hepatitis/reports/global-hepatitis-report-2026
18. H Roberts, KN Ly, S Yin, E Hughes, E Teshale, R Jiles. Prevalence of HBV infection, vaccine-induced immunity, and susceptibility among at-risk populations: US households, 2013-2018. *Hepatology*. 2021;74(5):2353-2365. doi:10.1002/hep.31991
19. Mindie H. Nguyen, A. Burak Ozbay, Iris Liou, Nicole Meyer, Stuart C. Gordon, Geoffrey Dusheiko, Joseph K. Lim. Healthcare resource utilization and costs by disease severity in an insured national sample of US patients with chronic hepatitis B. *Journal of Hepatology*. 2019;70(1):24-32. doi:10.1016/j.jhep.2018.09.021
20. Sandra E. Talbird, Seri A. Anderson, Misha Nossov, Nell Beattie, Aaron T. Rak, Francisco Diaz-Mitoma. Cost-effectiveness of a 3-antigen versus single-antigen vaccine for the prevention of hepatitis B in adults in the United States. *Vaccine*. 2023;41(23):3506-3517. doi:10.1016/j.vaccine.2023.04.022
21. Noele P. Nelson, Philippa J. Easterbrook, Brian J. McMahon. Epidemiology of Hepatitis B Virus Infection and Impact of Vaccination on Disease. *Clinics in liver disease*. 2016;20(4):607-628. doi:10.1016/j.cld.2016.06.006
22. KG Castro, SM Marks, MP Chen, AN Hill, JE Becerra, R Miramontes, others. Estimating tuberculosis cases and their economic costs averted in the united states over the past two decades. *Int J Tuberc Lung Dis*. 2016;20(7):926-933. doi:10.5588/ijtld.15.1001
23. 404. Accessed May 24, 2026. https://wwwnc.cdc.gov/eid/article/31/3/24-0633_article.
24. Eurosurveillance. Accessed May 24, 2026. https://www.eurosurveillance.org/content/10.2807/1560-7917.ES2014.19.35.20891
25. KR Ehresmann, CW Hedberg, MB Grimm, CA Norton, KL MacDonald, MT Osterholm. An outbreak of measles at an international sporting event with airborne transmission in a domed stadium. *J Infect Dis*. 1995;171(3):679-683. doi:10.1093/infdis/171.3.679
26. Quantifying the cost of measles outbreak in the u.s. And how costs scale with outbreak size. Accessed May 24, 2026. https://www.medrxiv.org/content/10.1101/2025.10.24.25338724v2.full-text#T1
27. MK Steele, ME Wikswo, AJ Hall, K Koelle, A Handel, K Levy, others. Characterizing norovirus transmission from outbreak data, united states. *Emerg Infect Dis*. 2020;26(8):1818-1825. doi:10.3201/eid2608.191537
28. Norovirus outbreaks: A systematic review of commonly implicated transmission routes and vehicles. Accessed May 24, 2026. https://www.cambridge.org/core/journals/epidemiology-and-infection/article/norovirus-outbreaks-a-systematic-review-of-commonly-implicated-transmission-routes-and-vehicles/13C8B35A600A54819F0F4EFF84A3FB9B
29. NoroSTAT data. Accessed May 24, 2026. https://www.cdc.gov/norovirus/php/reporting/norostat-data.html
30. Economic burden of foodborne illnesses acquired in the united states. Accessed May 24, 2026. https://journals.sagepub.com/doi/10.1089/fpd.2023.0157?url_ver=Z39.88-2003&rfr_id=ori:rid:crossref.org&rfr_dat=cr_pub%20%200pubmed
31. As norovirus outbreaks grow, health departments respond: Millions infected each year with virus. Accessed May 24, 2026. https://www.thenationshealth.org/content/43/2/1.3
32. Diana L. Von Stein, Alexandra Barger, Andrew Hennenfent, Robert Ramaekers, Amanda Mandi, Kenzie Teno, Karen Brust, Jonathan Simmons, Nicholas Mohr, Lisa Veach, Sudhir Kumar, Aneesa Afroze, Emily McCutchen, Amanda Bartling,

Michael Pentella, Megan Nelson, Jennifer Craft, Rikki Hetzler, Amy Thoreson, Alicia Coppedge, Sam Jarvis, Jennifer Miller, Alison M. Todres, Jessica L. Wickline, Sheena Tarrant, Leanna Sayyad, Inna Krapiunaya, Amy Schuh, Amy Whitesell, Gerard C. Kuotu, Kiara McNamara, Nancy Cornish, Shelly Schwedhelm, Angela Vasa, Angela Hewlett, Shantyl Galloway, Aaron D. Kofman, Katrin S. Sadigh, Robert Kruse, Barbara Knust, Matthew Donahue. Notes from the field: Response to a case of travel-associated lassa fever — iowa, october–november 2024. *Morbidity and Mortality Weekly Report*. 2025;74(11):194-196. doi:10.15585/mmwr.mm7411a3
33. Nicholas G. Reich, Justin Lessler, Jay K. Varma, Neil M. Vora. Quantifying the risk and cost of active monitoring for infectious diseases. *Scientific Reports*. 2018;8(1):1093. doi:10.1038/s41598-018-19406-x
34. J Lessler, T Dos Santos, X Aguilera, R Brookmeyer, PAHO Influenza Technical Working Group, DAT Cummings. H1N1pdm in the americas. *Epidemics*. 2010;2(3):132-138. doi:10.1016/j.epidem.2010.07.001
35. About c. Perfringens food poisoning. Accessed May 24, 2026. https://www.cdc.gov/clostridium-perfringens/about/index.html
36. Epidemiology and statistics. Accessed May 24, 2026. https://www.cdc.gov/anaplasmosis/hcp/statistics/index.html
37. Journal of vector borne diseases. Accessed May 26, 2026. https://journals.lww.com/jvbd/fulltext/2023/60030/global_status_of_anaplasma_phagocytophilum.6.aspx?context=latestarticles
38. Anthrax infection. Accessed May 24, 2026. https://www.ncbi.nlm.nih.gov/books/NBK535379/
39. Data and statistics on babesiosis. Accessed May 24, 2026. https://www.cdc.gov/babesiosis/php/data-stats/index.html
40. EZproxy. Accessed May 26, 2026. https://pmc-ncbi-nlm-nih-gov.libproxy.lib.unc.edu/articles/PMC9132453/#s0003
41. Www.sciencedirect.com. Accessed May 24, 2026. https://www.sciencedirect.com/science/article/abs/pii/S0963996924017216
42. Clinical overview of brucellosis. Accessed May 24, 2026. https://www.cdc.gov/brucellosis/hcp/clinical-overview/index.html
43. CG Laine, VE Johnson, HM Scott, AM Arenas-Gamboa. Global estimate of human brucellosis incidence. *Emerg Infect Dis*. 2023;29(9):1789-1797. doi:10.3201/eid2909.230052
44. Www.cdph.ca.gov. Accessed May 24, 2026. https://www.cdph.ca.gov/Programs/CID/DCDC/CDPH%20Document%20Library/BrucellosisFactSheet.pdf
45. Y Li, X Sun, N Dong, Z Wang, R Li. Global distribution and genomic characteristics of carbapenemase-producing escherichia coli among humans, 2005-2023. *Drug Resist Updat*. 2024;72:101031. doi:10.1016/j.drup.2023.101031
46. Candida auris. Accessed May 26, 2026. https://www.ncbi.nlm.nih.gov/books/NBK563297/
47. FastStats. Accessed May 24, 2026. https://www.cdc.gov/nchs/fastats/immunize.htm
48. HA Shah, A Meiwald, C Perera, G Casabona, P Richmond, N Jamet. Global prevalence of varicella-associated complications: A systematic review and meta-analysis. *Infect Dis Ther*. 2024;13(1):79-103. doi:10.1007/s40121-023-00899-7
49. Global measles outbreaks. Accessed May 26, 2026. https://www.cdc.gov/global-measles-vaccination/data-research/global-measles-outbreaks/index.html
50. Cost estimates of foodborne illnesses. Accessed May 24, 2026. https://www.ers.usda.gov/data-products/cost-estimates-of-foodborne-illnesses
51. Sandra Hoffmann, Alice E. White, Robert B. McQueen, Jae-Wan Ahn, Lauren B. Gunn-Sandell, Elaine J. Scallan Walter. Economic Burden of Foodborne Illnesses Acquired in the United States. *Foodborne Pathogens and Disease*. 2025;22(1):4-14. doi:10.1089/fpd.2023.0157
52. O Ergönül. Crimean-congo haemorrhagic fever. *Lancet Infect Dis*. 2006;6(4):203-214. doi:10.1016/S1473-3099(06)70435-2
53. Factsheet for health professionals about crimean. Accessed May 26, 2026. https://www.ecdc.europa.eu/en/crimean-congo-haemorrhagic-fever/facts/factsheet
54. Ilkay Bozkurt, Mustafa Sunbul, Hava Yilmaz, Saban Esen, Hakan Leblebicioglu, Nicholas J. Beeching. Direct healthcare costs for patients hospitalized with Crimean-Congo haemorrhagic fever can be predicted by a clinical illness severity scoring system. *Pathogens and Global Health*. 2016;110(1):9-13. doi:10.1080/20477724.2015.1136130
55. Prevalence of cryptosporidium infection in the global population: A systematic review and meta. Accessed May 26, 2026. https://link.springer.com/article/10.2478/s11686-020-00230-1

56. AO Markon, A Karasick, C Punzalan, AJ da Silva, B Wolpert. Evaluating foodborne cyclosporiasis using foodborne diseases active surveillance network and foodborne disease outbreak surveillance system data, 2015-2019. *Am J Trop Med Hyg*. 2025;112(2):319-326. doi:10.4269/ajtmh.24-0208
57. Diphtheria surveillance and trends. Accessed May 26, 2026. https://www.cdc.gov/diphtheria/php/surveillance/index.html
58. Risk factors for non. Accessed May 24, 2026. https://wwwnc.cdc.gov/eid/article/29/6/22-1521_article
59. Human monocytotropic ehrlichiosis—a systematic review and analysis of the literature. Accessed May 26, 2026. https://journals.plos.org/plosntds/article?id=10.1371/journal.pntd.0012377#sec007
60. RM Harris, BA Couturier, SC Sample, KS Coulter, KK Casey, R Schlaberg. Expanded geographic distribution and clinical characteristics of ehrlichia ewingii infections, united states. *Emerg Infect Dis*. 2016;22(5):862-865. doi:10.3201/eid2205.152009
61. Giardiasis NNDSS summary report for 2021. Accessed May 24, 2026. https://www.cdc.gov/healthy-water-data/documentation/giardiasis-nndss-summary-report-for-2021.html
62. Kareem Hatam-Nahavandi, Ehsan Ahmadpour, Milad Badri, Aida Vafae Eslahi, Davood Anvari, David Carmena, Lihua Xiao. Global prevalence of giardia infection in nonhuman mammalian hosts: A systematic review and meta-analysis of five million animals. *PLOS Neglected Tropical Diseases*. 2025;19(4):e0013021. doi:10.1371/journal.pntd.0013021
63. Glanders. Accessed May 24, 2026. https://www.woah.org/en/disease/glanders/
64. Estimated HIV incidence and prevalence. Accessed May 24, 2026. https://www.cdc.gov/hiv-data/nhss/estimated-hiv-incidence-and-prevalence.html
65. Fast facts: HIV in the united states. Accessed May 24, 2026. https://www.cdc.gov/hiv/data-research/facts-stats/index.html
66. RP Kerani, A Satcher Johnson, SE Buskin, D Rao, MR Golden, X Hu, others. The epidemiology of HIV among people born outside the united states, 2010-2017. *Public Health Rep*. 2020;135(5):611-620. doi:10.1177/0033354920942623
67. JO Wertheim, AM Oster, AL Hernandez, N Saduvala, MC Bañez Ocfemia, HI Hall. The international dimension of the u.s. HIV transmission network and onward transmission of HIV recently imported into the united states. *AIDS Res Hum Retroviruses*. 2016;32(10-11):1046-1053. doi:10.1089/AID.2015.0272
68. A(H5) bird flu surveillance and human monitoring. Accessed May 24, 2026. https://www.cdc.gov/bird-flu/h5-monitoring/index.html
69. Global human cases with avian influenza a(H5N1), 1997. Accessed May 24, 2026. https://www.cdc.gov/bird-flu/php/surveillance/chart-epi-curve-ah5n1.html
70. Higher rates of hand, foot, and mouth disease in 2025. Accessed May 24, 2026. https://www.vdh.virginia.gov/blog/2025/10/22/higher-rates-of-hand-foot-and-mouth-disease-in-2025/
71. Hand, foot and mouth disease: Current knowledge on clinical manifestations, epidemiology, aetiology and prevention. Accessed May 24, 2026. https://link.springer.com/article/10.1007/s10096-018-3206-x
72. Summary of human cases of hendra virus infection. Accessed May 24, 2026. https://www.health.nsw.gov.au/Infectious/controlguideline/Pages/hendra-case-summary.aspx
73. MB Pisano, C Campbell, C Anugwom, VE Ré, JD Debes. Hepatitis e virus infection in the united states: Seroprevalence, risk factors and the influence of immunological assays. *PLoS One*. 2022;17(8):e0272809. doi:10.1371/journal.pone.0272809
74. MH Kuniholm, RH Purcell, GM McQuillan, RE Engle, A Wasley, KE Nelson. Epidemiology of hepatitis e virus in the united states: Results from the third national health and nutrition examination survey, 1988-1994. *J Infect Dis*. 2009;200(1):48-56. doi:10.1086/599319
75. Hepatitis e basics. Accessed May 24, 2026. https://www.cdc.gov/hepatitis-e/about/index.html
76. Www.osha.gov. Accessed May 24, 2026. https://www.osha.gov/legionnaires-disease
77. Legionellosis. Accessed May 24, 2026. https://www.who.int/news-room/fact-sheets/detail/legionellosis
78. C Atherstone, R Galloway, I Schafer, A Artus, MM Rodriguez, K Ryff, others. Epidemiological, temporal, and geographic trends of leptospirosis in the united states, 2014-2020. *PLoS Negl Trop Dis*. 2025;19(8):e0013427. doi:10.1371/journal.pntd.0013427
79. Clinical overview of listeriosis. Accessed May 26, 2026. https://www.cdc.gov/listeria/hcp/clinical-overview/index.html
80. Listeriosis. Accessed May 24, 2026. https://www.who.int/news-room/fact-sheets/detail/listeriosis
81. BL Stone, Y Tourand, CA Brissette. Brave new worlds: The expanding universe of lyme disease. *Vector Borne Zoonotic Dis*. 2017;17(9):619-629. doi:10.1089/vbz.2017.2127

82. Melioidosis and cases around the world. Accessed May 26, 2026. https://www.cdc.gov/melioidosis/risk-factors/index.html
83. I. Gassiep, M. Armstrong, R. Norton. Human melioidosis. *Clinical Microbiology Reviews*. 2020;33(2):e00006-19. doi:10.1128/CMR.00006-19
84. James D. Stewart, Simon Smith, Enzo Binotto, William J. McBride, Bart J. Currie, Josh Hanson. The epidemiology and clinical features of melioidosis in far north queensland: Implications for patient management. *PLOS Neglected Tropical Diseases*. 2017;11(3):e0005411. doi:10.1371/journal.pntd.0005411
85. Tina J. Benoit, David D. Blaney, Thomas J. Doker, Jay E. Gee, Mindy G. Elrod, Dionne B. Rolim, Timothy J. J. Inglis, Alex R. Hoffmaster, William A. Bower, Henry T. Walke. A review of melioidosis cases in the americas. *The American Journal of Tropical Medicine and Hygiene*. 2015;93(6):1134-1139. doi:10.4269/ajtmh.15-0405
86. Chapter 8: Meningococcal disease. Accessed May 24, 2026. https://www.cdc.gov/surv-manual/php/table-of-contents/chapter-8-meningococcal-disease.html
87. Www.sciencedirect.com. Accessed May 24, 2026. https://www.sciencedirect.com/science/article/pii/S1201971218344321
88. AH Hasanpour, M Sepidarkish, A Mollalo, A Ardekani, M Almukhtar, A Mechaal, others. The global prevalence of methicillin-resistant staphylococcus aureus colonization in residents of elderly care centers: A systematic review and meta-analysis. *Antimicrob Resist Infect Control*. 2023;12(1):4. doi:10.1186/s13756-023-01210-6
89. MRSA infection rates by country. Accessed May 26, 2026. https://onehealthtrust.org/tools/methicillin_resistant_staphylococcus_aureus_infection_rates_united_states_and_other_countries/
90. Toufik Abdul-Rahman, Jann Ludwig Mueller-Gomez, Hala Ibrahim Thaalibi, Innocent Ayesiga, Oyinbolaji Akinwande Ajetunmobi, Aderinto Nicholas, Godfred Yawson Scott, Lukman Lawal, Muritala Adewale Lawal, Selimat Ibrahim. The emergence of clade IIb and ib mpox viruses: A state-of-the-art review. *Annals of Medicine and Surgery*. 2025;88(2):1495-1510. https://journals.lww.com/annals-of-medicine-and-surgery/fulltext/2026/02000/the_emergence_of_clade_iib_and_ib_mpox_viruses__a.48.aspx
91. Www.valueinhealthjournal.com. Accessed May 24, 2026. https://www.valueinhealthjournal.com/article/S1098-3015(23)04141-4/pdf
92. Mumps cases and outbreaks. Accessed May 24, 2026. https://www.cdc.gov/mumps/outbreaks/index.html
93. Www.science.org. Accessed May 24, 2026. https://www.science.org/doi/full/10.1126/scitranslmed.aao5945
94. J Pike, S Schwartz, M Kay, A Perez-Osorio, M Marin, M Jenkins, others. Cost of responding to the 2017 university of washington mumps outbreak: A prospective analysis. *J Public Health Manag Pract*. 2020;26(2):116-123. doi:10.1097/PHH.0000000000000957
95. Nipah virus fact sheet. Accessed May 24, 2026. https://www.who.int/news-room/fact-sheets/detail/nipah-virus
96. Sarah Anne J. Guagliardo, C. Roxanne Connelly, Shelby Lyons, Stacey W. Martin, Rebekah Sutter, Holly R. Hughes, Aaron C. Brault, Amy J. Lambert, Carolyn V. Gould, J. Erin Staples. Reemergence of oropouche virus in the americas and risk for spread in the united states and its territories, 2024. *Emerging Infectious Diseases*. 2024;30(11). https://wwwnc.cdc.gov/eid/article/30/11/24-1220_article
97. Nathan D. Burkett-Cadena. Culicoides paraensis (insecta: Diptera: Ceratopogonidae), vector of oropouche virus. *EDIS*. 2025;(5). doi:10.32473/EDIS-IN1451-2025
98. Anne F. Payne, Jessica Stout, Peter Dumoulin, Timothy Locksmith, Lea A. Heberlein, Molly Mitchell, Arnold Rodriguez-Hilario, Alan P. Dupuis II, Alexander T. Ciota. Lack of competence of US mosquito species for circulating oropouche virus. *Emerging Infectious Diseases*. 2025;31(3). doi:10.3201/eid3103.241886
99. Mary Van Beusekom. Chikungunya moving into new regions, disabling millions and racking up billions in costs, data suggest. *CIDRAP News*. Published online 2024. https://www.cidrap.umn.edu/chikungunya/chikungunya-moving-new-regions-disabling-millions-and-racking-billions-costs-data
100. National typhoid and paratyphoid fever surveillance. Accessed May 24, 2026. https://www.cdc.gov/typhoid-fever/php/surveillance/
101. National typhoid and paratyphoid fever surveillance. Accessed May 24, 2026. https://www.cdc.gov/typhoid-fever/php/surveillance/index.html
102. Pertussis surveillance and trends. Accessed May 24, 2026. https://www.cdc.gov/pertussis/php/surveillance/index.html
103. NNDSS weekly data. Accessed May 24, 2026. https://data.cdc.gov/NNDSS/NNDSS-Weekly-Data/x9gk-5huc/about_data

104. Www.cdc.gov. Accessed May 24, 2026. https://www.cdc.gov/pertussis/media/pdfs/2026/02/363295-A_FS_PertussisSurveillanceReport_011626_508pass.pdf
105. Maps and statistics. Accessed May 26, 2026. https://www.cdc.gov/plague/maps-statistics/index.html
106. Www.sciencedirect.com. Accessed May 26, 2026. https://www.sciencedirect.com/science/article/pii/S1413867026010044
107. Community. Accessed May 24, 2026. https://www.ncbi.nlm.nih.gov/books/NBK430749/
108. Polio dashboard. Accessed May 24, 2026. https://www.ecdc.europa.eu/en/publications-data/polio-dashboard
109. Yu Sheng, Le-ying Jin, Ning Li, Yan Zhang, Ya-jun Shi. Global prevalence of psittacosis in outbreaks: A systematic review and meta-analysis. *BMC Public Health*. 2025;25:Article 21612. doi:10.1186/s12889-025-21612-y
110. Epidemiology and statistics. Accessed May 24, 2026. https://www.cdc.gov/q-fever/data-research/index.html
111. HK Miller, RA Priestley, GJ Kersh. Q fever: A troubling disease and a challenging diagnosis. *Clin Microbiol Newsl*. 2021;43(13):109-118. doi:10.1016/j.clinmicnews.2021.06.003
112. Www.cdc.gov. Accessed May 24, 2026. https://www.cdc.gov/q-fever/media/pdfs/rr6203.pdf
113. Philippe Carrara, Philippe Parola, Philippe Brouqui, Philippe Gautret. Imported human rabies cases worldwide, 1990–2012. *PLOS Neglected Tropical Diseases*. 2013;7(5):e2209. doi:10.1371/journal.pntd.0002209
114. Y He, Q Jia, K Cai, S Xu, H Li, Q Xie, others. The global, regional, and national burden of invasive non-typhoidal salmonella (iNTS): An analysis from the global burden of disease study 1990-2021. *PLoS Negl Trop Dis*. 2025;19(4):e0012960. doi:10.1371/journal.pntd.0012960
115. Centers for Disease Control and Prevention (CDC). Clinical overview of shigellosis. Published online 2024. https://www.cdc.gov/shigella/hcp/clinical-overview/index.html
116. Rocky mountain spotted fever (RMSF): Background, etiology, pathophysiology. Accessed May 24, 2026. https://emedicine.medscape.com/article/228042-overview
117. Data and maps for st. Louis encephalitis. Accessed May 24, 2026. https://www.cdc.gov/sle/data-maps/index.html
118. IH Spicknall, KM Kreisel, HS Weinstock. Estimates of the prevalence and incidence of syphilis in the united states, 2018. *Sex Transm Dis*. 2021;48(4):247-252. doi:10.1097/OLQ.0000000000001364
119. Sexually transmitted disease surveillance 2018. Accessed May 24, 2026. https://stacks.cdc.gov/view/cdc/79370
120. Syphilis. Accessed May 24, 2026. https://www.who.int/news-room/fact-sheets/detail/syphilis
121. About trichinellosis. Accessed May 24, 2026. https://www.cdc.gov/trichinellosis/about/index.html
122. S Gürcan. Epidemiology of tularemia. *Balkan Med J*. 2014;31(1):3-10. doi:10.5152/balkanmedj.2014.13117
123. Tularemia data and statistics. Accessed May 24, 2026. https://www.cdc.gov/tularemia/data-research/index.html
124. R Sharma, RD Patil, B Singh, S Chakraborty, D Chandran, K Dhama, others. Tularemia - a re-emerging disease with growing concern. *Vet Q*. 2023;43(1):1-16. doi:10.1080/01652176.2023.2277753
125. About vibrio infection. Accessed May 24, 2026. https://www.cdc.gov/vibrio/about/index.html
126. AA Irekeola, AA Alshehri. Human west nile virus infection: A meta-analysis of recent global data (2019-24). *J Glob Health*. 2025;15:04278. doi:10.7189/jogh.15.04278
127. Nicole S. Fijman, Donald A. Yee. Mapping yellow fever epidemics as a potential indicator of the historical range of aedes aegypti in the united states. *Memórias do Instituto Oswaldo Cruz*. 2022;117. doi:10.1590/0074-02760220306
128. U.S. Department of Health and Human Services (HHS). Yellow fever. Published online 2026. https://www.hhs.gov/immunization/diseases/yellow-fever/index.html
